# The CONSIDER Intervention Fidelity Framework for Complex Interventions in Healthcare: A “Best-Fit” Framework Synthesis

**DOI:** 10.1101/2024.08.29.24312797

**Authors:** Arsenio Páez, David Nunan, Peter McCulloch, David Beard

## Abstract

**Background:** The focus of clinical trials is typically interventions’ efficacy, or whether they attain their desired outcomes. Comparatively less attention is focused on understanding how or why interventions succeed, or fail to attain, those outcomes. This may be particularly important in trials of complex interventions such as surgery or physiotherapy, which are multifaceted and often tailored to individual participants, providers, or settings, increasing the potential for variations in intervention delivery and effects. The correspondence between the intervention that was planned and what was actually delivered in a trial is the intervention’s fidelity. Several benefits for high levels of intervention fidelity have been proposed. However, a lack of a uniform definition for fidelity and its key components may hinder intervention delivery in clinical trials and the translation of evidence-based interventions to clinical practice.

**Methods:** In this study, we undertook systematic review, and “Best-fit” framework synthesis to develop an empirically-based intervention fidelity framework for complex interventions in rehabilitation.

**Results:** The resulting **CONSIDER (Complex iNterventionS Design dElivery Recepit) framework** is first integrated fidelity framework developed specifically for clinical trials of for complex interventions rehabiliation. CONSIDER consists of three main components, Design, Delivery and Receipt and the factors moderating them. Design encompasses the core elements of the intervention and the protocol for the clinical trial to evaluate its effectiveness. Delivery encompasses the actual implementation of the protocol and treatment integrity. It is focused primarily on the actions of the intervention providers. Receipt addresses the exposure of the trial participants to the intervention and their response to it.

**Conclusions:** This fidelity framework is the first designed suit the unique complexities and challenges presented by physical complex interventions. It can help promote transparency and reproducibility and helps researchers design clinical trials that reduce waste, enable uptake into clinical practice, and benefit the practice and evidencing of physiotherapy, surgery and other physical complex interventions.

## BACKGROUND

Outside of safety, the focus of clinical trials is typically intervention efficacy, or whether or not interventions attain their desired outcomes. By comparison, less attention is focused on understanding how or why interventions succeed, or fail to attain, their target outcomes.^1,2^ This may be of particular importance in trials of complex interventions.^3–7^ The term “complex intervention” can be used to describe a number of multifaceted interventions in healthcare and other domains influencing health and well-being.^5^ These include interventions in psychology, mental and behavioural health, nursing, education, public health, social or public policy, among others. The focus of this paper is on complex interventions in physical domains in healthcare, specifically surgery and rehabilitation, defined here as physiotherapy, occupational and speech-language therapies, and exercise or physical activity interventions.^8,9^

Complex interventions, such as such as surgery and physiotherapy, involve a number of components which may act independently and interdependently to achieve some desired end.^5,7,10,11^ Practitioner skill and experience, learning curves, provider-patient expectations, differences in individual patient characteristics, biopsychosocial factors, and other factors can contribute to the outcome of complex interventions,^5–7^ sometimes antagonistically.^12^ For example, the effectiveness of surgical interventions may be enhanced, or degraded, by surgeons’ experience with the procedure, the degree to which procedure components have been defined, patients’ expectations, and the quality of perioperative supportive care.^13^

Intervention fidelity generally refers to the faithfulness of the intervention delivered in a clinical trial to the intervention that was intended in the trial protocol. Several benefits from high levels of intervention fidelity and negative consequences arising from poor intervention fidelity have been proposed. High levels of intervention fidelity can reduce random and unintended variability arising from poor intervention delivery, limiting potential confounding from extraneous variables, supporting internal validity^14^ and decrease the likelihood of type I and II errors.^14–18^ Attention to fidelity also decreases the likelihood of Type III error, or a null finding arising from poor quality intervention delivery rather than a null finding arising from intervention ineffectiveness.^19^

If treatment fidelity is poor or has not been evaluated, one may not be sure that studies’ significant results are attributable to the treatment, rather than other, unknown factors, creating in Type I error. If the results are not significant, one can’t assume that the poor results are attributable to the treatment rather than addition or omission of other factors, leading to Type II error.^15,20^ The addition of unplanned, extraneous components or omission of key intervention ingredients can make it difficult to attribute observed effects to the action of the intervention.^13,21^ Poor intervention delivery (fidelity) can lead to nonsignificant outcomes resulting from poor intervention delivery, rather than an actual lack of intervention effectiveness, raising the risk of Type III error.^22,23^

The degree of intervention fidelity achieved in a study may be of equal clinical value with quantitative changes when interpreting the results of clinical trials in complex interventions.^24^ However, a number of overlapping terms and constructs are used to describe and operationalise intervention fidelity, hampering efforts to support and monitor it in clinical trials.^25–27^ A variety of frameworks have been developed to understand and monitor intervention fidelity in complex clinical trials, but these vary in content and focus. There is also little consensus on how best to define intervention fidelity or categorize its key components, further complicating efforts to enhance or monitor intervention fdielity.^25,26,28–30^ Those who develop, implement and study complex interventions have no common language by which they can make assessments and develop robust methods to support intervention fidelity.^25,29,31,32^

Updates to the CONSORT^33^ reporting guidelines (2010), The Standard Protocol Items: Recommendations for Interventional Trials (SPIRIT)^34^ statement (2013), Template for Intervention Description and Replication (TIDieR) Checklist^35^ (2014), and the Consensus on Exercise Reporting Template (CERT)^36^ (2016) were developed to improve intervention reporting and include items related to fidelity. However, they provide limited guidance for reporting strategies to maintain participants adherence to their assigned intervention or document modifications to intervention allocation, and no guidance on strategies to monitor or support the quality of intervention delivery. These checklists also only provide guidance on what should be reported and were not intended to offer researchers a practical guide on how to approach intervention fidelity in study design or conduct.^37^ What was *not* done in a trial cannot be reported.

Treatment effects observed in trials will not be reproducible unless well-described procedures facilitate and maintain intervention fidelity in clinical trials.^13,38,39^ Published fidelity frameworks have been developed for use in behavioural medicine, public health and education, but evidence-based guidance for fidelity monitoring in clinical trials of physical complex interventions such as surgery, physiotherapy, and rehabilitation is rare.^29,31,40,41^ A comprehensive fidelity framework for clinical trials in domains involving physical complex interventions, such as physiotherapy and rehabilitation (physical medicine and rehabilitation, broadly defined) is needed as a framework from which to investigate intervention fidelity in this complex interventions in rehabilitation.

A conceptual intervention fidelity framework is made up of a set of guidelines or recommendations detailing a combination of strategies and methods to assess, enhance and evaluate intervention fidelity at different stages of an intervention’s implementation during a clinical trial.^42^ In this chapter, the development of the COmplex iNterventions trialS fIDElity fRamework (CONSIDER) framework, a conceptual framework for intervention fidelity in clinical trials of complex interventions in the physical domain, is described. Further development of this framework through broader input from a wider range of stakeholders could support the design and conduct of more robust complex intervention clinical trials. ^1,32,43^

### Objectives

1. To provide a description of fidelity constructs, models, frameworks reporting the delivery of complex interventions in controlled trials in physical domains of health care: physiotherapy, surgery, physical medicine, rehabilitation, occupational therapy, speech and language therapy,
2. exercise, and physical activity promotion.
3. To conduct a best-fit framework synthesis of fidelity constructs to inform a conceptual fidelity
4. framework from which to investigate fidelity in trials of complex interventions in physical domains.
5. To produce an integrated, empirically based definition of intervention fidelity for use in clinical trials of complex interventions in healthcare.

## METHODS

### Reporting standards

The systematic review and framework analysis are reported along the Preferred Reporting Items for Systematic Reviews and Meta-Analyses (PRISMA) ^44^ and **EN**hancing **T**ransparency in **RE**porting the synthesis of **Q**ualitative research (ENTREQ) statement guidelines.^45^

### Registration

The protocol was registered on PROSPERO,^46^ (https://www.crd.york.ac.uk/prospero/display_record.php?ID=CRD42019135957) following the Preferred Reporting Items for Systematic Reviews and Meta-Analyses for Protocols (PRISMA-P) guidelines.^44^

### Systematic Review Design

The review followed a Best-fit Framework Synthesis (BFFS) approach to develop a comprehensive fidelity framework based on existing evidence (Fig. 2.1).^47^ Best-fit framework syntheses use initially-identified conceptual models found in published literature to serve as a platform from which a new framework can be developed through thematic synthesis of evidence from empirical studies.^47–50^ The approach is augmentative and deductive, building iteratively on the existing models.^50^ We chose to create a new intervention fidelity framework though BFFS as, to the best of our knowledge, no intervention fidelity frameworks had previously been developed to guide fidelity in clinical trials of complex interventions in the physical domain.

**Figure 2.1:**
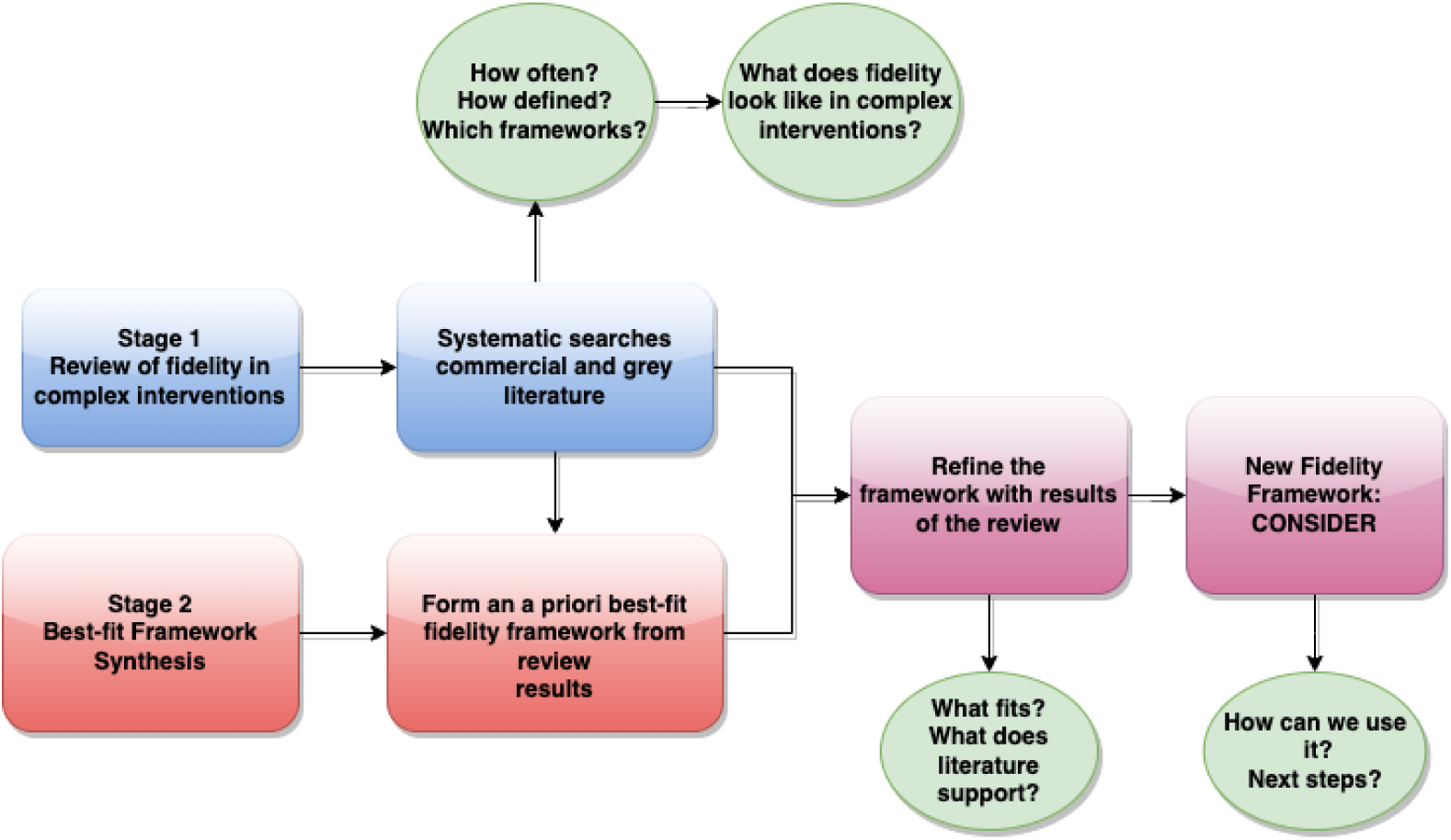
Stages of the review and best-fit framework synthesis.

Very detailed academic methodology was needed to construct the framework (part one). To enable it to be useful, it also had to be distilled into useful portions, headings, and silos that can be easily illustrated and recalled. The review is therefore divided into two stages (figure 2.1).

### Stage 1: Create the platform (“Best-fit” framework) on which to build the new framework

Once the research question or aim was determined, the creation of the *a priori*, best-fit framework (a platform from which to build a new framework) was conducted in parallel with systematic searches for papers to be included in the review and synthesis. These two “strands” then joined at the framework synthesis stage (stage 2).

### Stage 2: Build on the platform with empirical evidence to make a new framework

In stage 2, fidelity frameworks, theories, models and concepts identified in stage 1 were thematically synthesized and reciprocally translated through BFFS (figure 2.2) ^47,50^ to develop an integrated, conceptual fidelity framework for complex interventions in the physical domain. This new framework can subsequently be honed and validated in future empirical research.

**Figure 2.2:**
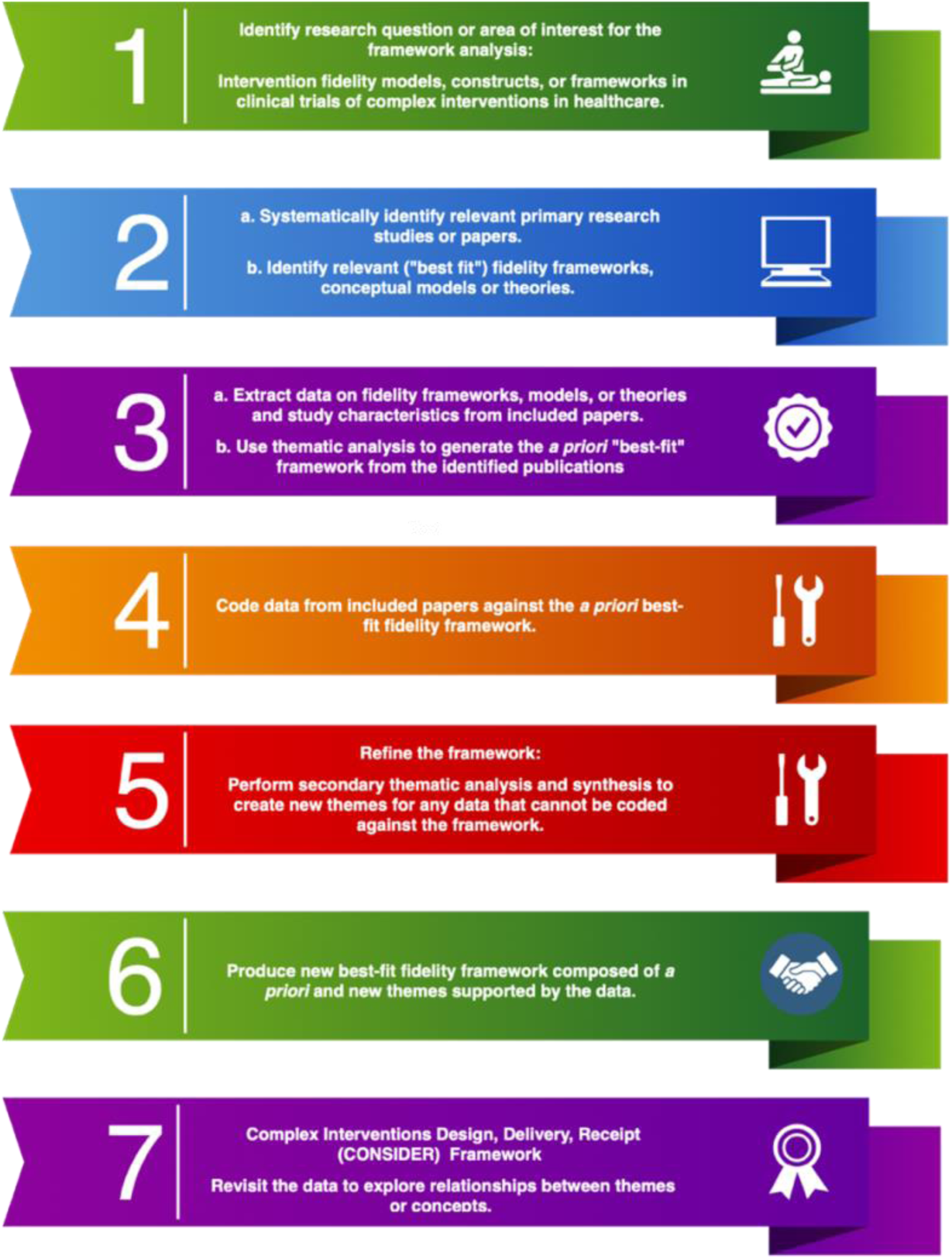
Best-Fit Framework Analysis’ Steps: Adapted from Booth, et al, 2014^59^.

#### Stage I: Systematic review

##### Search Strategy

As we were searching for theoretical frameworks, the search strategy was informed by the BeHEMoth (Behaviour of interest, health context, exclusions, and models or theories) method.^48,51^ ^49^

**Be:** Treatment or intervention fidelity, adherence, integrity, compliance, concordance, implementation, and related concepts.

**H:** Complex interventions in the physical domain in healthcare: physiotherapy, occupational therapy, speech-language therapy, exercise or physical activity interventions, surgery.

**E:** Statistical or economic models, models of care. Non-healthcare interventions.

**MoTh:** Model, theory, theories, framework, concept, conceptual, construct or strategy.

Keywords and MeSH terms related to fidelity (table 2.1) were used in Pubmed, Embase, CINHAL, Scopus and Google Scholar (appendix figure 2.1). PROSPERO was searched for ongoing or recently completed systematic reviews. Searches were limited to papers published from 2005, a year after the publication of the NIH Behaviour Change Consortium (NIH BCC) recommendations for conceptualizing and enhancing treatment fidelity.^52^ No language limits were imposed on searches. To optimize coverage and ensure literature saturation, grey literature searching was conducted on Scopus, Google Scholar, and PROSPERO. Grey literature includes a wide range of data sources not typically captured in searches for commercially published literature, including clinical trial and protocol registers such as clinicaltrials.gov^53^, conference proceedings, government reports, not-for-profit or non-governmental agency reports, international bodies’ reports or position papers (e.g., the World Health Organisation).^54^ The reference lists of also searched for additional papers. Study protocols or trial registrations were also searched for and reviewed when they were available.

**Table 2.1:**
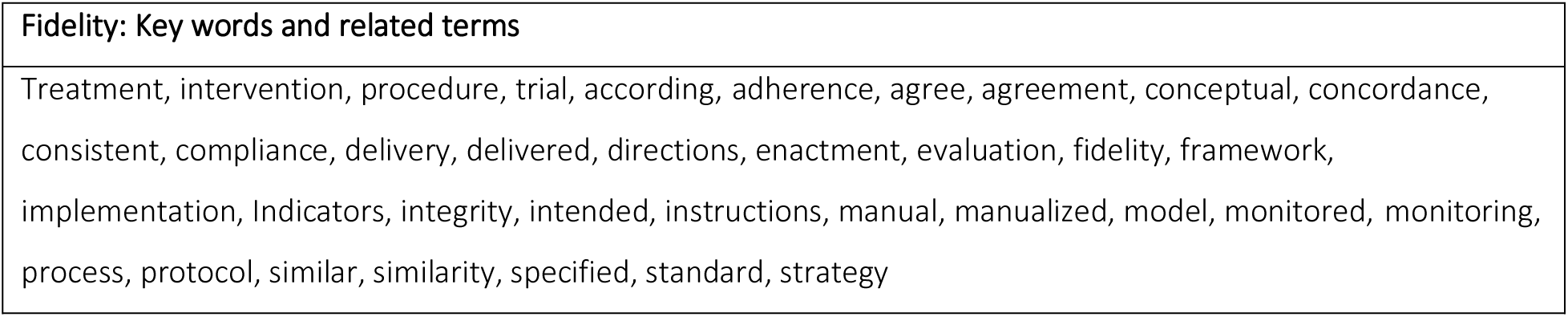
Fidelity keywords and related terms.

##### Inclusion criteria

The aim of the systematic review was to identify as many relevant studies as possible and reduce the risk of missing potentially eligible studies, maximizing sensitivity rather than precision.

Eligible papers were complex interventions empirical research, review, or theoretical papers including terms related to fidelity anywhere in the paper, either as a main focus or component (for example, as an analysis within a trial or process evaluation).

##### Exclusion criteria

Papers not describing fidelity, or a related term, were excluded (table 2.1). Papers investigating fidelity outside of complex interventions in the physical domain in healthcare were ineligible. These included papers for public-health interventions, such as smoking cessation or reproductive health interventions, interventions in education, such as reading proficiency, or in psychology, such as interventions for compulsive behaviour or depression.

##### Study selection

Potentially relevant citations, their abstracts and full-texts were screened against the review’s inclusion-exclusion criteria independently by the first author (AP) and a second author (DH). Disagreements were resolved by consensus. As the BFFS aimed for conceptual saturation and generalizability, rather than statistical generalizability, we aimed for study selection that was purposive rather than exhaustive.^55,56^ A criterion-sampling approach to purposive sampling was used to select papers from the systematic review for framework synthesis.^56,57^

##### Data Collection and Extraction Process

Citations, abstracts, and full text articles were managed digitally with Mendeley. A tailored, electronic data extraction form was created and calibrated for this review. We extracted papers’ bibliographic information, field, professional discipline or context, study design or paper format (e.g., methodological paper), descriptions or definitions of fidelity, fidelity monitoring or support methods, and fidelity model or framework if reported (Appendix table 2.1).

##### Quality assessment

Little consensus exists for the feasibility and utility of quality assessment in framework analysis or qualitative synthesis.^58^ As this systematic review sought to synthesize fidelity frameworks and models, rather than the outcomes or effects of processes or procedures in empirical work, no quality assessment of papers was performed in this review.

##### Data Synthesis

Objective 1: The results of the systematic review were described narratively and with simple descriptive statistics. No quantitative synthesis was undertaken.

Objective 2: Fidelity models or frameworks selected through purposive sampling were thematically synthesized with NVivo12 software.^47,50^

#### Stage II. Build on the platform to make a new framework (Best-Fit Framework Synthesis)

With the results of the systematic review, the “Best-fit Framework Synthesis (BFFS) ^47,50^” method was used following a series of predetermined steps (figure 2.2).^59^ As an overview, once an initial “platform” best-fit framework was constructed, fidelity data (fidelity descriptions, definitions, monitoring and enhancement) extracted from papers identified in the systematic review was thematically analysed and coded against the best-fit framework to expand or reduce it with the data to creating a new fidelity framework.^47–50^

Thematic analysis followed the guidance and recommended steps of Saldana’s (2016) Coding Manual for Qualitative Researchers, and the Streamlined Codes-to Theory Model for qualitative inquiry.^60^ First-cycle coding followed a descriptive (topic) coding method, generating a categorized inventory of codes identified and themed through careful reading and reflection on the data.^61^ In second-cycle coding, pattern coding was used to reanalyse and reconfigure the first-cycle codes categorically, thematically, and conceptually into a smaller number of themes (pattern codes) to develop an understanding of the corpus of data and relationships between its components. ^62^

##### Best-fit framework: steps

###### Step 1: Identify the review question or aim

**Aim:** to synthesize a conceptual framework for intervention fidelity in clinical trials of complex interventions in the physical domain (physiotherapy, surgery, rehabilitation).

###### Step 2: Systematically identify relevant research and “best-fit” frameworks, models, or theories

Comprehensive, systematic searches were conducted to identify as many relevant publications addressing the question as was feasible (Stage I). In a parallel process, these papers were reviewed to identify a best-fit “platform” fidelity models (Step 2b).^59^

###### Step 3: Generate the *a priori* framework from identified publications through thematic analysis

This *a priori* “best-fit” framework serves as a platform or starting point that will be built on with fidelity data from eligible papers to synthesise a new fidelity framework in the next steps.

###### Step 4: Code evidence from included studies against the best-fit framework

Once the BFF was constructed, passages describing fidelity in eligible papers were extracted as direct quotations and imported into in NVivio12. These quotations were systematically reviewed against the fidelity concepts categorised in the BFF, feeding them into the BFF theme they best represented.^63^ This “thematic coding” was performed and critically reviewed by two reviewers (AP and DH) and any discrepancies or disagreements were resolved by consensus.

###### Step 5: Create new themes

When fidelity data from the extracted quotations did not translate well into any of the BFF concepts (themes) or were applicable to more than one, new themes were created through interpretive, inductive secondary thematic analysis^47,64^ and reciprocal translation (Step 5).^65^ The criterion for forming a theme was that at least two quotations addressing the same concept were identified, and they did not correspond to an existing fidelity theme. The BFF was expanded upon, reduced, or added to iteratively as successive studies were analysed and data was coded, ultimately creating a revised fidelity framework (Step 6).^47,50,66^

###### Step 6: Produce a new framework composed of *a priori* and new themes

Consensus was sought among the reviewers (authors) on which of the BFF fidelity themes were supported, and whether quotations extracted from eligible papers mapped onto a pre-existing theme, or themes mapped onto each other (could be collapsed) through reciprocal translation. This, and steps 4-5 resulted in a new framework, included some fidelity themes from the BFF and new themes derived from fidelity data extracted from the results of the systematic review.

###### Step 7: Revisit evidence to explore the relationship between fidelity themes or concepts

The resulting finalized list of fidelity themes was used to create a new, integrated fidelity framework for physical complex interventions. An interpretation of the content of the fidelity themes, and relationships between them, is presented in the results section.

## RESULTS: Systematic Review

Searches produced 2857 records, 361 of which were screened for eligibility by full text (figure 2.3). Of these, 269 met inclusion criteria in the systematic review. One hundred forty-one papers defined or described fidelity, fidelity components or a fidelity framework, meeting criteria for informing the framework synthesis.

**Figure 2.3:**
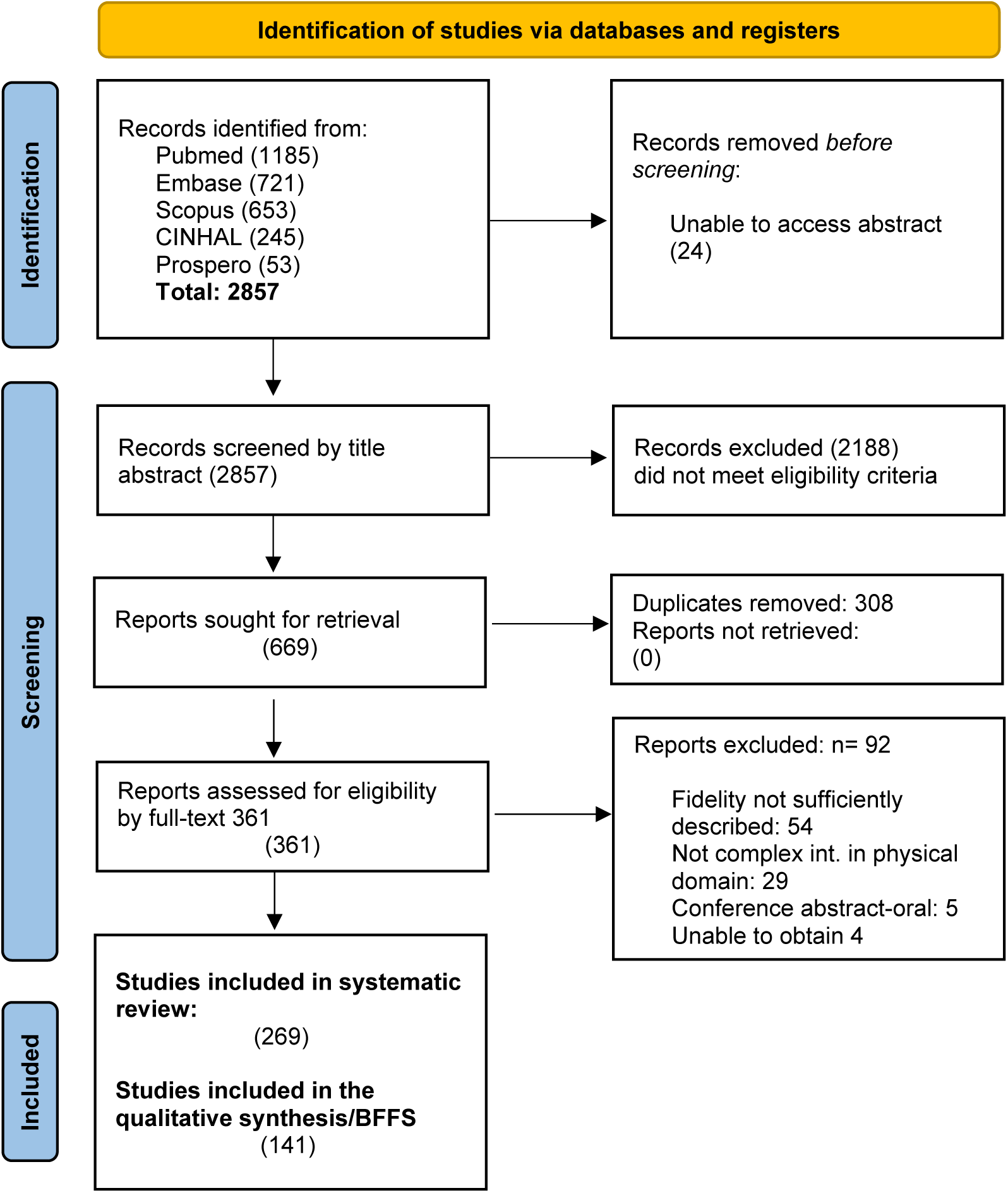
PRISMA flow chart.

## OBJECTIVE 1: Fidelity in complex interventions

The greatest number of eligible papers (73) came from physiotherapy literature (table 2.2). The least represented specialty was surgery, with 16 papers, including three randomised controlled trials (RCTs) of a surgical procedure^67–69^, two surgical trial protocols^70,71^, one pilot ^72^, three trials of peri-operative surgical care or procedures^73–75^, two systematic reviews of methodological reporting in surgical placebo-controlled trials^76,77^, two guidelines on the conduct of surgical trials^13,78^, two methodological papers^76,79^, and one set of reporting standards.^80^

**Table 2.2:** Included studies according to discipline and study design/type.

| Clinical discipline | Eligible papers | Study design or type of paper | Eligible papers |
| --- | --- | --- | --- |
| Physical Therapy | 73 | Protocol paper | 51 |
| Physical activity interventions | 66 | Randomized, parallel group trials | 47 |
| Rehabilitation (multidisciplinary) | 37 | Feasibility studies and trials | 35 |
| Occupational Therapy | 35 | Fidelity/Implementation studies, process evaluations | 33 |
| Speech Language Pathology | 25 | Literature or narrative reviews | 28 |
| Cardiopulmonary Rehabilitation | 17 | Methodological papers | 27 |
| Surgery and surgical care | 16 | Systematic reviews and Meta-Analyses | 23 |
|  |  | Case and pilot studies | 19 |
|  |  | Uncontrolled studies | 6 |
| Total | 269 | Total | 269 |

## OBJECTIVE 2: A best-fit framework synthesis

### BFFS Steps 1-3: Fidelity frameworks

Steps 1-2b identified 28 fidelity frameworks or constructs across 108 papers. The two most frequently cited or applied fidelity frameworks were the National Institutes of Health Behaviour Change Consortium (NIH-BCC) Comprehensive Treatment Fidelity Framework^81^ as described by Bellg^52^ or Borelli^81^ (60 papers cited or applied), and the Comprehensive Intervention Fidelity Guide developed by Gearing^29^ (23 papers cited or applied (table 2.3). The next most cited (19 papers) was the Implementation Fidelity Framework by Carroll et al (2007)^2^. The NIH-BCC and Gearing frameworks were thematically analysed to identify commonalities and differences between them and generate fidelity themes supported with examples extracted from the original paper (Table 2.3). These themes generated the best-fit framework (BFF) that served as a starting point, or platform, from which to build a new fidelity framework with fidelity data extracted from eligible papers (Step 4).^47,50,59,66^

**Table 2.3:**
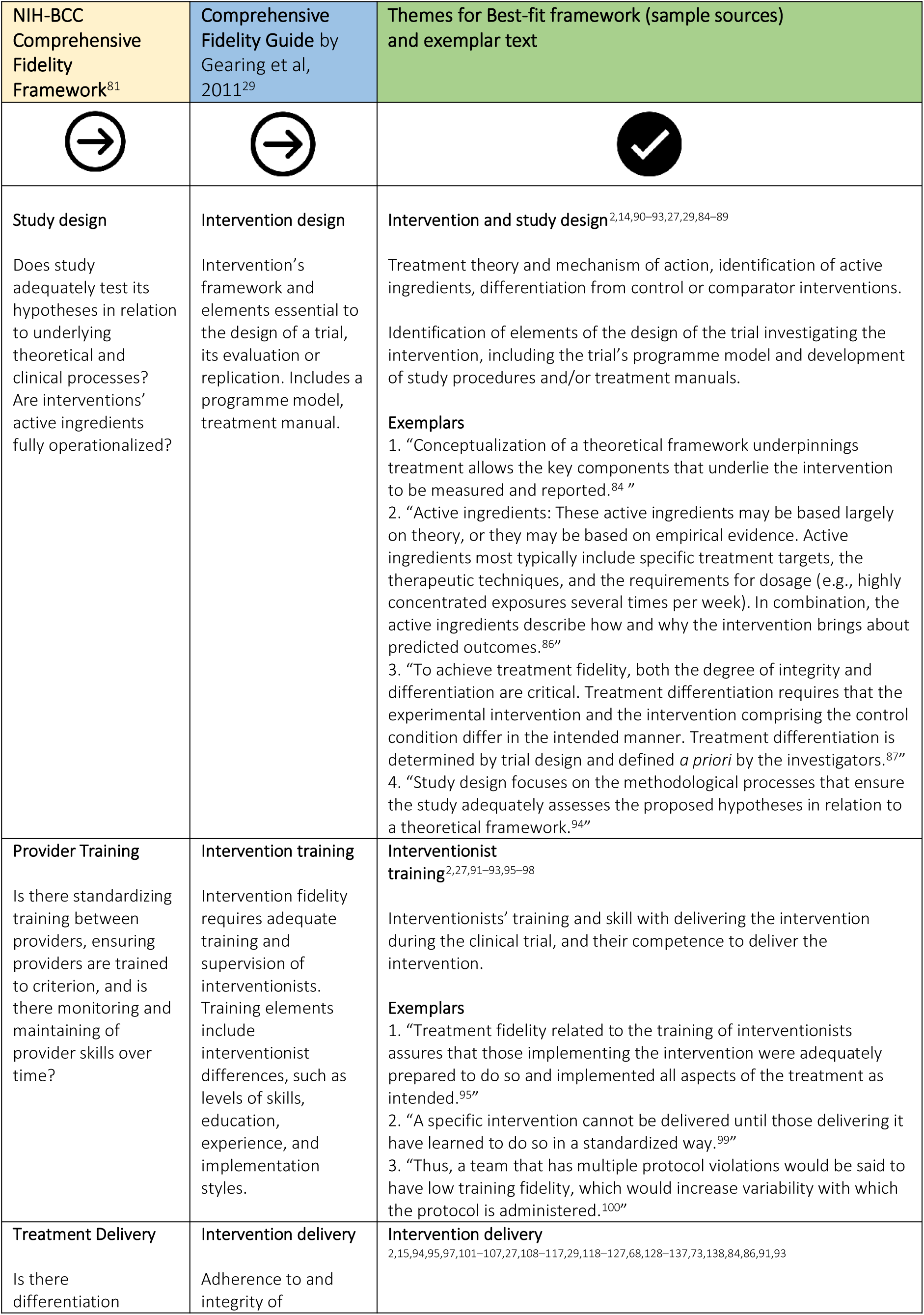
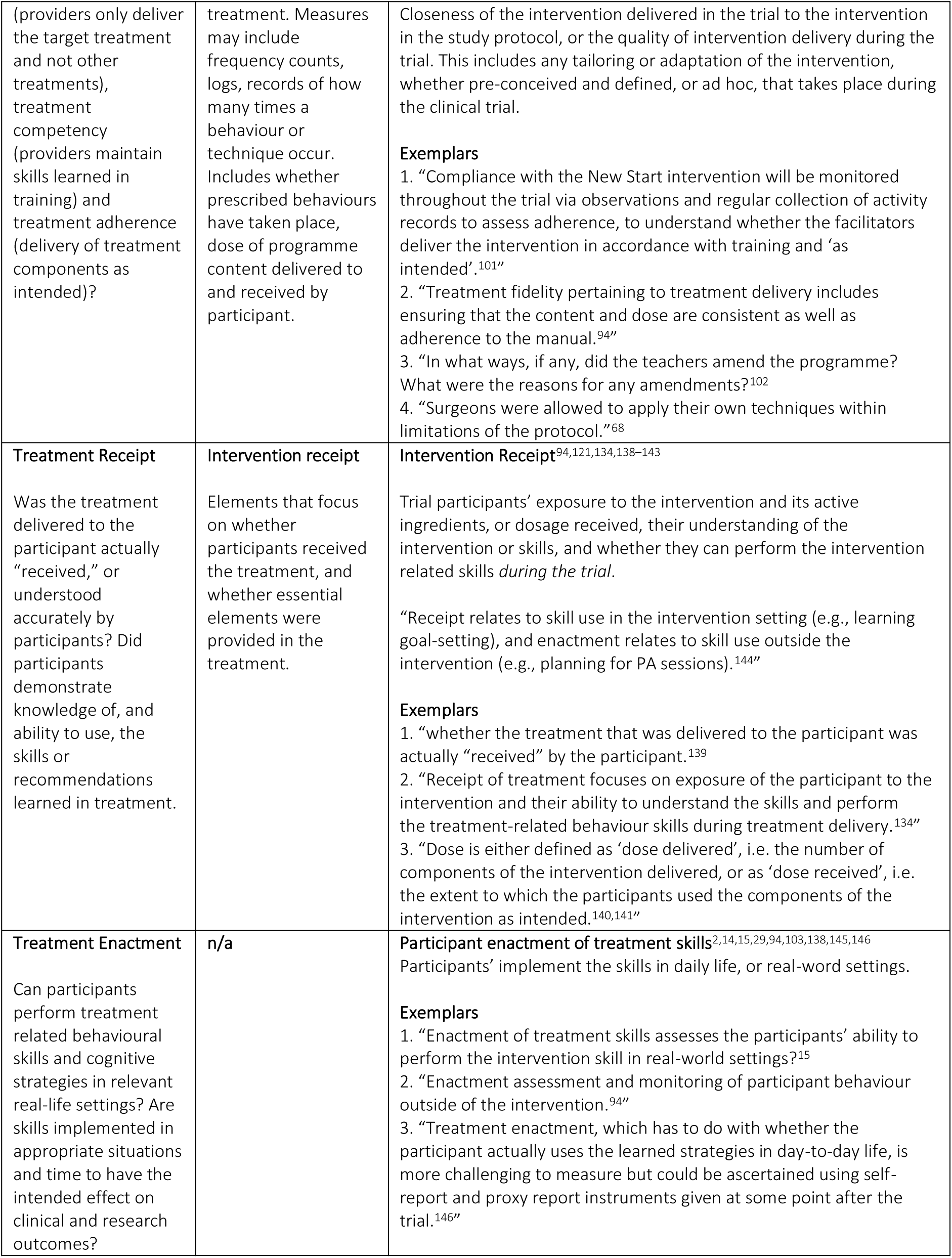
NIH-BCC, Gearing’s Comprehensive Intervention Fidelity Guide Fidelity, and Best-fit framework Frameworks.

| NIH-BCC Comprehensive Fidelity Framework<sup>81</sup> | Comprehensive Fidelity Guide by Gearing et al, 2011<sup>29</sup> | Themes for Best-fit framework (sample sources) and exemplar text |
| --- | --- | --- |
| **Study design**<br>Does study adequately test its hypotheses in relation to underlying theoretical and clinical processes? Are interventions' active ingredients fully operationalized? | **Intervention design**<br>Intervention's framework and elements essential to the design of a trial, its evaluation or replication. Includes a programme model, treatment manual. | **Intervention and study design**<sup>2,14,90–93,27,29,84–89</sup><br>Treatment theory and mechanism of action, identification of active ingredients, differentiation from control or comparator interventions.<br>Identification of elements of the design of the trial investigating the intervention, including the trial's programme model and development of study procedures and/or treatment manuals.<br>**Exemplars**<br>1. "Conceptualization of a theoretical framework underpinnings treatment allows the key components that underlie the intervention to be measured and reported.<sup>84</sup>"<br>2. "Active ingredients: These active ingredients may be based largely on theory, or they may be based on empirical evidence. Active ingredients most typically include specific treatment targets, the therapeutic techniques, and the requirements for dosage (e.g., highly concentrated exposures several times per week). In combination, the active ingredients describe how and why the intervention brings about predicted outcomes.<sup>86</sup>"<br>3. "To achieve treatment fidelity, both the degree of integrity and differentiation are critical. Treatment differentiation requires that the experimental intervention and the intervention comprising the control condition differ in the intended manner. Treatment differentiation is determined by trial design and defined *a priori* by the investigators.<sup>87</sup>"<br>4. "Study design focuses on the methodological processes that ensure the study adequately assesses the proposed hypotheses in relation to a theoretical framework.<sup>94</sup>" |
| **Provider Training**<br>Is there standardizing training between providers, ensuring providers are trained to criterion, and is there monitoring and maintaining of provider skills over time? | **Intervention training**<br>Intervention fidelity requires adequate training and supervision of interventionists. Training elements include interventionist differences, such as levels of skills, education, experience, and implementation styles. | **Interventionist training**<sup>2,27,91–93,95–98</sup><br>Interventionists' training and skill with delivering the intervention during the clinical trial, and their competence to deliver the intervention.<br>**Exemplars**<br>1. "Treatment fidelity related to the training of interventionists assures that those implementing the intervention were adequately prepared to do so and implemented all aspects of the treatment as intended.<sup>95</sup>"<br>2. "A specific intervention cannot be delivered until those delivering it have learned to do so in a standardized way.<sup>99</sup>"<br>3. "Thus, a team that has multiple protocol violations would be said to have low training fidelity, which would increase variability with which the protocol is administered.<sup>100</sup>" |
| **Treatment Delivery**<br>Is there differentiation | **Intervention delivery**<br>Adherence to and integrity of | **Intervention delivery**<br><sup>2,15,94,95,97,101–107,27,108–117,29,118–127,68,128–137,73,138,84,86,91,93</sup> |
| (providers only deliver the target treatment and not other treatments), treatment competency (providers maintain skills learned in training) and treatment adherence (delivery of treatment components as intended)? | treatment. Measures may include frequency counts, logs, records of how many times a behaviour or technique occur. Includes whether prescribed behaviours have taken place, dose of programme content delivered to and received by participant. | Closeness of the intervention delivered in the trial to the intervention in the study protocol, or the quality of intervention delivery during the trial. This includes any tailoring or adaptation of the intervention, whether pre-conceived and defined, or ad hoc, that takes place during the clinical trial.<br>**Exemplars**<br>1. "Compliance with the New Start intervention will be monitored throughout the trial via observations and regular collection of activity records to assess adherence, to understand whether the facilitators deliver the intervention in accordance with training and 'as intended'.<sup>101</sup>"<br>2. "Treatment fidelity pertaining to treatment delivery includes ensuring that the content and dose are consistent as well as adherence to the manual.<sup>94</sup>"<br>3. "In what ways, if any, did the teachers amend the programme? What were the reasons for any amendments?<sup>102</sup>"<br>4. "Surgeons were allowed to apply their own techniques within limitations of the protocol."<sup>68</sup> |
| **Treatment Receipt** | **Intervention receipt** | **Intervention Receipt<sup>94,121,134,138-143</sup>** |
| Was the treatment delivered to the participant actually "received," or understood accurately by participants? Did participants demonstrate knowledge of, and ability to use, the skills or recommendations learned in treatment. | Elements that focus on whether participants received the treatment, and whether essential elements were provided in the treatment. | Trial participants' exposure to the intervention and its active ingredients, or dosage received, their understanding of the intervention or skills, and whether they can perform the intervention related skills *during the trial*.<br>"Receipt relates to skill use in the intervention setting (e.g., learning goal-setting), and enactment relates to skill use outside the intervention (e.g., planning for PA sessions).<sup>144</sup>"<br>**Exemplars**<br>1. "whether the treatment that was delivered to the participant was actually "received" by the participant.<sup>139</sup>"<br>2. "Receipt of treatment focuses on exposure of the participant to the intervention and their ability to understand the skills and perform the treatment-related behaviour skills during treatment delivery.<sup>134</sup>"<br>3. "Dose is either defined as 'dose delivered', i.e. the number of components of the intervention delivered, or as 'dose received', i.e. the extent to which the participants used the components of the intervention as intended.<sup>140,141</sup>" |
| **Treatment Enactment** | n/a | **Participant enactment of treatment skills<sup>2,14,15,29,94,103,138,145,146</sup>** |
| Can participants perform treatment related behavioural skills and cognitive strategies in relevant real-life settings? Are skills implemented in appropriate situations and time to have the intended effect on clinical and research outcomes? |  | Participants' implement the skills in daily life, or real-world settings.<br>**Exemplars**<br>1. "Enactment of treatment skills assesses the participants' ability to perform the intervention skill in real-world settings?<sup>15</sup>"<br>2. "Enactment assessment and monitoring of participant behaviour outside of the intervention.<sup>94</sup>"<br>3. "Treatment enactment, which has to do with whether the participant actually uses the learned strategies in day-to-day life, is more challenging to measure but could be ascertained using self-report and proxy report instruments given at some point after the trial.<sup>146</sup>" |

The BFF was comprised of five fidelity themes (table 2.3).^31^ Each theme was supported with a definition and supporting exemplars from the original papers. Fidelity data extracted from eligible papers was later coded against these themes in step 4.^47^ The resulting BFF described fidelity on both a theoretical and an operational level (table 2.4).^31^ The theoretical level addresses construct fidelity, or interventions’ faithfulness to their underlying theoretical basis. Similar to Meehl and Cronbach’s^82^ concept of “construct validity,” encompassing relations between hypothesized entities and processes and observed effects, the theoretical level encompasses interventions’ design, proposed mechanisms of action, and their relation to the interventions’ core effects.^29,83^ The operational level encompasses interventions’ fidelity in terms of the quality of intervention delivery, receipt (dosage) of interventions, participant engagement, and participant enactment, or whether participants apply intervention skills in their daily lives outside of treatment sessions.^29^

**Table 2.4:**
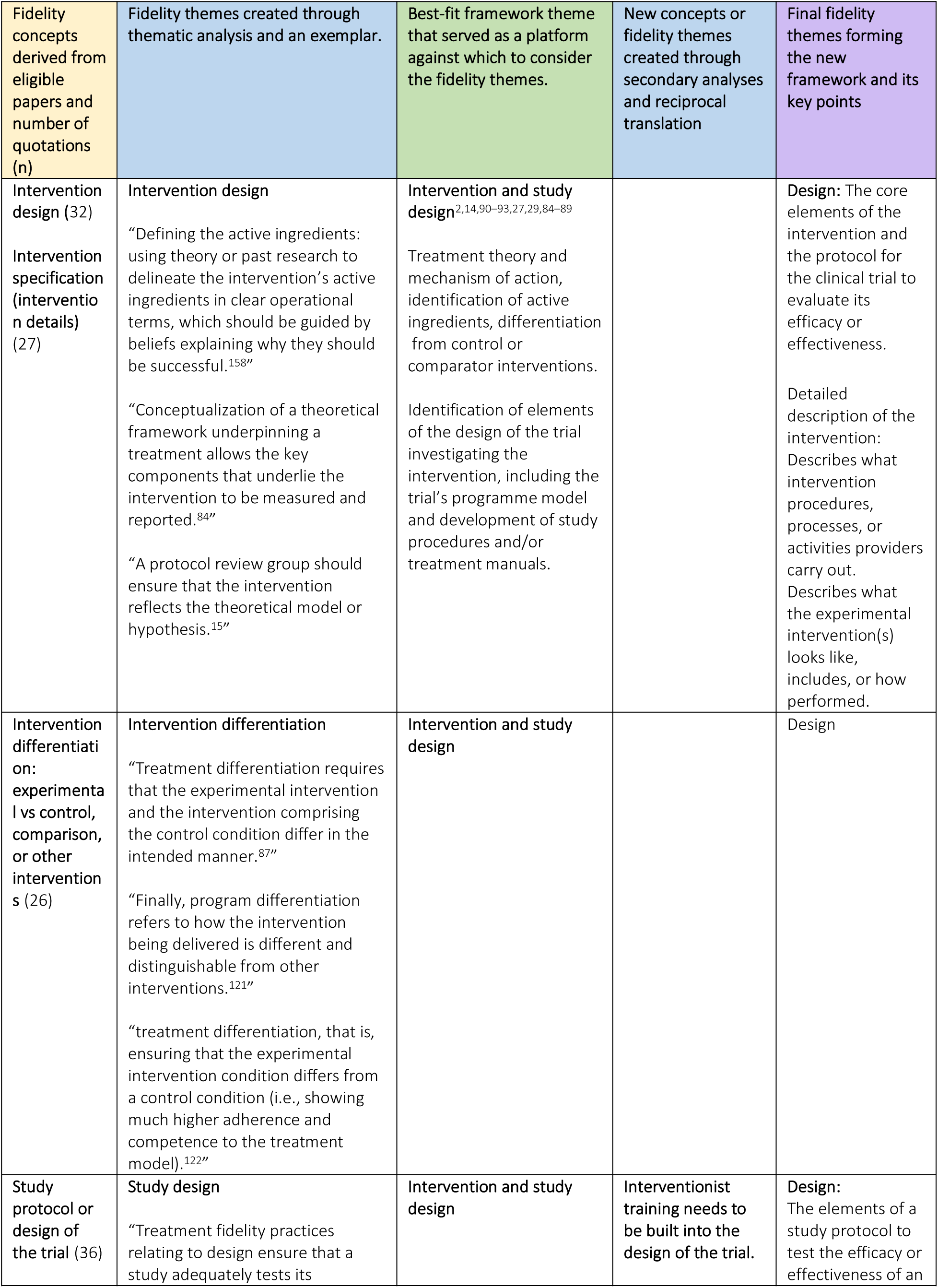
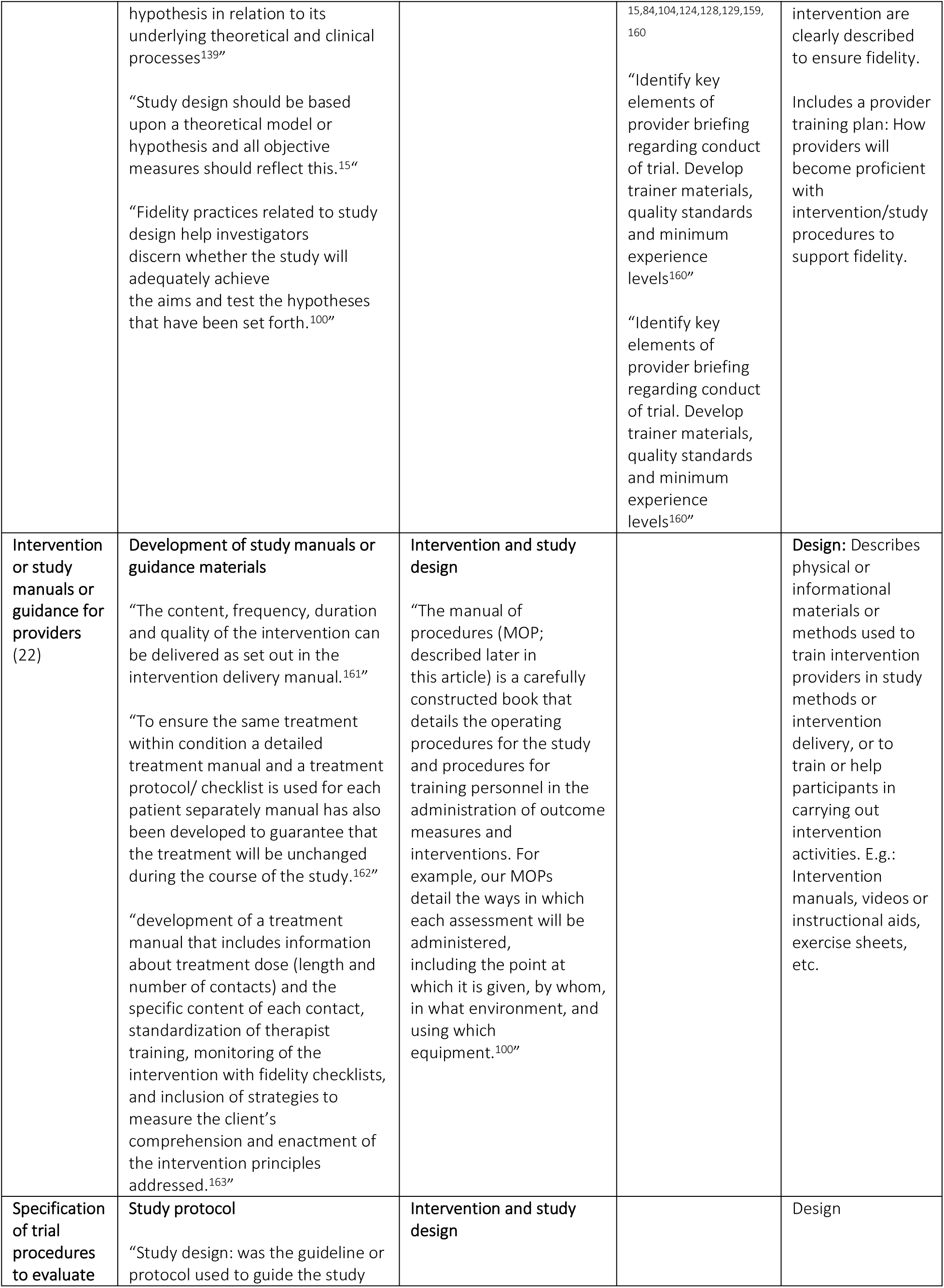
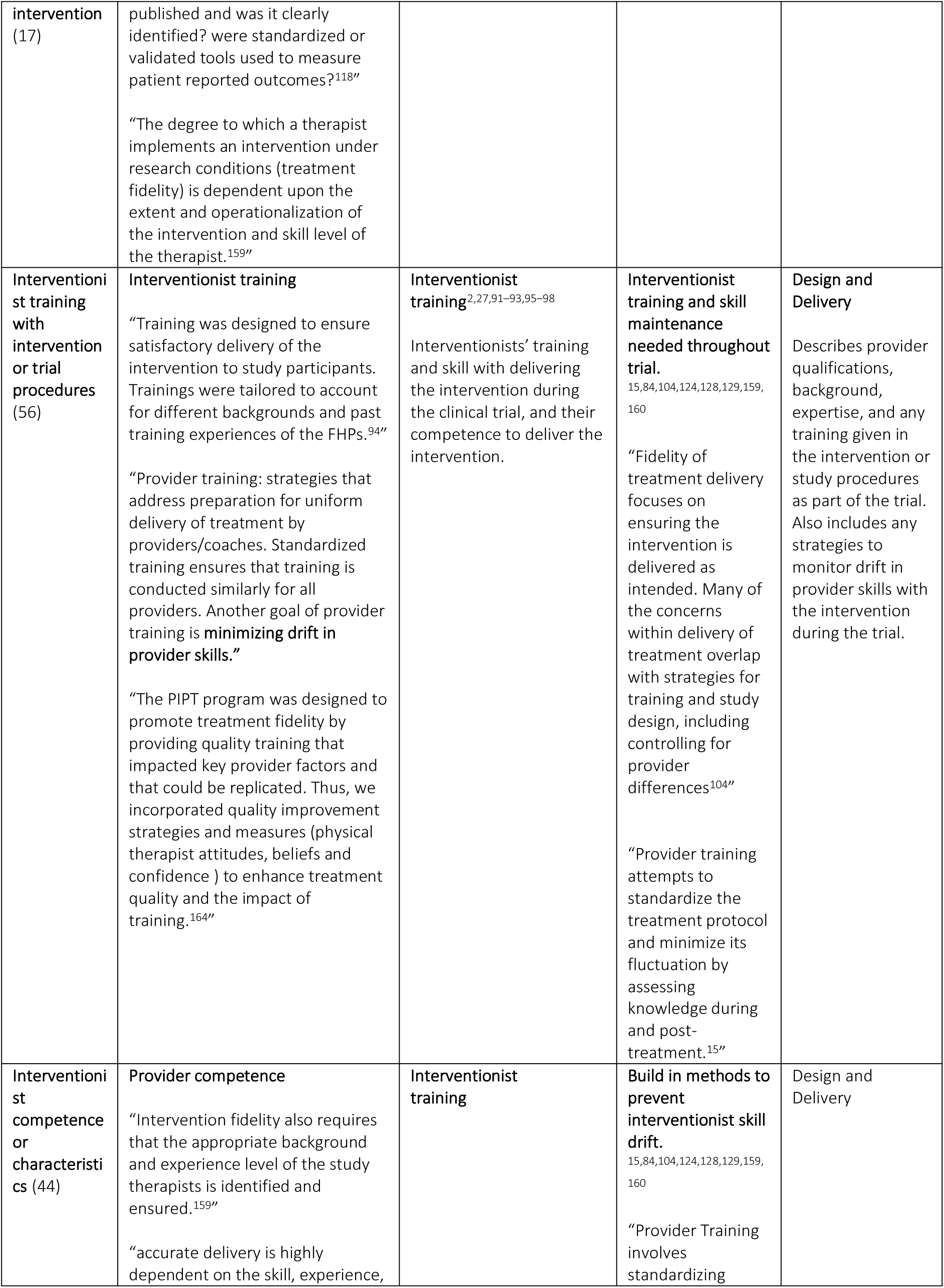
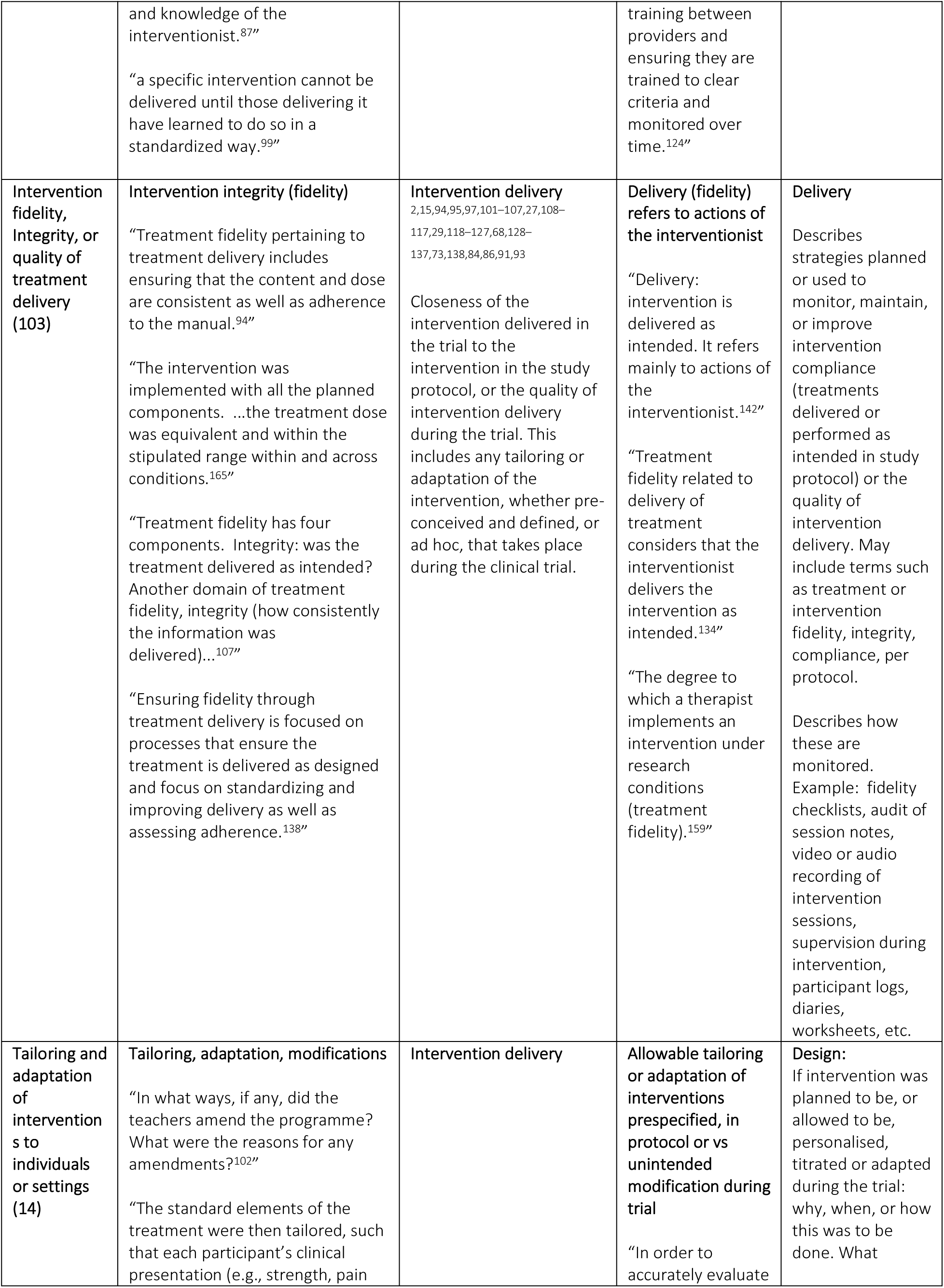
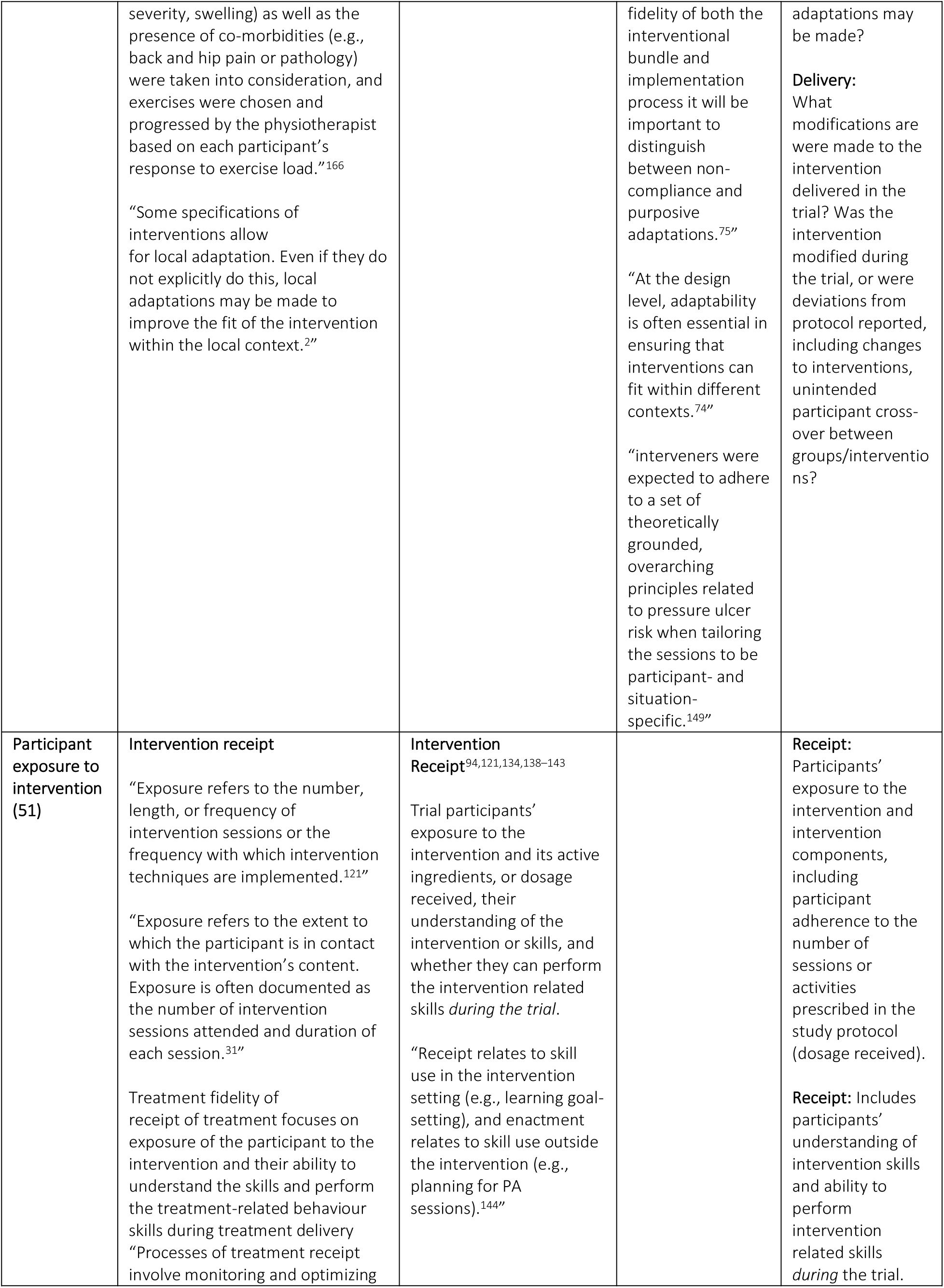
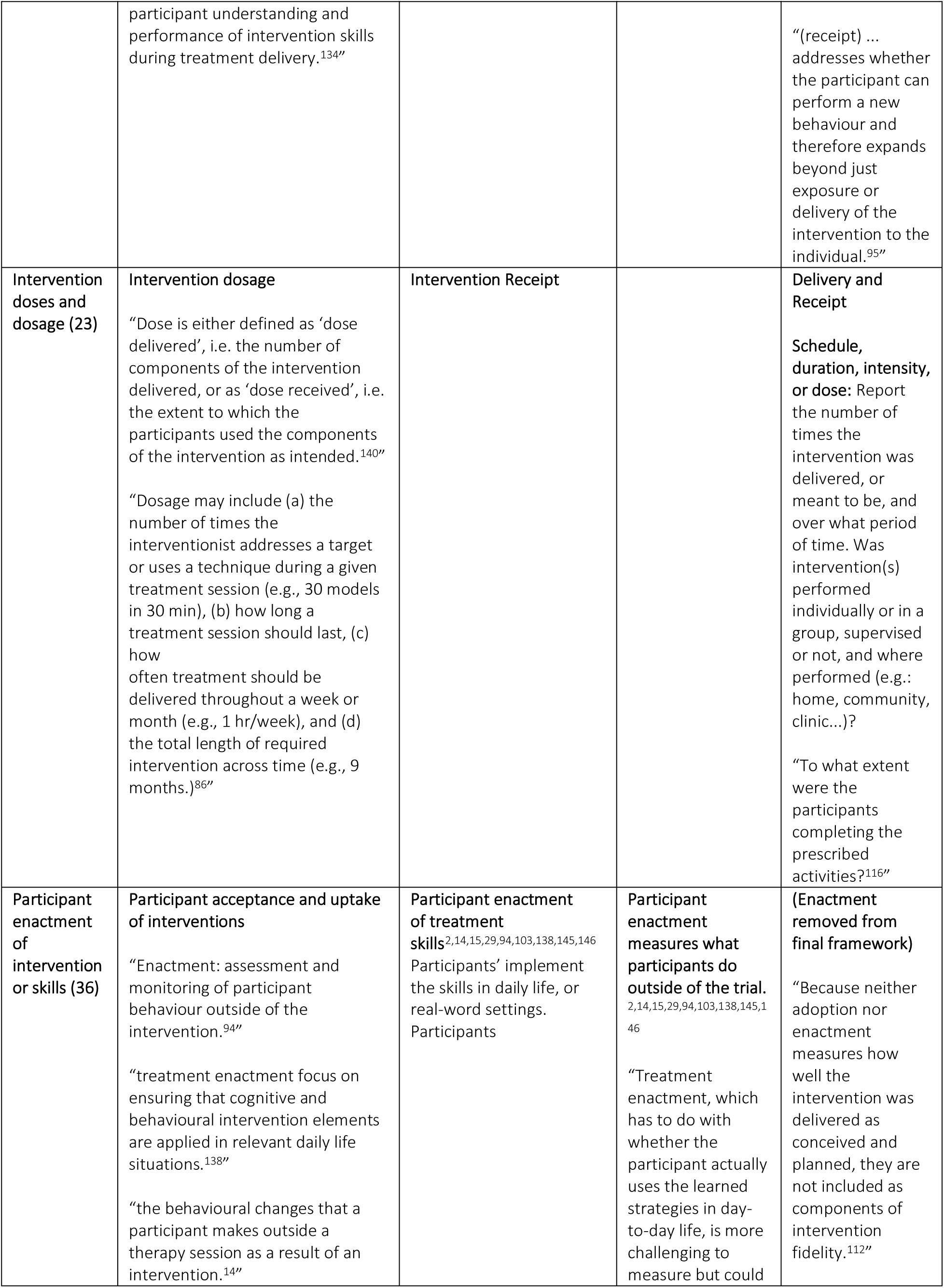
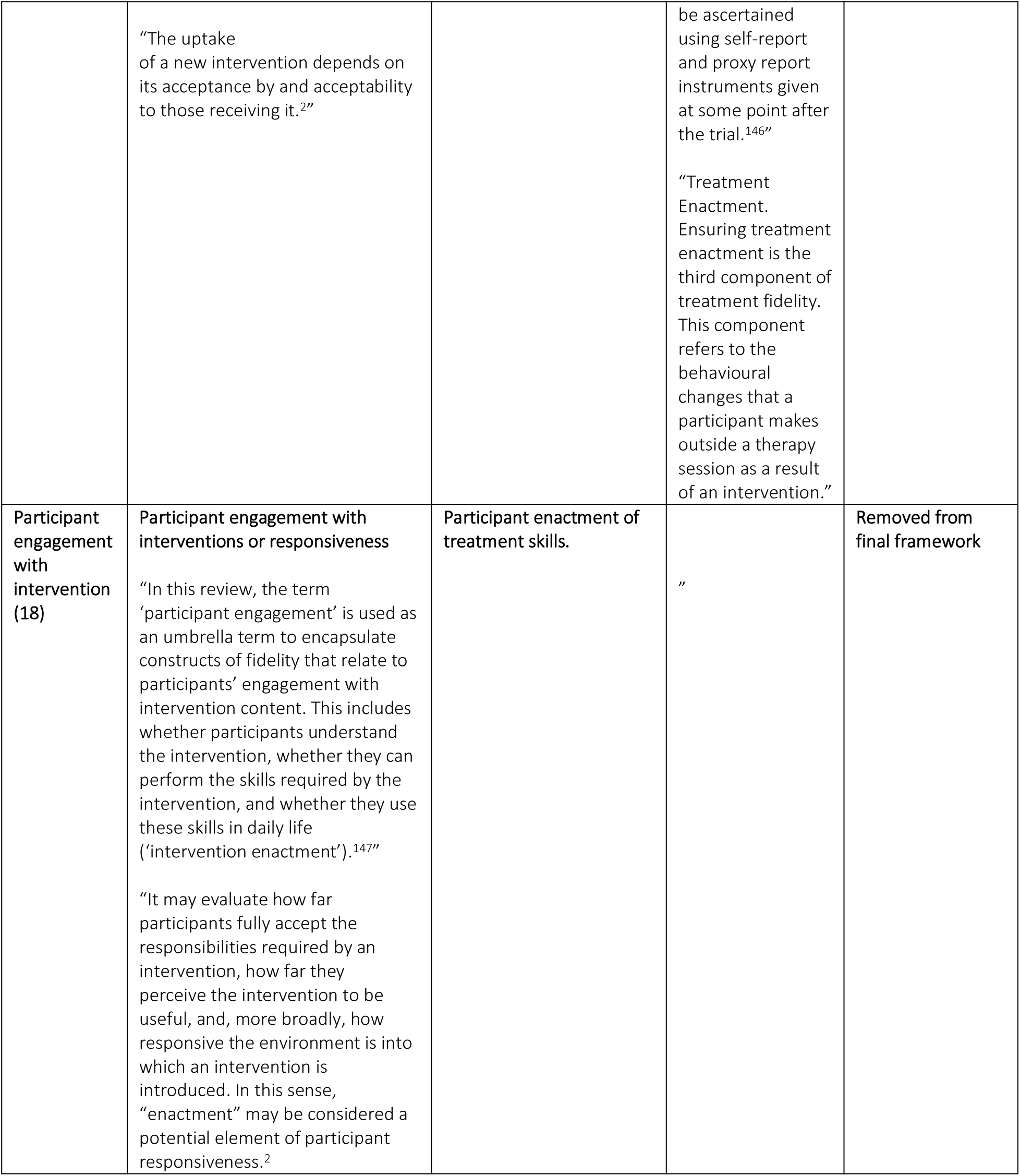
Coding of fidelity data.

### BFFS Step 4: Coding of the data

The first review of fidelity data (quotations) extracted from eligible papers a resulted in 14 fidelity concepts derived from fidelity definitions, descriptions, components, or processes supporting, maintaining, or monitoring fidelity (table 2.4). These were then collapsed through thematic analysis into overarching fidelity themes, supported by exemplars (direct quotations) from included papers, and fed into the BFF framework in the steps 5 and 6.

### BFFS Steps 5 and 6: Thematic analysis and synthesis

Some fidelity themes created in step 4 were collapsed into each other and into an existing BFF category (table 2.4). For example, themes for “intervention design,” “intervention differentiation,” “study design,” “study protocol,” and “development of study manuals or guidance materials” were collapsed into the BFF’s “intervention and study design” theme. This theme includes elements related to the design of both an intervention and the study assessing its effects.

Similarly, themes for “participant engagement with interventions” and “participant enactment of intervention skills” were collapsed into the BFF’s “participants enactment of treatment skills (table 2.4).” Engagement has been described as an umbrella term that includes both skill enactment and intervention acceptability.^52,144,147^ Reviewing quotations supporting both enactment and engagement, enactment was often used synonymously with participant engagement to describe a range of behaviours and perspectives influencing how participants interact with therapeutic interventions.^148^

Secondary thematic analysis and synthesis of the BFF themes and supporting quotations (exemplars) was undertaken to refine the framework further. The “intervention and study design” and “interventionist training” themes were expanded based on exemplars emphasizing the need to include interventionist training strategies and skill monitoring in the design phase of a trial and monitor drift of provider skills during intervention delivery(table 2.4). Similarly, the “tailoring and adaptation” BFF theme was expanded to accommodate differentiation of allowable interventions tailoring or adaptation prespecified in a study protocol from unintended modifications made to interventions during a trial.^75,149^

After secondary thematic analysis and discussion among researcher-reviewers, fidelity data extracted from the included studies supported all but one concept in the best-fit framework. Enactment was removed from the finalised fidelity framework. Enactment was the least frequently addressed component of intervention fidelity in studies assessing the reporting of intervention fidelity in physical complex intervention literature.^84,98,150^ Thirty-one of 36 papers enactment as a component of fidelity described it encompassing participants’ behaviour *outside* of the clinical trial or intervention, differentiating it from receipt, or participants’ use of intervention skills during the intervention itself.^29,81,144,151,152^

A trial participant may receive a treatment delivered with perfect fidelity, and yet be unwilling or unable to apply it in daily life.^2,144^ This may be influenced by a variety of factors not related to the degree of fidelity with which the intervention was delivered during the trial, such as participants forgetting to do it, lacking a suitable setting, not seeing the intervention as being relevant to them, or losing interest in the intervention.^2,14^ Enactment may reflect intervention acceptability, or participants’ affective attitude or responses to the intervention, rather than the fidelity with which it was administered in the trial.^144,153^ Enactment may relate to treatment effectiveness in influencing participants’ behaviour, rather than the fidelity with which treatments were delivered during a trial.^27,29,154,155^

While it is possible to ascertain if participants or their caregivers have understood what an intervention is meant to achieve or can perform intervention activities in the trial (both aspects captured in Receipt), participant enactment during trials may not be measurable if an intervention does not involve participants learning a set of measurable skills.^156,157^ Measurement of enactment may also be impractical due to difficulty defining what it constitutes and how to capture and analyse data for it.^29,152^ It is also unclear how enactment differs from other concepts describing participant behaviours frequently used in rehabilitation literature, such as participant engagement or equipoise.^52,147^ Consequently, treatment enactment was removed from our model of clinical trial intervention fidelity. The remaining fidelity themes formed the new fidelity framework, described in step 7.

### BFFS Steps 7: The CONSIDER Framework

The resulting fidelity framework, the **Co**mplex I**n**terventions De**si**gn, **D**elivery, **R**eceipt (CONSIDER) framework (Figure 2.4), is a multidimensional construct consisting of three main components: Design, Delivery and Receipt. These encompass most of the life cycle of a complex intervention clinical trial, from study design through the clinical trial and process evaluation, with fidelity processes playing a key role in each. Their components were specified and supported with examples and direct quotations from empirical papers identified in the systematic searches in stage 1 and used to create a CONSIDER checklist to facilitate their identification in complex intervention trial papers. The checklist is described later in this chapter. Its reliability was assessed and is described in the following chapter.

**Figure 2.4:**
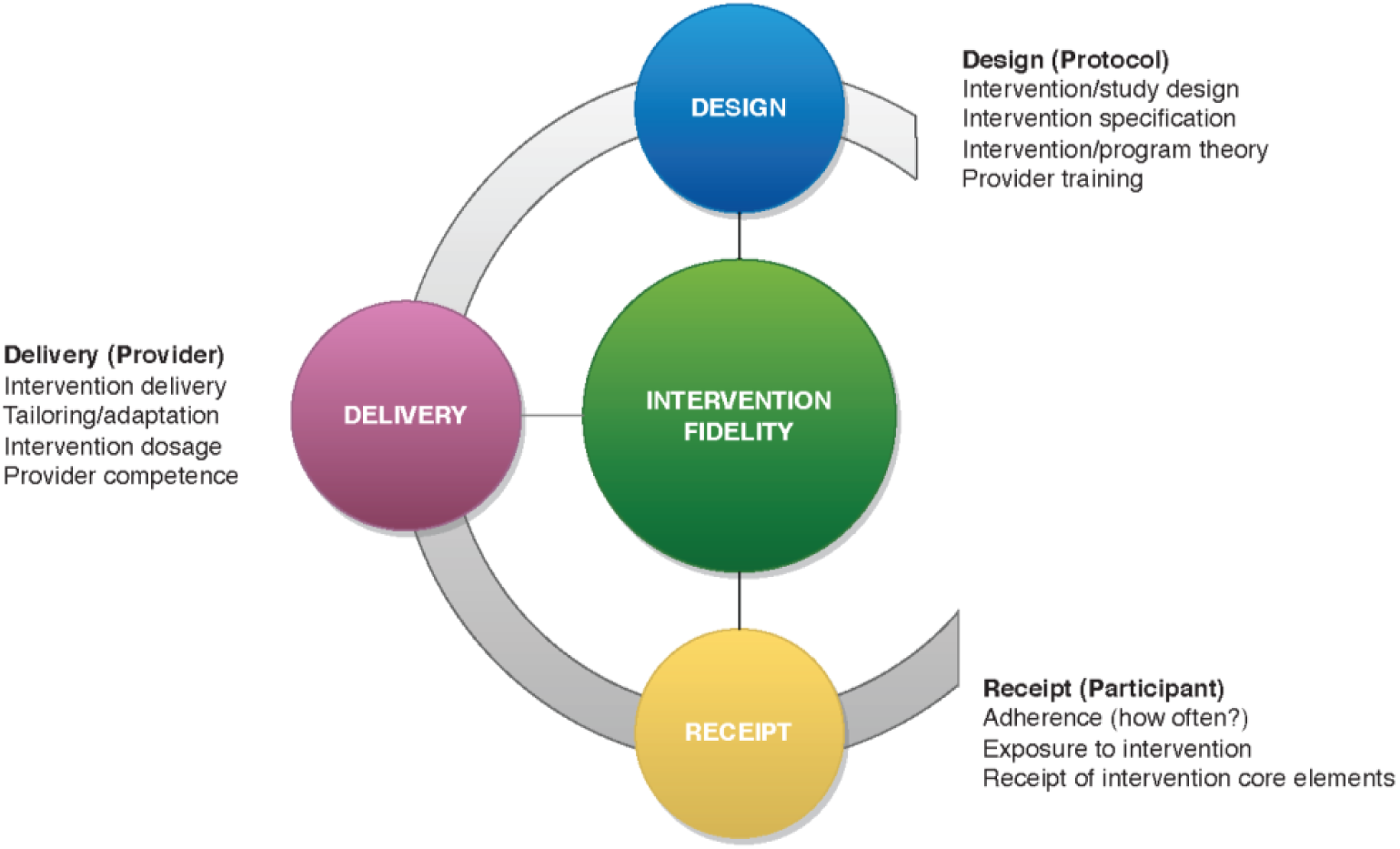
The Consider Framework.

#### The CONSIDER Framework: Design

In the design category, the essential elements of the intervention and fidelity processes in the clinical trial to evaluate it are specified.^2,146,167^ It includes intervention definition, intervention or programme theory, trial design, and the provider training plan for the clinical trial. Fifty-five papers in the thematic synthesis addressed components of intervention and study design and 34 addressed aspects of provider training. These items closely correspond to the TIDieR checklist’s items; 1 (Intervention name or description), 2 “Why” (rationale, theory, or goal of the elements essential to the Intervention), 3 “What” (Materials: physical or informational materials used in the intervention, delivery or in training of intervention providers), 4. (Procedures: activities, and/or processes used in the intervention, including any enabling or support activities, 5 “Who provided” (intervention provider, expertise, and any specific training given), 6 “How” (modes of delivery of the intervention).^35^ They also align with Items 11a–d in the SPRIT checklist, relating to trial protocols providing information about “each group with sufficient detail to allow replication’ and ‘procedures for monitoring adherence to intervention protocols.^34^”

#### Elements of “Design”

**Intervention definition** is the specification of an intervention’s active ingredients and components and forms the foundation for fidelity.^167–170^ Active ingredients are the treatment elements hypothesized to produce intervention effects.^171^ They are the essential components influencing the physiological and behavioural effects that the intervention is designed to deliver (object of treatment).^2,29,146,167,172^ Examples include mechanical force applied by a brace to a joint, lysis of intra-abdominal adhesions during laparoscopy, or learning that modifies a pattern of behaviour. Identifying interventions’ active ingredients is vital for differentiation between the experimental and control interventions, ensuring that they do not overlap or provide the same therapeutic elements, confounding attribution of treatment effects to the investigated intervention.^2,87,107,109,132,146,167,173^

Intervention definition also describes the intervention’s **treatment theory**^174^, or how particular ingredients directly alter specific aspects of functioning and what actions interventionists take to deliver them to influence the object of treatment.^2,29,84,104,146,167,172^ An intervention may be designed to influence the object of treatment directly or to produce effects distal to the object of treatment.^89,175,176^ For example, endovascular thrombectomy (surgical intervention) with mechanical clot removal (active ingredient) may be designed to achieve vascular reperfusion (direct object of treatment). Progressive resistive exercises (active ingredient) may be prescribed to improve muscle strength as a direct object of treatment object, or as a component in a programme to improve stair climbing, a more distal outcome in which the treatment mechanism does not directly act on the outcome. Treatment theory can help ensure that interventions are targeting appropriate outcomes. More extensive use of treatment-theory has been associated with greater effectiveness and statistically significant increases in effect sizes.^177,178^

Treatment theory can help delineate interventions’ key active ingredients when there are several potential ingredients present.^2,29,84,104,146,167,172^ This can facilitate identification of core and flexible components of an intervention and setting of allowable parameters within which tailoring or adaptation can take place while ensuring interventions remaining faithful to their underlying theory of action.^2,29,84,104,146,167,172^ This corresponds with TIDieR^35^ checklist items 9 “Tailoring” (if the intervention is planned to be personalized, titrated or adapted, then describe what, why, when, and how) and CERT^36^ items 14a and 14b (tailoring and adaption, including detailed description of how exercises are tailored to the individual).

Complex interventions often require **tailoring and adaptation** of interventions to individual participants and contexts.^6,7,179^ This requires identification of contextual factors and participant characteristics that may necessitate intervention adaptation.^179^ Some complex interventions, such as surgical procedures, may require adaptation and tailoring that is unforeseeable during protocol planning, and strict adherence to every element of very detailed processes for these interventions may be unsafe or unfeasible. In such cases, it becomes important to determine, *a priori*, which active ingredients or components of the surgical intervention are essential for patient safety as well as intervention integrity and monitor fidelity to those while allowing necessary flexibility for the surgical provider and maintaining essential intervention fidelity.^13,180^ Consideration of the limits of acceptable tailoring and pre-determining allowable parameters for in-trial intervention adaptation are also important when interventions are being evaluated against an active control or standard of care, ensuring that interventions do not deliver the same active ingredients and there is no carryover between groups, maintaining intervention differentiation.^2,29,95,181^

CONSIDER’s Design category also supports key elements of trials to test the effectiveness of an intervention. These include the trial’s **programme theory**, or how interventions should be structured or administered to achieve a therapeutic outcome and goals that define the structure, process, and outcomes of a clinical trial.^182^ Other key elements include best-practice methods for trial protocols and clinical trials following appropriate recommendations such as SCARE^183^, PROCESS^184^, CONSORT^185^, SPIRIT^34^, TIDieR^35^, etc. These include selection of an appropriate study design, experimental and control intervention dosages, and delivery methods, etc. It also supporting the guidance of the Rehabilitation Treatment Specification System (RTSS), ^167,170^ and recommendations for surgical trials developed by Blencowe, et al,^80^ by recommending explicit documentation of interventions’ hypothesized active ingredients, treatment theory and targets of treatment in the design stage and in study protocols.

#### Monitoring and reporting elements of Design

CONSIDER emphasizes pre-determining mechanisms for monitoring and reporting of intervention fidelity and ensuring interventionists can deliver the intervention competently and with high fidelity during the trial.^29,84,98,186,187^ These correspond with: CERT^36^ checklist item 2 (who: qualifications, expertise, or training undertaken by the exercise instructor); 5 (detailed description of how adherence to exercise is measured and reported); items 13 (when and how much, or dosage); and items 16a (describe how adherence or fidelity were measured). Accurate intervention delivery is highly dependent on the skill, experience, and knowledge of the interventionist.^29,87^ Study protocols should include mechanisms to assess for ongoing supervision of intervention delivery throughout the trial to ensure delivery consistency across providers and settings, minimizing provider drift from the protocol over time.^104,118,130,146^

These mechanisms should be determined, *a priori*, and may include interventionist training in the study protocol with well-defined and study procedures,^99,188^ manualisation of the intervention,^15,29,122,123^ review of the manual with interventionists,^104,122^ interventionist supervision, support and audit of delivery during the trial.^29,104,160^ Provider training may also be enhanced by use of case scenarios and group learning experiences to help support different training needs among intervention providers.^189^ Consideration of barriers to successful interventionist training such as intervention complexity, the number of treatment components and the specificity of each should also inform the interventionist training plan.^29,104,181^

Intervention manuals may contain key details about the trial design, procedures, and programme. They often include an overview of the intervention and the intervention theory, detailed descriptions or depictions of intervention activities, equipment and materials needed, mode of delivery, intervention goals and strategies, and the role and responsibilities of interventionists.^29,161^ They may also provide important guidance for allowable tailoring and adaptation of interventions for individual participants and addressing problems that may arise in the intervention.^18,29,52,92^ Review of intervention manuals that describe intervention and study procedures and assessments with detailed written and photographic descriptions, visual aids, exemplars or decision-making aids and can enhance provider training and intervention fidelity.^2,87,104,122,190^

#### Delivery

Delivery encompasses the provision of interventions to participants as specified in the study protocol.^2,29,106^ It focuses mostly on a trial’s independent variables and the actions of the provider. Intervention delivery is the most frequently addressed component of fidelity in complex interventions literature.^27,92^ In thematic synthesis, 60 papers described adherence to protocols, 83 addressed aspects of intervention delivery, 29 papers describing maintenance of interventions’ integrity, 6 describing intervention tailoring and adaption to individual patients or providers, and 37 describing aspects of provider competence and training in the intervention to ensure fidelity to protocols or treatment integrity.

#### Elements of Delivery

Key themes in “Delivery” include maintenance of intervention delivery fidelity, also often referred to as treatment integrity or procedural fidelity (was the intervention delivered as intended?) in included papers; quality of intervention delivery; adherence to treatment protocols, tailoring and adaptation of interventions within prespecified limits; ensuring providers are trained and competent to deliver interventions, and controlling for provider differences.^104^ This category corresponds with TIDieR^35^ checklist items 10 “Modifications” (if modified during delivery, what, why, when, and how), and items 11 and 12 “How well (intervention fidelity and adherence and assessment).^35^ It also corresponds with CERT^36^ item 5 (detailed description of how adherence to exercise is measured and reported), items 16a and 16b (describe how adherence or fidelity were measured, and extent to which the intervention was delivered as planned) and The SPIRIT^34^ item 11c (strategies to improve adherence to intervention protocols, and any procedures for monitoring adherence).

Aspects of Delivery may overlap with, but proceed from, components of Design. For example, adhering to intervention protocols during a trial requires that interventions be operationalized in detail in the Design phase.^29^ **Intervention delivery fidelity** describes whether intervention components and interventionist behaviours are delivered as intended, ensuring the interventions’ delivery of active ingredients.^130,169^ Monitoring of intervention delivery fidelity, preserving interventions’ integrity is essential for maintaining differentiation between interventions during a clinical trial. Differentiation minimizes contamination of the intervention under investigation with elements of other interventions or mixing of active ingredients between the control and experimental interventions.^123,125,129,131,189,191^

Interventionist adherence to study protocols was the most frequently identified component of fidelity of delivery in the thematic analysis, identified in 60 of the 130 papers. While adherence to study protocols supports intervention integrity, ^124,125,127,128^ intervention fidelity may not necessarily require strict adherence to every component of the protocol, as discussed previously in the “Design” section, and may be supported by having prespecified allowable tailoring and adaptation of interventions to individual participants or contexts in the design stage. In highly pragmatic trials, however, it may be necessary to prioritise fidelity to treatment theory or the trial’s programme theory (theoretical fidelity) to maintain fidelity to interventions’ underlying causal processes and reflect real-world clinical practice conditions, rather than fidelity to specific procedures during intervention delivery (content fidelity).^21,182,192^ This is explored with real-time data gathered prospectively in an ongoing pragmatic trial (ACL-SNNAP^193^) in chapter V.

An important theme identified in thematic analysis was the importance of **provider competence** (trained interventionists) for maintaining fidelity during intervention delivery, corresponding to TIDieR^35^ item 5, “who provided (describe their expertise, background and any specific training given) and 3 “what (describe physical or informational materials used in training of intervention providers.” Competence captures practitioners’ skill in delivering the intervention and ability to comply with proscribed behaviours and avoid contaminating the intervention with prohibited components or behaviours. This may be influenced by provider training built into a trial at the design stage and providers’ ability to maintain skill in delivering the intervention throughout the trial.^29,52,194^ The influence of provider training and competence extends beyond intervention delivery, also encompassing non-specific treatment effects such as interventionist ability to engage with participants, warmth, and communication skills.^29,81^

Provider competence during the trial can be supported by ensuring providers are familiar with the intervention manual and can access it as a source of guidance or support, and have supervision to prevent deviation from the intervention protocol or provide guidance when unforeseen modifications are required.^29,109^ Methods designed to enhance and support provider competence should control for provider differences in education, and experience with intervention delivery and monitor whether interventionists maintained the skill set learned in training throughout the clinical trial.^130,139^

#### Monitoring of elements of Delivery

Delivery can be monitored by video recording and assessment of patient sessions with a fidelity checklist to ensure the intervention is delivered as specified in the protocol and intervention integrity is maintained.^106^ Other assessment options include assessment of randomly sampled audio or video recorded treatment sessions,^195^ observation of treatment sessions,^196,197^ interviewing of participants,^198,199^ provider self-assessments,^85,155^ review of provider treatment notes or adherence logbooks^200,201^ with comparison to the protocol or intervention manual,^29,87,202^ and process evaluation to assess protocol adherence and treatment integrity.^101,106^ Many of these actions can also facilitate monitoring provider competence and maintenance of skills learned in training throughout the clinical trial.^130,139^

#### Receipt

While Delivery focuses mostly on the actions of intervention providers, Receipt mostly focuses on the actions of the intervention recipients.^2,15,98,203^ It was most often referred to as participant adherence in the eligible papers^2,4,29,140,204,205^ and partially corresponds to CERT^36^ item 13, SPIRIT^34^ item 11c and TIDieR^35^ item 8-“when and how much (number of times the intervention was delivered, when, how much, intensity and dosage) and 11 “how well.”

#### Elements of Receipt

Intervention Receipt was identified in 42 papers as participants’ exposure to the intervention or its active ingredients (dose), their adherence to the frequency of the intervention or attendance in interventions sessions or appointments (adherence), degree to which they followed clinic and/or home-based components of the treatment, understanding of intervention skills, and ability to perform intervention-related skills during treatment in the trial.^28,52,206,81,95,107,130,134,146,165,194^ In complex interventions literature, less focus has been placed on monitoring and reporting of intervention receipt than intervention delivery, interventionist training, or other aspects of fidelity.^28,29,84,152^ Nevertheless, key components of receipt such as intervention dose, participant session attendance, comprehension and performance of intervention related skills can greatly influence intervention fidelity and intervention outcomes.^28,52^

Participants’ **exposure to interventions** and their active ingredients can be represented by participants’ acceptance and initiation of their allocation intervention (**participant adherence**), the frequency and intensity with which interventions are delivered (dose); the degree of participants’ attendance in treatment sessions and performance of intervention activities (adherence).^207^

**Intervention dose** can be further classified as either the intervention dose delivered-the number, frequency or intensity with which intervention components are delivered by interventionists, or as the intervention ‘dose received’, or the extent to which the participants performed the components of the intervention or attended intervention sessions as intended.^28,52,81,86,95,130,134,165^ Measures of the number of treatment sessions or units of an active ingredient participants received can be used to indicate if a treatment met its prescribed dose.^73,208^ For example, dosage may be measured by the number of times the interventionist addresses a target or uses a technique during a given treatment session, the number of times or duration with which a participant achieves a desired physiological state (e.g. amount of time spent exercising at a desired percentage of maximal heart rate during the intervention), the number of treatment sessions a participant attended or number of time a participant performed an intervention activity (for example, twice weekly over six weeks).^73,86,143,208,94,121,134,138–142^ Dose can be monitored with instruments measuring participants’ exposure to the intervention, such as interventionist or participant logs, intervention notes, checklists, or attendance records.^18^

Receipt also includes ensuring participants’ ability to perform intervention skills during the trial.^73,86,143,208,94,121,134,138–142^ This assesses not just whether (or how much) participants performed intervention activities, but also how well they did so.^209^ Participants’ ability to perform intervention related skills during the trial is important for supporting their exposure to the interventions’ active ingredients. This is particularly important for maintaining fidelity in interventions relying on participant-generated movement, such as physiotherapeutic exercise or rehabilitation interventions.^130^ Participants’ ability to perform intervention-related skills may also be influenced by moderating factors such as intervention complexity and interventionists’ skill in communicating with participants.^2,29,81,181^ CONSIDER’s emphasizes manualizing intervention components, and ensuring provider competence in delivering interventions, supporting Receipt.

#### Monitoring Receipt

Receipt has been operationalized and monitored in a variety of ways in complex interventions literature.^28^ These include assessment of records from intervention sessions or treatment logs, participant attendance logs, participant-completed checklists or activity logs, field notes, website monitoring or monitoring of competition of online intervention modules, and qualitative interviews with participants. Other examples included participants being contacted by trialists or receive informational material, DVDs weblinks, emails, texts or other contacts and resources to ensure their understanding of the intervention instructions and enhance intervention receipt. Assessments of participant receipt based on attendance logs, treatment session notes, field notes, daily journals, completion of practice logs, logins/website monitoring, were generally collected during the intervention period.

#### Moderating factors for intervention fidelity

The Medical Research Council’s (MRC) guidance on process evaluations describes the term *context* as including, “anything external to the intervention that may act as a barrier or facilitator to its implementation, or its effects.^210^” Several such potential moderating factors for intervention fidelity were identified during the BFFS and may also need consideration when monitoring fidelity in clinical trials.^2,29^ Factors outside of the intervention, such as scheduling and difficulty accessing the intervention site may influence participant receipt and engagement with interventions. Comorbid conditions reducing participants’ ability to perform the intervention, or participation affected by poor interactions with the intervention or interventionist, may also reduce both intervention delivery and intervention receipt in a trial.^2,29^ Providers’ prior expertise with an intervention and can also influence participants’ receipt and engagement and should be considered when evaluating factors influencing intervention effectiveness.^139^ The acceptability of interventions to providers and provider or participant equipoise may also influence their delivery of the intervention and participants’ receipt.^143,186,207,211^ While these and other factors external to the intervention lie somewhat outside of the core aims of this framework synthesis, they should also be considered as part of a trial’s intervention implementation plan or process evaluation.^2,206^

## OBJECTIVE 3: Definitions of Fidelity

Ninety-five descriptions or definitions of intervention fidelity were identified in the systematic review’s eligible papers. Multiple terms, such as: adherence, integrity, compliance, concordance, fidelity, or specification were used, often interchangeably, to describe concepts or processes related to intervention fidelity. Researchers conceptualised or discussed fidelity in terms of interventionists’ adherence to a study protocol or treatment manual; the extent to which the intervention delivered resembled the intervention that was intended; the extent to which intervention was delivered as planned^212^; protocol adherence and acceptability; adherence and provider competence^213^; and, methodological practices used to ensure that a research study reliably and validly tests a clinical intervention (table 2.5).^88^

**Table 2.5:**
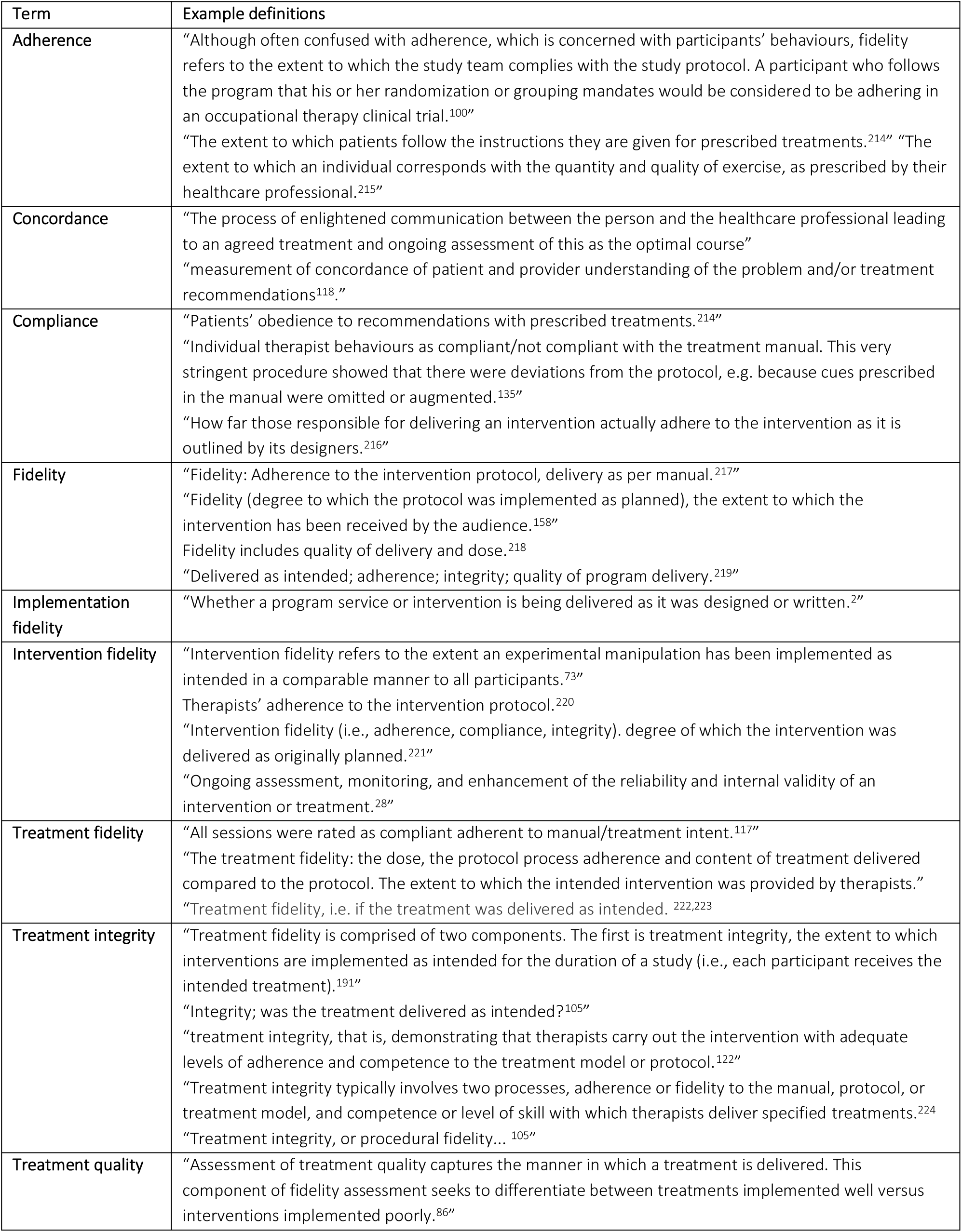
Terms and definitions for fidelity.

| Term | Example definitions |
| --- | --- |
| **Adherence** | "Although often confused with adherence, which is concerned with participants' behaviours, fidelity refers to the extent to which the study team complies with the study protocol. A participant who follows the program that his or her randomization or grouping mandates would be considered to be adhering in an occupational therapy clinical trial.<sup>100</sup>"<br>"The extent to which patients follow the instructions they are given for prescribed treatments.<sup>214</sup>" "The extent to which an individual corresponds with the quantity and quality of exercise, as prescribed by their healthcare professional.<sup>215</sup>" |
| **Concordance** | "The process of enlightened communication between the person and the healthcare professional leading to an agreed treatment and ongoing assessment of this as the optimal course"<br>"measurement of concordance of patient and provider understanding of the problem and/or treatment recommendations<sup>118</sup>." |
| **Compliance** | "Patients' obedience to recommendations with prescribed treatments.<sup>214</sup>"<br>"Individual therapist behaviours as compliant/not compliant with the treatment manual. This very stringent procedure showed that there were deviations from the protocol, e.g. because cues prescribed in the manual were omitted or augmented.<sup>135</sup>"<br>"How far those responsible for delivering an intervention actually adhere to the intervention as it is outlined by its designers.<sup>216</sup>" |
| **Fidelity** | "Fidelity: Adherence to the intervention protocol, delivery as per manual.<sup>217</sup>"<br>"Fidelity (degree to which the protocol was implemented as planned), the extent to which the intervention has been received by the audience.<sup>158</sup>"<br>Fidelity includes quality of delivery and dose.<sup>218</sup><br>"Delivered as intended; adherence; integrity; quality of program delivery.<sup>219</sup>" |
| **Implementation fidelity** | "Whether a program service or intervention is being delivered as it was designed or written.<sup>2</sup>" |
| **Intervention fidelity** | "Intervention fidelity refers to the extent an experimental manipulation has been implemented as intended in a comparable manner to all participants.<sup>73</sup>"<br>Therapists' adherence to the intervention protocol.<sup>220</sup><br>"Intervention fidelity (i.e., adherence, compliance, integrity). degree of which the intervention was delivered as originally planned.<sup>221</sup>"<br>"Ongoing assessment, monitoring, and enhancement of the reliability and internal validity of an intervention or treatment.<sup>28</sup>" |
| **Treatment fidelity** | "All sessions were rated as compliant adherent to manual/treatment intent.<sup>117</sup>"<br>"The treatment fidelity: the dose, the protocol process adherence and content of treatment delivered compared to the protocol. The extent to which the intended intervention was provided by therapists."<br>"Treatment fidelity, i.e. if the treatment was delivered as intended. <sup>222,223</sup> |
| **Treatment integrity** | "Treatment fidelity is comprised of two components. The first is treatment integrity, the extent to which interventions are implemented as intended for the duration of a study (i.e., each participant receives the intended treatment).<sup>191</sup>"<br>"Integrity; was the treatment delivered as intended?<sup>105</sup>"<br>"treatment integrity, that is, demonstrating that therapists carry out the intervention with adequate levels of adherence and competence to the treatment model or protocol.<sup>122</sup>"<br>"Treatment integrity typically involves two processes, adherence or fidelity to the manual, protocol, or treatment model, and competence or level of skill with which therapists deliver specified treatments.<sup>224</sup><br>"Treatment integrity, or procedural fidelity... <sup>105</sup>" |
| **Treatment quality** | "Assessment of treatment quality captures the manner in which a treatment is delivered. This component of fidelity assessment seeks to differentiate between treatments implemented well versus interventions implemented poorly.<sup>86</sup>" |

The words most frequently used when describing or defining fidelity were fidelity (n = 260), delivery (131), training (97) adherence (93), compliance (86) protocol (84), delivered (84) intended (71) and receipt (67) (Figure 2.5).

**Figure 2.5:**
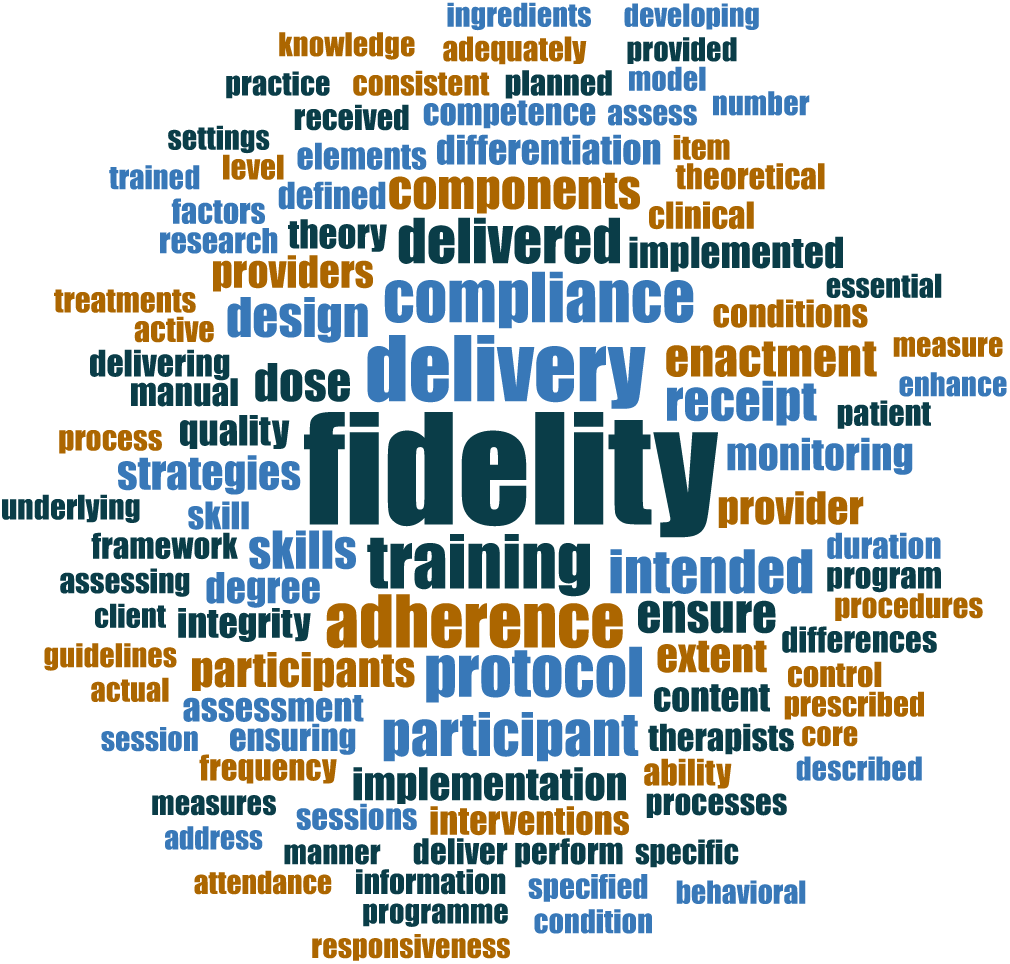
Fidelity terms word cloud.

### Fidelity and adherence

Within these definitions and descriptions, two distinct concepts emerged: Fidelity and adherence. **Fidelity** (intervention, treatment, or implementation) most often referred to the action of interventionists and the quality of their intervention delivery during the trial. Operational constructs defined fidelity in procedural terms related to the administration of a therapeutic intervention, including the integrity of treatment delivery, or the closeness or concordance of the intervention delivered to the intervention intended in the trial protocol or manual. Definitions focused on construct fidelity referred to the extent to which interventions delivered in the trial were faithful to their underlying theoretical basis, active ingredients or clinical guidelines.^173^

The other concept, **adherence**, most often referred to action of participants, or the extent to which participants complied (compliance) with, performed, were exposed to or received the intervention in the trial.^100^ For example, adherence was exemplified by participants following their randomised group allocation and not crossing over to the comparison condition in surgical or rehabilitation trials,^2,4,34,78^ and attending the prescribed number or therapy sessions or fully performing the required number of home exercises or activities in rehabilitation trials.^2,4^ Participants who are either underexposed to the intervention because they did not attend intervention sessions or failed to perform intervention activities, or were overexposed to the intervention because they received greater intervention dosage or greater number of intervention sessions than prescribed in the study protocol would be considered non-adherent.^100^

### Integrated definition of intervention fidelity

Through thematic analysis, we also derived an integrated definition of intervention fidelity in which intervention fidelity is an umbrella concept encompassing two distinct but related and interacting components: intervention fidelity and participant adherence. Both are essential for a clinical trial to be faithful to its protocol, and both can influence study outcomes, individually or together:^3,225–227^

1. **Intervention delivery fidelity (fidelity)** focuses mainly on the actions of the interventionist. It encompasses the quality of intervention delivery or performance within the trial, and reflects the correspondence of interventions delivered in the trial with the intervention specified in the study protocol, or in accordance with study procedures, treatment manuals, etc.
2. **Participant adherence (adherence)** focuses mainly on the actions of the participant. Adherence encompasses both whether participants accept and initiate the intervention allocated, and how well they comply with the prescribed, allocated intervention. For example, this could be represented by measuring whether participants attended the required number of therapy sessions, the frequency of intervention sessions or frequency participants performed intervention activities (for example, completed a home exercise a certain number of times per week). This definition also parallels adherence as defined in pharmaceutical trials.^228^

## DISCUSSION

Intervention fidelity is an essential part of conducting intervention research and implementing the findings into clinical practice. ^13,18,41,92^ A lack of a uniform definition of fidelity and its key components has been identified as a barrier to fidelity planning and intervention implementation in clinical trials and their translation to clinical practice.^29,229^ The broad range of fidelity terms, definitions and concepts used in complex interventions literature also makes it difficult to systematically identify fidelity reporting in clinical trials. The CONSIDER framework synthesizes key aspects of intervention fidelity from 269 empirical and methodological papers to create a fidelity framework developed specifically for clinical trials of physical complex interventions.

CONSIDER was developed as a basis from which to frame and investigate intervention fidelity in rehabilitation and similar complex interventions in healthcare, but also represents an important first step in providing practical guidance for intervention fidelity in the planning and implementation of clinical trials in domains involving physical complex interventions such as physiotherapy and rehabilitation. To the best of our knowledge, it is the first “Best-fit” framework synthesis of intervention fidelity and first empirically based fidelity framework created specifically for complex interventions in the physical domain. Further development of the framework and checklist with broader input from a wider range of stakeholders is needed to refine the framework and enhance its applicability for future evaluations of complex intervention clinical trials.

Much of the fidelity monitoring identified in this framework synthesis and in previous systematic reviews focuses on intervention components adhering to trial protocols or participant adherence to treatment frequencies.^28,29,84,152^ While these are important elements of intervention fidelity, focusing on these alone neglects the influence of other key elements of fidelity on the outcomes of a clinical trial. ^2,80,194^ For example, poor treatment specification or interventionist training in the Design stage may lead to suboptimal intervention delivery, which may lead to poor participant exposure to the intervention (Receipt). CONSIDER offers a more complete conceptualization of fidelity, encompassing both the interventions and the design and conduct of trials to assess their effectiveness.

CONSIDER also supports intervention fidelity that is flexible, recognizing that tailoring and adaptation of interventions may be necessary to accommodate individual participants and clinical contexts. ^230,231^ Rather than rigid adherence to large numbers of intervention components, CONSIDER emphasises tailoring and adaptation within pre-determined boundaries that is based on fidelity to interventions’ treatment theory, retaining intervention fidelity while not impeding the application and effectiveness of complex interventions.^2,80,194^

The CONSIDER framework can be used in conjunction with existing clinical trial models or frameworks to contribute a deeper, broader conceptualization of intervention fidelity. It complements other, established design and reporting frameworks such as CONSORT, TIDieR^35^ or CERT^36^. Enhancing fidelity in the design and intervention implementation of clinical trials supports enactment of processes reported on TIDieR^35^, CERT^36^and SPIRIT^34^. While Intervention fidelity is a separate concept from intervention reporting, an important relationship exists between the two. Intervention fidelity cannot be reported adequately if it has not been previously considered or monitored. The processes which support intervention fidelity also support transparency and enhance the documentation of intervention details needed to support reproducibility and the dissemination of evidence-based methods.^13,30,92,167,170,186^ Variable and imprecise description of intervention components in clinical trial papers makes it difficult to identify the active ingredients interventions were meant to deliver or whether departures from the intended intervention took place.^1,35,39,105,167,232,233^ CONSIDER complements the recommendations of the Rehabilitation Treatment Specification System (RTSS), emphasizing identification of interventions’ active ingredients and treatment theory and the development of empirically testable interventions.^167,170^

The recommendations of the CONSIDER framework can also support the development of better comparison-control treatments for complex interventions, complementing the DITTO (**D**econstruct, **I**dentify, **T**ake out, **T**hink risk, **O**ptimise) framework^234^ and ASPIRE guidelines for placebo and sham intervention controls in surgical and rehabilitation trials.^79^ Placebo controls or sham interventions appear similar to the experimental treatments but lack their active ingredients,^235–238^ and minimize the risk of biases such as expectation, performance, detection and confirmation biases.^17,79,235–240^ These biases are high in trials of surgical and physiotherapy interventions and weaken the validity of studies’ findings, but placebo interventions are methodologically difficult to construct and present to patients.^79^

Interventions’ active ingredients and fidelity, as specified in CONSIDER, can be manipulated to move them from the experimental intervention to a placebo intervention delivering no or very low dose of the active ingredients.^241^ Once the intervention being evaluated has been operationalized, its placebo control can be constructed by “moving the needle” between varying levels of intervention fidelity to produce placebo interventions that are identical to the experimental surgical procedure but lack its active or essential components. For example, in the Can Shoulder Arthroscopy Work? (CSAW) trial, a RCT assessing the clinical and cost-effectiveness of arthroscopic subacromial decompression for shoulder pain, the essential surgical element (bone and soft tissue removal) was manipulated to randomise participants to an arthroscopic surgery group with the essential surgical element (active arm), a diagnostic arthroscopy only without the essential surgical element (no spur removal) placebo arm) or an active monitoring group.^242^

No similar guidance exists for the construction of placebo interventions in Physiotherapy or Physical and Rehabilitation Medicine (PRM), encompassing a larger spectrum of medicine and rehabilitation disciplines.^239^ The intensive provider-participant contact and multi-modal nature of physical therapy and PRM interventions present unique challenges for the construction of placebo-controlled trials.^239^ The International Placebo Symposium Working Group was convened in 2010 to address these and made a number of recommendations that would be supported by use of CONSIDER, including greater efforts to reduce variability in intervention implementation, greater evaluation of the isolated components of rehabilitation interventions, and use of structural equivalence, in which the experimental and placebo groups have similar degree of therapeutic contact.^239^

### Strengths, limitations, and future directions

CONSIDER has been developed to encompass the unique challenges and opportunities posed by interventions and clinical trials in domains involving physical complex interventions such as physiotherapy and rehabilitation. We followed a thorough, systematic best-fit framework approach and derived evidence from empirical, methodological, and theoretical literature in these complex interventions, supporting its applicability in their clinical trials. A reported limitation of many existing implementation frameworks and models is that they describe determinants and moderators of fidelity without elaborating on the relationships between them or the mechanisms linking them to implementation outcomes.^243,244^ Our best-fit framework synthesis sought to overcome this limitation though the secondary thematic analysis and reciprocal translation of themes derived from eligible papers, extensive use of exemplars from the complex interventions literature base, and linking of concepts between and across CONSIDER stages.

Our search strategy was comprehensive and maximized sensitivity rather than precision. It was unrestricted by language and included both commercial and grey literature sources.^54^ A broad range of search terms to ensure relevant papers were captured.^54^ Although the search strategies used in this analysis were comprehensive and conducted in multiple search engines, it is possible that some papers describing intervention fidelity may have been missed. The lack of consensus on definitions and components of fidelity, the many terms used to describe it, and poor reporting of fidelity in complex intervention literature increase the risk that some eligible papers may not have been captured by our search terms. However, we employed citation searching and extensive full-text screening to ensure that papers describing intervention fidelity with unanticipated terms were also captured.

Systematic reviews of complex interventions have found poor or completely absent reporting of fidelity monitoring or assessment across clinical trials.^41,80,118,245,246^ This may reflect some degree of editorial constraint, in which word count limits and manuscript length restrictions limit reporting of some aspects of the conduct of clinical trials.^246,247^ We attempted to overcome this by rigorous full-text screening of all papers for concepts or processes related to fidelity and searched for trials’ protocols or registrations, reviewing them and searching for information about intervention fidelity when they were available. The framework synthesis undertaken in this review also aimed for conceptual saturation and generalizability, rather than statistical power ^55,56^, and study selection was purposive rather than exhaustive. The large number of papers in our best-fit framework synthesis maximized the likelihood that conceptual saturation was reached.

Even with our comprehensive searches, we found few reports of trials assessing intervention fidelity in surgical interventions, despite having worked with surgical trialists to enhance the search strategy’s sensitivity and extensive efforts to identify application of fidelity principles in surgical trials. It is possible that some surgical trials may have been missed because the processes that support fidelity during clinical trials were described in terms falling outside of our search strategies. To overcome this, full text screening was undertaken for any surgical papers identified with our search strategies or citation searches to identify papers applying any fidelity principles, even if not labelled as such.

Nevertheless, previous systematic reviews and methodological papers have also identified a paucity of surgical trials monitoring fidelity principles. Beard et al. (2020) reviewed 96 papers describing surgical placebo controlled trials in the development of the ASPIRE guidelines, finding only four papers reporting elements of fidelity and seven reporting standardization of the intervention, a component of intervention fidelity for clinical trials in CONSIDER.^79^ Methodological papers have also identified the unique challenges to intervention fidelity and adherence posed by surgical trials, including inherent and unpredictable variability in surgical procedures due to surgical findings, surgeon learning curve effects, and high potential for cross-over (poor adherence) between trial arms in trials comparing operative versus nonoperative therapy.^78^ As a result of the poor representation of surgical papers in the BFFS, the CONSIDER framework represents the perspective of rehabilitation (i.e. physiotherapy, occupational therapy, speech-language therapy, exercise interventions, etc.) and not of surgery.

Finally, although the “best-fit” framework syntheses method is particularly suited for developing a comprehensive framework based on existing evidence, and our database of empirical and methodological papers was extensive, CONSIDER and the checklist are initial steps that need to be developed further with broader input from a wider range of stakeholders before they can be presented as a tools trialists should be using. In future stages of their development, a Delphi process will be needed to build consensus about the synthesized definition of fidelity, which fidelity components and qualities are most important, and which qualities should be given the most weighting when developing and evaluating intervention fidelity in complex intervention trials.

Additionally, the applicability of the framework and checklist to study settings challenging for fidelity needs to be explored. Explanatory randomised trials are conducted under idealised conditions to give interventions the best chance to demonstrate an effect (efficacy).^248,249^ These tightly controlled conditions can facilitate maintenance and monitoring of intervention fidelity. However, pragmatic randomised clinical trials are designed to evaluate the relative effectiveness of interventions under real-life^250^ conditions, with diverse clinical populations,^251^ and against usual care interventions.^252,253^ Key aspects of intervention delivery may be less tightly controlled in pragmatic trials, creating challenges for assessing and maintaining intervention fidelity.^233,251,254^ The applicability of the CONSIDER framework was investigated in an ongoing pragmatic trial of surgical versus rehabilitation management, the Anterior Cruciate Ligament (ACL) Surgery Necessity in Non Acute Patients (ACL SNNAP)^193^ trial in Chapter V.

## CONCLUSION

This framework synthesis represents an important first step in addressing a gap in our understanding of intervention fidelity in complex interventions in the physical domain. While growing attention has been paid to fidelity when interventions are translated and implemented in clinical practice, far less research has focused on intervention fidelity during the clinical trials themselves. Guidance specifically tailored to the planning and implementation of intervention fidelity in clinical trials of these complex intervention is rare, and fidelity frameworks developed for psychology and public health trials do not translate well to physiotherapy and surgery. Failure to implement interventions with a high degree of fidelity could negatively affect the accuracy and validity of clinical trials, undermining patient care and the translation of evidence-based interventions into clinical practice.^13,29,92,255^

The CONSIDER framework offers guidance for intervention fidelity in the planning and implementation of clinical trials, with implications for reproducibility and the translation of evidence-based interventions to clinical practice.^251,256–258^ Further development of the CONSIDER framework with broader input from a wider range of stakeholders is needed. Ultimately, the framework may help researchers design clinical trials that enable research reproducibility and uptake, reducing waste and benefiting the practice and evidencing of complex interventions in rehabilitation.

## Data Availability

All data produced in the present study are available upon reasonable request to the authors.

