## appendix for "The CONSIDER Intervention Fidelity Framework for Complex Interventions in Healthcare: A “Best-Fit” Framework Synthesis"

Appendix I: supplementary materials

Figure 1.1 Search strategies: Pubmed

((((((((((((((((((((((("framework") OR "model") OR "procedure") OR "assessment") OR "process") OR "monitoring") OR "monitored") OR "strategy") OR "indicators")) AND (((((((((((("treatment integrity") OR "treatment fidelity") OR "treatment adherence") OR "treatment implementation") OR "treatment delivery") OR "treatment enactment") OR "intervention fidelity") OR "intervention integrity") OR "intervention adherence") OR "intervention implementation") OR "intervention delivery") OR "implementation strategy" AND ( "2006/01/01"[PDat] : "2019/12/31"[PDat] )))))) AND "physiotherapy")) OR (((((((((((((((("framework") OR "model") OR "procedure") OR "assessment") OR "process") OR "monitoring") OR "monitored") OR "strategy") OR "indicators")) AND (((((((((((("treatment integrity") OR "treatment fidelity") OR "treatment adherence") OR "treatment implementation") OR "treatment delivery") OR "treatment enactment") OR "intervention fidelity") OR "intervention integrity") OR "intervention adherence") OR "intervention implementation") OR "intervention delivery") OR "implementation strategy" AND ( "2006/01/01"[PDat] : "2019/12/31"[PDat] )))))) AND "physical therapy")) OR (((((((((((((((("framework") OR "model") OR "procedure") OR "assessment") OR "process") OR "monitoring") OR "monitored") OR "strategy") OR "indicators")) AND (((((((((((("treatment integrity") OR "treatment fidelity") OR "treatment adherence") OR "treatment implementation") OR "treatment delivery") OR "treatment enactment") OR "intervention fidelity") OR "intervention integrity") OR "intervention adherence") OR "intervention implementation") OR "intervention delivery") OR "implementation strategy" AND ( "2006/01/01"[PDat] : "2019/12/31"[PDat] )))))) AND "rehabilitation")) OR (((((((((((((((("framework") OR "model") OR "procedure") OR "assessment") OR "process") OR "monitoring") OR "monitored") OR "strategy") OR "indicators")) AND (((((((((((("treatment integrity") OR "treatment fidelity") OR "treatment adherence") OR "treatment implementation") OR "treatment delivery") OR "treatment enactment") OR "intervention fidelity") OR "intervention integrity") OR "intervention adherence") OR "intervention implementation") OR "intervention delivery") OR "implementation strategy" AND ( "2006/01/01"[PDat] : "2019/12/31"[PDat] )))))) AND "surgery")) OR (((((((((((((((("framework") OR "model") OR "procedure") OR "assessment") OR "process") OR "monitoring") OR "monitored") OR "strategy") OR "indicators")) AND (((((((((((("treatment integrity") OR "treatment fidelity") OR "treatment adherence") OR "treatment implementation") OR "treatment delivery") OR "treatment enactment") OR "intervention fidelity") OR "intervention integrity") OR "intervention adherence") OR "intervention implementation") OR "intervention delivery") OR "implementation strategy" AND ( "2006/01/01"[PDat] : "2019/12/31"[PDat] )))))) AND "occupational therapy")) OR (((((((((((((((("framework") OR "model") OR "procedure") OR "assessment") OR "process") OR "monitoring") OR "monitored") OR "strategy") OR "indicators")) AND (((((((((((("treatment integrity") OR "treatment fidelity") OR "treatment adherence") OR "treatment implementation") OR "treatment delivery") OR "treatment enactment") OR "intervention fidelity") OR "intervention integrity") OR "intervention adherence") OR "intervention implementation") OR "intervention delivery") OR "implementation strategy" AND ( "2006/01/01"[PDat] : "2019/12/31"[PDat] )))))) AND speech therapy)) OR (((((((((((((((("framework") OR "model") OR "procedure") OR "assessment") OR "process") OR "monitoring") OR "monitored") OR "strategy") OR "indicators")) AND ((((((((((("treatment integrity") OR "treatment fidelity") OR "treatment adherence") OR "treatment implementation") OR "treatment delivery") OR "treatment enactment") OR "intervention fidelity") OR "intervention integrity") OR "intervention adherence") OR "intervention implementation") OR "intervention delivery") OR "implementation strategy" AND ( "2006/01/01"[PDat] : "2019/12/31"[PDat] )))))) AND physiatry)

Figure 1.2 Search strategies: SCOPUS

( ALL ( "treatment fidelity" OR "treatment integrity" OR "Treatment implementation" OR "treatment enactment" OR "intervention fidelity" OR "intervention integrity" OR "intervention implementation" OR "intervention enactment" OR “fidelity” OR “compliance” OR “concordance” or “adherence” OR “agreement” OR “delivery” OR “manualized” OR “treatment manual” OR “intervention manual” OR “per protocol” ) AND ALL (therapy OR process OR monitoring OR monitored OR framework OR assessment OR procedure OR strategy OR model OR indicator ) AND TITLE-ABS-KEY ( "physical therapy" OR physiotherapy OR surgery OR physiatry OR "occupational therapy" OR "speech therapy" OR rehabilitation ) ) AND PUBYEAR > 2004 AND ( EXCLUDE ( SUBJAREA , "PSYC" ) OR EXCLUDE ( SUBJAREA , "SOCI" ) OR EXCLUDE ( SUBJAREA , "BIOC" ) OR EXCLUDE ( SUBJAREA , "ARTS" ) OR EXCLUDE ( SUBJAREA , "PHAR" ) OR EXCLUDE ( SUBJAREA , "ENGI" ) OR EXCLUDE ( SUBJAREA , "AGRI" ) OR EXCLUDE ( SUBJAREA , "ENVI" ) OR EXCLUDE ( SUBJAREA , "PHYS" ) OR EXCLUDE ( SUBJAREA , "BUSI" ) OR EXCLUDE ( SUBJAREA , "CENG" ) OR EXCLUDE ( SUBJAREA , "DENT" ) OR EXCLUDE ( SUBJAREA , "EART" ) OR EXCLUDE ( SUBJAREA , "IMMU" ) OR EXCLUDE ( SUBJAREA , "ECON" ) )

Figure 2.1 Search strategies: **Embase**

| Set# | Searched for | Results |
|---|---|---|
| S39 | (S30 OR S31 OR S32 OR S33 OR S36 OR S37) and (pd(20050101-20191205)) | 721° |
| S38 | S30 OR S31 OR S32 OR S33 OR S36 OR S37 | 758° |
| S37 | S27 AND physiotherapy | 89° |
| S36 | S27 AND (speech therapy) | 46° |
| S35 | S27 AND ("speech therapy") | 11° |
| S34 | S27 AND physiatry | 0° |
| S33 | S27 AND ("occupational therapy") | 46° |
| S32 | S27 AND surgery | 399° |
| S31 | S27 AND rehabilitation | 278° |
| S30 | S29 OR S28 | 212° |
| S29 | S27 AND (physical therapy) | 212° |
| S28 | S27 AND ("physical therapy") | 51° |
| S27 | S26 AND S5 | 2437° |
| S26 | S13 OR S25 | 3733° |
| S25 | ("concordance") OR ("treatment implementation") OR (“treatment adherence”) OR (“treatment compliance”) OR | 1362° |
| S13 | ("intervention fidelity") OR ("intervention adherence") OR ("intervention implementation") OR ("intervention integrity") OR ("intervention delivery") OR (“intervention adherence”) | 2446° |
| S12 | ("treatment fidelity") OR ("treatment adherence") OR ("treatment implementation") OR ("treatment integrity") | 10297* |
| S5 | framework OR model OR process OR procedure OR strategy OR assessment OR indicat$ OR monitor$ OR checklist | 9301276 |

* Duplicates are removed from the search but included in the result count.

° Duplicates are removed from the search and from the result count.

Results: 721, 166 “Hits” after Screening

Figure 2.1 Search strategies: CINHAL

| # | Query | Last Run Via | Results |
|---|---|---|---|
| S11 | (S4 OR S5 OR S6 OR S7) AND (S9) | Interface - EBSCOhost Research Databases<br>Search Screen - Advanced Search<br>Database - CINAHL Complete | 245 |
| S10 | S4 OR S5 OR S6 OR S7 | Interface - EBSCOhost Research Databases<br>Search Screen - Advanced Search<br>Database - CINAHL Complete | 706,700 |
| S9 | (S3) AND (S8) | Interface - EBSCOhost Research Databases<br>Search Screen - Advanced Search<br>Database - CINAHL Complete | 1,487 |
| S8 | ( framework or model or theory) OR (process) OR (procedure ) OR (strategy) | Interface - EBSCOhost Research Databases Search Screen - Advanced Search<br>Database - CINAHL Complete | 1,091,68<br>2 |
| S7 | physical medicine and rehabilitation | Interface - EBSCOhost Research Databases Search Screen - Advanced Search<br>Database - CINAHL Complete | 5,152 |
| S6 | complex interventions in health | Interface - EBSCOhost Research Databases Search Screen - Advanced Search | 15 |
| S5 | surgery or operation or surgical procedure or surgical treatment | Interface - EBSCOhost Research Databases Search Screen - Advanced Search<br>Database - CINAHL Complete | 492,957 |
| S4 | ( physiotherapy or physical therapy or rehabilitation ) OR occupational therapy OR ( speech therapy or speech pathology or speech language pathology ) | Interface - EBSCOhost Research Databases Search Screen - Advanced Search<br>Database - CINAHL Complete | 232,880 |
| S3 | (S1) OR (S2) | Interface - EBSCOhost Research Databases Search Screen - Advanced Search<br>Database - CINAHL Complete | 3,136 |
| S2 | treatment delivery OR intervention delivery | Interface - EBSCOhost Research Databases Search Screen - Advanced Search<br>Database - CINAHL Complete | 1,074 |
| S1 | TX ( treatment integrity or treatment fidelity ) OR intervention integrity OR intervention fidelity | Interface - EBSCOhost Research Databases Search Screen - Advanced Search<br>Database - CINAHL Complete | 2,120 |

Table 2.1: Papers included in the Best-fit framework synthesis

| Author s | Year | Field/ Discpl. | Study design | Fidelity monitoring, measurement, or support method | Fid. Model | Fidelity Model Components, if reported , or direct quote/passage about fidelity extracted from paper. |
|---|---|---|---|---|---|---|
| Abell, et al | 2015 | CR<br>PA | SR | TIDIER items 10-12 | NR | When and How Much Describes the dose/schedule of the intervention including the following: (a) Intensity The intensity of exercise used in the intervention (e.g., % |

| | | | | | | |
|---|---|---|---|---|---|---|
| | | | | | | heart rate) (b) Frequency The frequency of exercise sessions (c) Session Time The duration of each individual exercise<br>session (d) Overall Duration The overall duration of the exercise<br>intervention<br>9 Tailoring Describes the what, why, when, and how of intervention titration, personalization, or progression<br>10 Modifications Describes any modifications to the intervention during the course of the study<br>11 How well: planned Describes strategies used to maintain or improve fidelity (how and by whom)<br>12 How well: actual Describes the extent to which the intervention was delivered as planned (if adherence or fidelity was assessed) |
| Adams, et al | 2012 | SLP | RCT | Audit of planned intervention sessions versus received sessions and adherence to written activity procedure as stated in the manual | Reported elsewhere | Full details in Adams, C. and Gaile, J., Forthcoming 2012, Managing Children's Pragmatic and Social Communication Needs in the Early School Years ( Manchester : Roundway Centre Publ.) (available at: http://www.roundwaycentre.org.uk/publications). |
| Allan, et al | 2018 | OT, PT, RH | PTCL FS | Assessment of fidelity to components (protocol?) | NR | NR |
| Allan, et al | 2018 | OT, PT, RH | PTCL FS | Assessment of fidelity to components (protocol?) | NR | NR |
| Anderson, et al | 2012 | PT<br>PA | RCT | Inferred from proxy. | NR | NR |
| Ang, et al | 2012 | PA | PTCL | Audio taped contacts to provide ongoing feedback and coaching to the MI health practitioner, on-going supervision and feedback. 25% of the audio taped phone contacts were formally evaluated for treatment integrity using the Motivational Interviewing Treatment Integrity (MITI) method. The MITI evaluators were blind to randomization. | NR | NR |
| Ang,et al | 2011 | PA | RCT | Phone contacts were taped to provide ongoing feedback and coaching to the therapists. supervision and feedback and checklist for integrity. | NR | NR |
| Asenlof, et al | 2009 | PT | RCT | Patients' reports on treatment content, PT session by session documentation of treatment content, as well as individual working sheets were collected in order to obtain an estimate of the proportions of | Bellg (NIH-BCC) | NR<br><br>Fidelity: Treatment dose was equivalent and within the stipulated range within and across conditions. Maintenance of treatment provider skills. |

| | | | | | | |
|---|---|---|---|---|---|---|
| | | | | participants in each condition that received the program as was intended. | | |
| Asenlof , et al | 2005 | PT | RCT | Patient reports of treatment content, therapists' documentation of treatment content session by session, and individual working sheets were collected to obtain an estimate of the proportions of subjects in each condition that received the program as it was intended. | Bellg NIH-BCC | NR<br><br>Fidelity: intervention was implemented with all the planned components. treatment dose monitored. |
| Aunger , et al | 2019 | PA | PTCL (RCT FS) | self-reported checklist, document/ recording assessment checklist. | Fidelity to theory. | NR |
| Avery, et al | 2014 | PA LI | PTCL RCT | standardised training, study manuals, ongoing process evaluation and video recording and assessment of patient sessions (intervention and usual clinical care) with fidelity check-list developed for the study | NR<br><br>Cites Bellg NIH-BCC | NR<br><br>Fidelity: presence/absence and appropriate use of intervention components. fidelity of delivery (adherence to intervention components). Adherence to the study protocol (fidelity assessment |
| Barber et al | 2015 | PA | FS | Assessed with NIH-BCC guidance. observer attended sessions in the initiation phase at each intervention school; intervention delivery was scored on a scale from 1 to 4. | NIH-BCC | Fidelity: Adherence to the intervention protocol, Delivery as per manual. |
| Barber et al | 2016 | PA | PILOT RCT | assessed according to NIH Behaviour Change Consortium guidance | NIH-BCC | Fidelity: Delivery as per manual |
| Baron et al | 2016 | RH | SR | TIDieR used to assess reporting of fidelity and adherence | Bellg/NIH-BCC | Fidelity of intervention delivery is extremely important as efficacy can only be determined if an intervention has been delivered as intended. |
| Bavan, et al | 2019 | PT PA | FS US | n/a | | |
| Beneciuk, et al | 2019 | PT | PT | n/a | Borelli NIH-BCC | Fidelity framework consisting of five domains (i.e., study design, training of providers, treatment delivery, treatment receipt, and treatment enactment. The PIPT program was designed to promote treatment fidelity by providing quality training that impacted key provider factors and that could be replicated. Thus, we incorporated quality improvement strategies (PIPT treatment checklist and booster training) and measures (physical therapist attitudes, beliefs, and confidence, described in greater detail below) to enhance treatment quality and the impact of training. |

| | | | | | | |
|---|---|---|---|---|---|---|
| Bennet , et al | 2011 | Pain PT | SR | n/a | Carroll | This is defined as the degree to which the intervention and control are delivered and assessed as intended. |
| Bergström et al | 2013 | PA | cRCT | fidelity was measured as the dose delivered. | NR | Intervention fidelity, defined as the extent to which a programme adheres to its programme theory (Fraser, 2009), is crucial to understanding the causal mechanisms (Mercer, DeVinney, Fine, Green, & Dougherty, 2007). In this study fidelity was measured as the dose delivered. |
| Birken, et al | 2018 | OT | DS | standardised training on use of the intervention, and ensuring skill acquisition during the training through active discussion of the occupational therapists' role during GLOW. An intervention Fidelity checklist was developed to monitor the extent to which the clinicians adhered to the content of the intervention manual. | Bellg NIH-BCC | NR<br><br>NIH-BCC model used in Bellg." fidelity to the intervention was enhanced, as recommended by Bellg and colleagues [27] by providing standardised training on use of the intervention, and ensuring skill acquisition during the training through active discussion of the occupational therapists' role during GLOW." |
| Blanche, et al | 2011 | OT | PP | Manualisation of therapeutic intervention recommended, with fidelity check against manual. | NIH-BCC | NIH-BCC: study design, training of providers, delivery of treatment, receipt of treatment, and enactment of treatment skills…. development of a treatment manual that includes information about treatment dose (length and number of contacts) and the specific content of each contact, standardization of therapist training, monitoring of the intervention with fidelity checklists, and inclusion of strategies to measure the client's comprehension and enactment of the intervention principles addressed. |
| Blencowe et al | 2015 | Surg | PP | n/a | Carroll 2007 | During a trial, it may be necessary to assess whether the intervention was delivered as intended (fidelity). Fidelity has been defined as 'how far those responsible for delivering an intervention actually adhere to the intervention as it is outlined by its designers' [14]. The degree to which intervention fidelity needs to be monitored within RCTs will largely depend upon the extent to which the trial protocol prescribes standardisation.<br>Fidelity is also referred to as compliance or adherence. |
| Blencowe et al | 2016 | Surg | PP | n/a | NR | Monitoring how surgical interventions are actually delivered in a trial (fidelity) is essential to inform the interpretation of results and subsequent implementation of interventions in practice. Three possibilities for recording and reporting fidelity were identified: the intervention, component or step is not delivered at all; an intervention, component or step from another trial group is delivered instead; or an entirely different intervention, component or step is delivered. |
| Boden, et al | 2016 | PT | MMS Nested in RCT | Nested MMS to investigate Rx differentiation, receipt, and enactment. Interviews, recordings, thematic analysis. | Borrelli NIH-BCC | the intervention is differentiable from standard care, is memorable, and adhered to by the patient. Treatment fidelity has four components [13]: (i) Integrity; was the treatment delivered as intended? (ii) Differentiation; did two treatments differ from one another as intended? (iii) Receipt; does the patient understand the treatments provided and are they equipped to perform them as intended?; and (iv) Enactment; does the patient enact the learnt skills and perform the intervention as intended? |
| Boden, et al | 2018 | PT | PTCL RCT | TIDIER | NR* | *Described elsewhere? Boden I, El-Ansary D, Zalucki N, Robertson IK, Browning L, Skinner EH, et al Physiotherapy education and training prior to upper abdominal surgery is memorable and has high treatment fidelity: a nested mixed-methods |

| | | | | | | |
|---|---|---|---|---|---|---|
| Bowen, et al | 2018 | SLP | FS RCT | Intervention details were recorded in speech therapy notes and retrieved following completion of follow-up. | NR | NR |
| Boyd | 2017 | PT<br>OT | PTCL<br>RCT | Manual, checklist for fidelity, observations/video analysis | NIH-BCC | study design, training of intervention providers, treatment delivery, treatment receipt and enactment |
| Boyle, et al | 2009 | SLP | RCT | evaluated modes of service delivery | NR | NR<br>evaluated modes of service delivery with MRC guidance to ensure fidelity |
| Breckenridge | 2015 | OT | PP | NA | Perplkova | Fidelity is achieved by ensuring: practitioner adherence to specified procedures; practitioner competence in delivering the intervention; and clear differentiation between interventions: that is, the intervention is substantively different to other approaches with similar goals. |
| Broekhuizen, et al | 2012 | LI<br>PA | PE<br>RCT | MI fidelity) was assessed by two MI experts, following the Motivational Interviewing Treatment Integrity code. Process evaluation. | RE-AIM | NR<br>including dose and fidelity monitored. Fidelity is defined as the extent to which the intervention was implemented as intended. To what extent was face-to-face counselling delivered as planned by MI guidelines?<br><br>DOSE NOT PART OF FIDELITY |
| Brogan | 2019 | SLP | PP | N/a | NIH-BCC | study design, training providers, delivery of treatment, receipt of treatment, and enactment of treatment skills |
| Brogan | 2018 | SLP | PP | N/a | NIH-BCC | study design, training providers, delivery of treatment, receipt of treatment, and enactment of treatment skills |
| Bronars, et al | 2017 | PA<br>LI | FidSt | Various: Protocol deviations manually recorded using a checklist form. Interviews, audits, etc. | NIH-BCC | design focuses on the methodological processes that ensure the study adequately assesses the proposed hypotheses in relation to a theoretical framework.<br>Training was designed to ensure satisfactory delivery of the intervention to study participants. Trainings were tailored to account for different backgrounds and past training experiences of the FHPs.<br>Treatment fidelity pertaining to treatment delivery includes ensuring that the content and dose are consistent as well as adherence to the manual.<br>Receipt ensures that participants received and understood the treatment provided.<br>Enactment assessment and monitoring of participant behaviour outside of the intervention |
| Bryant, et al | 2014 | PT | FidSt | This audio recording was reviewed by the site psychologists against specified criteria for<br>fidelity to the content and quality of<br>its delivery | NIH-BCC | treatment fidelity, a term that refers to the consistent and reliable delivery of interventions. achieving training fidelity: a specific intervention cannot be delivered until those delivering it have learned to do so in a standardized way. |
| Burkart | 2017 | PA | PE | semi-structured questionnaire completed by trained data collector observing sessions. Participant accelerometers. | Durlak and Dupre | Intervention fidelity (i.e., adherence, compliance, integrity). degree of which the intervention was delivered as originally planned |
| Busse, et al | 2017 | PT | RCT | Fidelity of the physical activity intervention was measured using a combination of self-report checklists, independent analysis of audio recordings, and a self- | Not reported, but see Quinn et al, 2016, for | NR |

| | | | | | | |
|---|---|---|---|---|---|---|
| | | | | assessment completed by the intervention coaches. | description. | |
| Butel, et al | 2015 | LI<br>PA | cRCT | fidelity monitoring, which documented how well intervention components were being implemented, assessed intervention adaptations (expected given vastly different intervention site contexts), and identified ways to improve intervention delivery. | NR<br><br>Cites Gearing | NR |
| Bysterveldt et al | 2010 | SLP | SS | Analysis of videotaped sessions. | NR | NR |
| Campbell, et al | 2015 | PA | FidSt | Process evaluation, quantitative and qualitative | NR | Fidelity is whether the intervention is delivered as expected whether and how variations in delivery occurred. To what extent was the intervention delivered as planned? To what extent was the intervention delivered as planned? IN what ways, if any, did the teachers amend the programme? and<br>What were the reasons for any amendments? |
| Carlson, et al Lifestyle intervention for adults with spinal cord injury | 2019 | RH<br>LI | RCT | Interveners received 30 hours of standardized training, were assessed monthly in session delivery using a standardized rating scale, and attended weekly troubleshooting meetings to mitigate protocol drift | NIH-BCC | NR |
| Carlson, et al | 2019 | PA<br>SCI | RCT | multi-faceted treatment fidelity plan : standardized training and check ins. | Bellg<br>NIH-BCC | To ensure that the intervention protocol was properly implemented, we adhered to a multi-faceted treatment fidelity plan consistent with guidelines for monitoring complex interventions. According to this plan, interveners received 30 hours of standardized training, were assessed monthly in session delivery using a standardized rating scale and attended weekly troubleshooting meetings to mitigate protocol drift.50–52 All interveners were blind to the study hypotheses and design. |
| Carroll | 2007 | Fidelity | PP | n/a | Carroll | 1.Adherence: whether "a program service or intervention is<br>being delivered as it was designed or written.<br>2.exposure or dose: the amount of an intervention received by participants (frequency and duration as specified in protocol…)<br>3.quality of delivery: the manner in which therapist delivers intervention. E.g.: adherence achieved but the intervention delivered badly. It concerns whether an intervention is delivered in a way appropriate to achieving what was intended. If the content of an intervention is delivered badly, then this may affect the degree to which full implementation is realised.<br>4.participant responsiveness: how far patients respond to or are engaged by an intervention. |

| | | | | | | |
|---|---|---|---|---|---|---|
| | | | | | | 5.programme differentiation: Identification of an intervention's essential components.<br><br>CARROLL: **Adherence**: content, coverage, frequency, duration. Adherence is essentially the bottom-line measurement of implementation fidelity. If an implemented intervention adheres completely to the content, frequency, duration, and coverage prescribed by its designers, then fidelity can be said to be high. The **content** of the intervention may be seen as its 'active ingredients'; the drug, treatment, skills, or knowledge that the intervention seeks to deliver to its recipients. Subcategories of adherence concern the frequency, duration, or coverage of the intervention being delivered, i.e., what is more broadly defined as "**dose**" Identifying these essential components also provides scope for identifying adaptability to local conditions. Moderators include complexity, delivery qual… |
| Case-Smith, et al | 2011 | OT | PS | Customized Fidelity measure | NR | NR |
| Case-Smith, et al | 2011 | OT | UCT | | Adherence to Rx principles. | NR<br><br>Fidelity: Adherence to Rx principles. |
| Casey, et al | 2018 | PA<br>PT | PTCL<br>RCT | Treatment fidelity will be assessed by a health psychologist who is highly experienced in the delivery of group psychological interventions for chronic pain using ACT. All eight ACT sessions from one treatment group will be audio recorded and reviewed. An ACT treatment fidelity tool that has been modified for chronic pain will be used to evaluate the intervention. Treatment fidelity of the physiotherapy components of the trial will be assessed review the checklists and notes at monthly intervals | Borrelli (NIH-BCC) | Fidelity: Fidelity to protocol. |
| Casida, et al | 2012 | Surg<br>RH | PS | Log to check compliance to protocol (nurses) | NIH-BCC | Treatment fidelity consists of five components of design (for accurate testing of the study's clinical process), training (of those researchers providing the intervention), delivery (to monitor the intervention ac-curacy), receipt (to ensure that the patient is able to perform the intervention), and enactment (the ability to perform the intervention in real life settings) |
| Chesworth, et al | 2015 | RH | FidSt | Assessment of nurse's logs<br>Focus on fidelity of treatment delivery on study. | NIH-BCC | These recommendations include five areas of treatment fidelity: study design, provider training, treatment delivery, treatment receipt, and enactment of treatment skills. Focus on fidelity of treatment delivery on study. |
| Chewning, et al | 2019 | PA | RCT | Master teacher's observation/a 40-item dichotomous checklist. | Borrelli, NIH-BCC Re-AIm | "Also, demographic information, fidelity of course delivery, course size and attendance, and fidelity of course receipt and enactment in terms of participant tai chi home practice was collected. Trained researchers |

| | | | | | | |
|---|---|---|---|---|---|---|
| | | | | | | collected physical, confidence and executive function data at community sites using specific stations for each measure with participants systematically rotating between these stations.” |
| Clark, et al | 2014 | RH | PP RCT | 30 hours of standardized intervener training; (b) one monitoring session per month in which each intervener’s adherence to the protocol was assessed using a specialized rating scale; and (c) weekly supervisory protocol adherence meetings. | NR | A balance between protocol adherence and clinical judgment must be maintained. We achieved this requirement through two strategies. First, the six intervention modules served as a toolbox demarcating the core components of the intervention and the range of appropriate content and provided troubleshooting guidelines. Second, interveners were expected to adhere to a set of theoretically grounded, overarching principles related to pressure ulcer risk when tailoring the sessions to be participant- and situation-specific |
| Cooke, et al | | RH | PTCL RCT | training and a standardised procedure manual (detailing protocol, plans for dealing with intervention fidelity issues, and monitoring the delivery and receipt of the intervention. | TDF<br><br>Cites Spillan e | NR<br><br>Fidelity: Theoretical Domains Framework, delivery, and receipt of the intervention. |
| Cox, et al | 2018 | PT | FS PILOT RCT | subjective description of case notes by the study team. The co-applicant physiotherapists rated each physiotherapy session for optimisation using data from the study documentation. Optimisation assessment of whether or not any adaptations had been made and whether or not the session had been completed | | Fidelity is usually interpreted as the consistent delivery of intervention components.112 However, physiotherapy interventions do not always benefit from consistency as they involve the revision of treatment plans to account for changing and uncertain experiences.113 Evidence suggests that the fidelity to the treatment theory is more important114–116 than consistent delivery of the intervention. We therefore evaluated treatment optimisation to the patients’ needs and capabilities. Adherence to the prescription. Assessment of overall treatment optimization. It follows that optimisation, or ‘fidelity of function’ is more an appropriate construct for assessing implementation quality than ‘fidelity of form’,114 ensuring congruence with the intervention theory. |
| Cristian sen, et al | 2010 | PT PA | RCT | NR | NR Cites Pereple tchikov a | NR |
| Cutchin , et al | 2009 | OT | FS RCT | Manual. Feasibility is being measured by the rate of recruitment, representativeness of the study sample, and retention rates. | NIH-BCC<br><br>Cites Frank | fidelity in design, training, delivery, receipt, and enactment.<br><br>Frank https://doi.org/10.1177/0733464807308621 |
| Dean, et al | 2018 | PA RH | PILOT RCT | attendance registers, accelerometery, exercise ‘homework’ diaries, trainer completed session checklists and video analysis of (early, middle, and late programme) training sessions | NR | NR<br><br>No definition of fidelity: Adherence to protocol/manual. |

| | | | | | | |
|---|---|---|---|---|---|---|
| Dean, et al | 2016 | RH stroke | PTCL Pilot RCT | attendance, accelerometery, exercise diaries, session checklists and video analysis of selected training sessions in each programme. | NR | NR<br><br>Fidelity: adherence to the intervention manual by participants and trainers |
| Deary, et al | 2018 | SLP | Pilot RCT | Sessions audio recorded and structured analysis of the content of intervention for treatment fidelity and inter-treatment contamination. monitoring the content of the sessions via real-time clinical supervision and retrospective content analysis of contemporaneous case notes. | NR | NR<br><br>Fidelity of delivery, protocol violations. |
| Deary,e t al | 2018 | SLP | RCT DS | assessed by monitoring the content of the sessions via real-time clinical supervision and retrospective content analysis of TM's contemporaneous case notes. | NR | NR<br><br>No results of Fidelity Assessment. |
| Desveaux, et al | 2016 | PT PA | PTCL RCT | Fidelity will be monitored throughout the study through semi-annual check-ins with community facilities. Facilities will explicitly outline the operationalization of the intervention at their respective facility, including frequency, duration, supervision, attendance monitoring, and individual program components. | NR | NR |
| DeVito, et al | 2011 | RH | FidSt | n/a | Custom | Delivery: the extent to which the intervention is delivered as intended; (b) receipt: the extent to which the intervention is received as intended; and (c) technology acceptance: the extent to which the participant has positive perceptions, attitude, and intention to use a system. Relationships purported in the framework include the following: (a) Intervention fidelity extends beyond delivery to include receipt and technology acceptance (perceived ease of use, perceived usefulness, attitudes toward use, and intention to use). (b) There is a reciprocal relationship between delivery and receipt (i.e., qualities of delivery affect receipt and vice versa). And (c) human factors (technology acceptance) moderate the relationship between delivery/receipt and ultimate adoption (use of technology). |

| | | | | | | |
|---|---|---|---|---|---|---|
| | | | | | | Measurement of delivery typically includes assessing whether all the intervention components and activities were delivered and implemented in the proper manner. Adoption is the extent to which the individual participant uses the technology-based behavioural intervention. It is akin to terms such as intervention usage, utilization, and intervention dose and should not be confused with the use of the term for describing diffusion of innovations (how new ideas and technologies spread among groups. Enactment is the extent to which the participant performs the behaviours that the technology-based behavioural intervention is intended to promote (e.g., follow an exercise regimen, monitor health indicators). Because adoption moderates the relationship between the intervention and treatment effects (enactment), it is important to quantify both to determine the strength by which one can conclude that the intended outcomes were indeed due to the use of intervention. Because neither adoption nor enactment measures how well the intervention was delivered as conceived and planned, they are not included as components of intervention fidelity. |
| Di Rezze | 2012 | RH<br>PT<br>OT | NR | | IFF | evaluating the adherence of the therapist to an intervention and differentiating therapist behaviours between interventions. Therapist behaviours “client responsiveness,” a concept that is also applicable to paediatric rehabilitation. Such items examined client or client-therapist interaction behaviours. Examples include, “how receptive and/or engaged was the client during the session? Fidelity includes therapy process (e.g., assessment) and delivery process (e.g., rapport). |
| Di Rezze et al | 2014 | RH | NR | n/a | IFF | actors that mediate intervention fidelity focus on therapist adherence to delivering the intervention as expected and include only therapist behaviours. Moderating factors of intervention fidelity examine concepts beyond the therapist's adherence to the intervention. Examples include therapist quality of intervention delivery, client attributes independent of the therapist, and client attributes dependent on the therapist (i.e., therapist-client interaction). |
| Di Rezze, et al | 2013 | PT<br>OT | FidSt<br>PP | Paediatric Rehabilitation Observational measure of Fidelity (PROF) developed for study. Scoring video of Rx. | IFF | The fidelity measure created to support this process was based on a conceptual model, the implementation fidelity framework (IFF). In this model, therapist adherence in delivering the intervention mediates intervention fidelity. potential moderators may occur before intervention delivery, such as the complexity of an intervention (i.e., comprehensiveness of policy) and defining strategies to facilitate therapy better. Quality of therapy delivery and participant responsiveness moderators were related to examining the deliverer (i.e., therapist) and recipient (i.e. client) within an intervention session. Adherence: details of content, coverage, frequency, and duration. |
| Di Rezze,e t al | 2014 | RH | PP | CS | NR | NR |
| Dillon, et al | 2018 | | PTCl<br>PE | quantitative (fidelity checklist score, number of completed sessions, survey data and a habit formation scale), as well as qualitative (open responses from program staff and semi-structured interviews with study participants) data | MRC | program adherence (fidelity): How well did the specialists deliver the program to participants?<br>complete delivery (dose delivered) To what extent were all of the intended components of the v-LiFE program delivered to participants?<br>participant receipt (dose received: To what extent were participants engaged/satisfied with the v-LiFE program?<br>participant enactment: To what extent were the participants completing the prescribed activities? |
| Drew, et al | 2016 | CI | CS<br>PP | Evaluation of recordings. | NR | fidelity of the delivery of the intervention in line with the theoretical framework on which it was developed |

| | | | | | | |
|---|---|---|---|---|---|---|
| Dunn, et al | 2017 | OT<br>LI/HP | FidSt | Hand coding of observations/ transcripts of randomly selected sessions against checklist made for study, based on Gearing and components of OPC. *Blind coding/assessment of fidelity in Rx sessions.* | Geariing and Bellg (NIH-BCC) | Gearing Model of NIH-BCC. These authors described four key elements of fidelity: (a) intervention design, (b) intervention training, (c) monitoring intervention delivery, and (d) monitoring receipt of intervention. Researchers must use a clearly outlined theoretical framework, program goals, and a consistent methodology to measure the fidelity of an intervention design.<br>ESSENTIAL actions of the intervention and UNIQUE actions of the intervention.<br>It is also important to monitor specific aspects of intervention receipt, including dose and participant comprehension, resistance/acceptability, attendance, and/or knowledge before and after the intervention. assessed intervention receipt by monitoring participant attendance and participation throughout the study. We queried parents regarding acceptability of OPC for their families and their goals via post intervention questionnaires. |
| Elinder, et al | 2012 | PA | UCT | Process evaluation. Fidelity to school action plans was evaluated through interviews with health teams guided by a checklist. | NR<br><br>Cites Fraser | Fidelity to the programme as a whole was assessed in relation to whether schools had implemented all components in the programme according to the logic model.<br>Fidelity, defined as the extent to which a programme adheres to its programme theory |
| Eyre, et al | 2016 | CR | PTCL Pilot RCT | A customized, 13-item intervention fidelity checklist. audio recordings by intervention facilitators and then reviewed and coded (using the fidelity checklist) by two *independent researchers.* | NR | NR<br><br>Fidelity: Adherence by facilitators to the intervention protocols |
| Eysenbach, et al | 2019 | PA<br>RH | PTCL RCT | "expert auditing of calls, monitored study website, coaching call notes. Monitoring all phases. | NIH-BCC | "study design, provider training, delivery of treatment, receipt of treatment, and enactment of treatment. " "**study design** area focuses on practices that ensure study procedures and implementation are in line with current theory and clinical processes. Study design fidelity goals include ensuring that conditions are congruent with relevant theory and practice, ensuring equivalent treatment dose within and across conditions, and planning for implementation setbacks." **Provider training: "**strategies that address preparation for uniform delivery of treatment by providers/coaches." "standardized training; this ensures that training is conducted similarly for all providers. " "Another goal of provider training is minimizing drift in provider skills. **"Delivery**: "Fidelity of treatment delivery focuses on ensuring the intervention is delivered as intended. Many of the concerns within delivery of treatment overlap with strategies for training and study design, including controlling for provider differences and adhering to created protocols; however, this area further addresses differences within treatment conditions and minimizes contamination " "**treatment receipt** involves strategies and monitoring of a participant's ability to understand and adopt treatment-related behavioural skills and cognitive strategies. " "participants' comprehension and ability to utilize digital media in delivering content and tracking goals" "**enactment** of treatment described as strategies aimed at monitoring and improving participant ability to perform treatment-related behavioural skills and cognitive strategies in relevant real-world settings." |
| Ferrante, et al | 2019 | PT | PTCL RCT | systematic process, specific therapist training for | Gearing | Key components of intervention fidelity include a well-designed framework, therapist training, monitoring of treatment delivery, and monitoring |

| | | | | | | |
|---|---|---|---|---|---|---|
| | | | | motor learning principles and techniques as well as administration of outcome measures, establishment of interrater reliability with outcome measures, treatment session observation, critique of participant | | of treatment receipt (by participants) |
| Flynn<br><br>SMART | 2018 | Pain<br>RH | RCT | Role-playing ensuring that team members understand the protocol. All contact with participants is scripted and randomly and regularly reviewed by the principal investigator. Treatment delivery is monitored by the interdisciplinary pain management centre clinical team leader | NIH-BCC | NR |
| Foster, et al | 2016 | RH<br>PT | FS<br>PILOT<br>RCT | PTs recorded full details of the advice and treatments, number and mode of treatment sessions, any non-attendance, acupuncture points used, any sensations during acupuncture treatments and any adverse events on specifically designed CRFs. Audited against the PT clinical notes to ensure accuracy and to determine protocol adherence. | NR | NR<br><br>Fidelity to protocol<br><br><br>Audited against the physiotherapists' clinical notes to ensure accuracy and to determine protocol adherence by participating physiotherapists before collation by the research team in order to fully describe the interventions delivered. Where protocol deviations were noted, these were discussed with the physiotherapists involved in order, whenever possible, to enhance adherence to the agreed intervention protocol. |
| French, et al | 2015 | PR | SR | five-area (eight-element) intervention fidelity tool, | NIH-BCC | Intervention fidelity (study design, training of providers, delivery of treatment, receipt, and enactment of treatment?<br>Study design: was the guideline or protocol used to guide the study published and was it clearly identified? were standardized or validated tools used to measure patient reported outcomes?<br>Training of providers: formal training of providers related to the guideline or protocol used, and was an intervention manual used to guide providers?<br>Delivery of Treatment: core treatment interventions consistent with the guideline or protocol used to develop the intervention manual and/or to guide the study? "was there assessment of response to treatment at specified timeframes?<br>Receipt of Treatment: Was there any mention and/or measurement of concordance of patient and provider understanding of the problem and/or treatment recommendations?<br>Enactment: patient's ability to engage in the treatment recommendations in daily life |
| French, et al | 2011 | PA | PTCL | Manualized intervention. Taped intervention sessions are being analysed to quantify the extent | Bellg (NIH-BCC) | Provider fidelity of delivery to the intervention manual and maximise recipient fidelity of receipt and enactment of the intervention techniques. If interventions are not delivered or received as intended (i.e., as per protocol) then it is difficult, if not impossible to be certain that the |

| | | | | | | |
|---|---|---|---|---|---|---|
| | | | | to which each intervention technique was delivered as specified by the intervention manual. Selection of patients were interviewed immediately after receiving the intervention to monitor intervention receipt. | | results can be attributed to the intervention itself. fidelity in relation to delivery and receipt of the intervention |
| Fuentes, et a; | 2014 | PT | RM | Therapist adherence was based on how closely the therapists followed the experimental protocol assessed by videotaping all treatment sessions, of which 28 (20%) were randomly selected for evaluation . Two research assistants not involved with the study separately rated each session regarding treatment fidelity. | NR | NR |
| Fuentes, et al | 2014 | PT | expContSt | Therapist adherence was assessed by videotaping all treatment sessions, of which 20% were randomly selected for evaluation. Two research assistants not involved with the study separately rated each session regarding treatment fidelity. *Independent assessments.* | NR<br><br>Cites Moncher | Therapist adherence was based on how closely the therapists followed the experimental protocol. |
| Furnes et al | 2018 | PA | PE | Process evaluation of fidelity | MRC | NR |
| Galaviz, et al | 2014 | PA | SR | RE-AIM criteria in checklist | RE-AIM | the duration and frequency of the intervention, the extent to which the protocol was delivered as intended, and the cost of delivery. Additional implementation indicators used include whether articles reported the theoretical framework of the intervention, the consistency of implementation across settings and delivery agents, the degree to which the participants received intervention components, and the use of qualitative methods for measuring implementation. |
| Gibson, et al | 2016 | PR | SR | five-area (eight-element) intervention fidelity tool, | NIH-BCC | Study Design, Provider Training, Treatment Delivery, Treatment Receipt, and Treatment Enactment |
| Giesbrecht, et al | 2017 | OT<br>PT<br>RH | RCT<br>FS | Trainers indicated any protocol deviations on the checklists and completed a trainer post-treatment evaluation form after EPIC | NR | Adherence to protocol |

| | | | | | | |
|---|---|---|---|---|---|---|
| Gladwell, et al | 2016 | PT | QualStdy | Semi-structured interviews | Carrol | adherence and moderating factors. Adherence is defined as “how far those responsible for delivering an intervention actually adhere to the intervention as it is outlined by its designers”11 and includes the subcategories of treatment content, coverage, frequency, and duration. Moderators are factors that influence the degree of fidelity with which an intervention is implemented and include intervention complexity, facilitation strategies, quality of delivery, and participant responsiveness to a treatment program. |
| Glombiewski, et al | 2010 | CP BC | RCT | Manual, videotape analysis. | NR | All therapists were trained in a standardised treatment plus biofeedback and were supervised weekly by licensed cognitive-behavioural therapists and supervisors. Treatments were described in detailed session-by-session protocols to maintain treatment fidelity. Selected videotapes of therapy sessions were discussed during supervision contacts to ensure adherence to protocols. |
| Glombiewski, et al | 2010 | Pain RH | | Selected videotapes of therapy sessions were discussed during supervision contacts to ensure adherence to protocols. | Fidelity to manuals | No theory reported.<br>“Treatments were described in detailed session-by-session protocols to maintain treatment fidelity.” |
| Godecke, et al | 2015 | SLP | FidSt | Therapy fidelity was measured according to the TIDiER statement with treatment adherence and treatment differentiation monitored throughout the trial. sessions were reviewed by the independent therapy fidelity monitor and feedback was provided to therapists as part of the ongoing adherence to the treatment protocol. | NR | NR |
| Godfrey, et al | 2016 | PT | PTCL | sessions from every PT will be rated in terms of adherence to the manual and checklist/modified fidelity measure | NR | Fidelity to manual/protocol. |
| Golos, et al | 2011 | OT | qRCT | consultation and monitoring sessions were documented using forms for the purpose of determining treatment fidelity. Customised Fidelity measure | NR | NR<br>No results of Fidelity assessment. |
| Graham, et al | 2018 | PT PA | PTCL cRCT FS | Observations of the delivery of a sample of the training will be conducted in each intervention home. Semi-structured interviews with staff and trainers. Process evaluation. a trial-specific observational tool designed to record instances of care | NR | Intervention fidelity and adherence (training, content, delivery, attendance |

| | | | | | | |
|---|---|---|---|---|---|---|
| | | | | behaviours reflective of skills (that could have been) learned during SCTP training will be developed and tested in more than one site. | | |
| Granbom et al | 2019 | OT | PTCL | demonstrate the last session's exercises to the OT on the following session and show the logs of activities. Sessions are audio-recorded, and study team members review 10% of the sessions. bi-weekly meetings with the OT and PI case presentations evaluated in terms of fidelity to intervention delivery | NIH-BCC | "The fidelity plan, based on the National Institutes of Health (NIH) Behaviour Change Consortium, addresses fidelity through design (distinct program based on theory), training (using established LiFE program training, home safety training, and program manual), delivery (reminder calls the night before sessions), engagement (records of home sessions by date and duration), and receipt (completing checklists on program engagement)." |
| Granbom, et al | 2019 | OT | PTCL<br>PS<br>RCT | Sessions are audio-recorded,<br>and study team members review 10% of the sessions. bi-weekly meetings with the OT and (PI) case presentations evaluated for fidelity to intervention delivery. | NIH-BCC | addresses fidelity through design (distinct program based on theory), training<br>(using established LiFE program training, home safety training, and program manual), delivery (reminder calls the<br>night before sessions), engagement (records of home sessions<br>by date and duration), and receipt (completing checklists on program engagement). To assure enactment,<br>participants in the LIVE LiFE group demonstrate the last session's exercises to the OT on the following session and show the logs of activities. |
| Guagliano, et al | 2019 | PA | FS<br>RCT | Participant questionnaires. | NR | NR |
| Gunn, et al | 2018 | | | fidelity of a random sample of a minimum of 25% of the delivered sessions will be assessed using audio recordings of the session. This sample will include at least two recordings of each session type. Checklist used, two team members not Rx. | NIH-BCC<br><br>MRC | NR<br><br>Process evaluation for Fidelity. |
| Hahne | 2011 | PT | PTCL<br>RCT | comprehensive treatment manual, initial training of PTs, a monthly teleconference involving all treating PTs, clinical notes directing PT along decision making algorithms, reviewing the clinical notes of every participant at three points during their treatment program, and the use of standardised participant | NR<br><br>Cites Borrelli | NR |

| | | | | | | |
|---|---|---|---|---|---|---|
| | | | | information sheets are methods chosen to ensure that all participants receive treatment from PTs that is standardised, accountable, and reproducible. | | |
| Hand, et al | 2018 | PT<br>OT | SR | n/a | Carroll<br><br>Dane and Schneider. | Fidelity is the faithfulness of an intervention to its underlying therapeutic principles and clinical guidelines. Fidelity consists of five key components: (1) adherence, (2) quality of delivery, (3) exposure, (4) participant responsiveness, and (5) program differentiation. Adherence refers to the extent to which program components are delivered as intended, whereas quality of delivery is a subjective aspect of treatment delivery that extends beyond delivery of prescribed content. For example, aspects of quality of delivery may include clinician enthusiasm or attitude. Exposure refers to the number, length, or frequency of intervention sessions or the frequency with which intervention techniques are implemented. Participant responsiveness includes participants' judgments about the outcomes and relevance of an intervention (Carroll et al, 2007) and is a key aspect of intervention fidelity. Finally, program differentiation refers to how the intervention being delivered is different and distinguishable from other interventions<br>. |
| Hankonen, et al | 2016 | PA | PTCL cRCT | Intervention facilitators keep track of components delivered, as well as the quality of delivery (e.g., interaction elements), by filling in a self-assessment form after each session, to assess whether the intervention was delivered as intended and to ensure high fidelity | NIH-BCC | receipt and use of intervention materials (e.g., use of workout and activity break videos) and enactment of the BCTs taught in intervention classes |
| Harris, et al (Nauta) | 2013 | PA | PS,<br>FS | Inferred from proxy. | NR | "Another strength of the study was that both interventions were implemented by highly skilled interventionists with years of training in their fields suggesting high levels of treatment fidelity by those implementing the intervention and assessment." |
| Hart, et al | 2012 | RH | PP | n/a | NR | Fidelity may be defined as the extent to which the core components of treatment have been delivered as intended. concepts of treatment receipt and treatment enactment. Treatment receipt refers to the extent to which the patient understands the strategies or techniques taught and demonstrates the capacity to use them. For this purpose, one could administer pre- and posttreatment tests of knowledge related to treatment. Treatment enactment, which has to do with whether the participant actually uses the learned strategies in day-to-day life, is more challenging to measure but could be ascertained using self-report and proxy report instruments given at some point after the trial. |
| Harwood, et al | 2018 | PT<br>OT<br>HP/LI | PTCL<br>FS | Records of therapy sessions undertaken, and self-directed activity will be examined, and video-recorded therapy sessions will be assessed | NR | We will undertake a process evaluation, studying fidelity, understanding mechanisms and context. Implementation (delivery of intervention), includes fidelity (quality of delivery) and dose (quantity of delivery). |

| | | | | | | |
|---|---|---|---|---|---|---|
| | | | | qualitatively for fidelity. Mechanisms of impact and contextual factors include engagement and adherence. | | |
| Healey, et al | 2018 | PA | FS<br>RCT | Audio recording of intervention sessions and intervention fidelity checklist specifically developed for the trial to assess whether components of the consultation intended to be included | Gearin g | fidelity – the degree to which the intervention is delivered as intended. which elements of this intervention were delivered. |
| ***Hildebr and*** | | | | | | |
| Hill, et al | 2014 | RH | cRCT | Therapists complete the online training programme. Patient's unit lists are checked. education and training are repeated for new staff on the unit. | NR | NR<br><br>Adherence to protocol |
| Hinckle y, et al | 2013 | SLP | NR | n/a | Moncher and Prinz | Treatment integrity refers to how well a treatment condition was implemented as planned. treatment differentiation refers to whether the treatment conditions being studied differed from each other sufficiently so that the intended manipulation of the independent variable can be assumed to have occurred. Both of these concepts are important to consider because it is possible to administer a treatment as planned without differentiation from a comparison treatment or to successfully differentiate two treatments in a research study without implementing the treatment with a high degree of integrity |
| Hoekst ra,et al | 2018 | PA<br>(MI) | | n/a | MICAS | It is expected that the MICAS contains four subscales related to the factors: Acceptance (10 items), Partnership (8 items), Evocation (3 items), and MI non- adherence (5 items). |
| Hofma n, et al | 2013 | PA<br>PT | PTCL<br>RCT | BET is delivered by the same BET-therapist, who will be intensively trained in the use of the program and will receive a detailed trainer manual. Regularly, announced visitations during the Rx phase will occur in order to assure adherence to the treatment protocol. Individual deviations from the Rx protocol will be recorded. | NR | high treatment integrity. The latter refers particularly to a potential mixture between the study group and the control group through a communication about intervention contents between patients and therapists within each rehabilitation centre, as well as individual deviations from the treatment protocol. |
| Holland , et al | 2018 | SLP | FS | Video and audio footage assessed for fidelity with MIS Checklist. | | intervention maintains fidelity to the core principles of the therapy. session number, duration, and content; therapist background, training, and support. |
| Holland s, et al | 2013 | PT | | Adherence to the intervention by therapists will be | NR | NR |

| | | | | | | |
|---|---|---|---|---|---|---|
| | | | | assessed during their involvement in the trial by A. Wimperis and K. Hollands through video observation at weeks 2 and 6 of each therapists' first treatment period. Further training for the therapist will be provided, if necessary, to improve compliance with treatment protocols. | | |
| Holt, et al | 2018 | PA | RCT | *assessed by facilitator talk time, checklist and direct observation of the facilitator behaviour and conduct at sessions. | NIHBCC Linnan and Steckler's process evaluation framework | **Design**: Ensure the same treatment dose within conditions. Ensure an equivalent dose across conditions. Plan for implementation setbacks. **Training:** Standardise training. Ensure provider skill acquisition. Minimise 'drift' in provider skills. Accommodate provider differences **Delivery:** Control for provider differences. Reduce differences within treatment. Ensure adherence to treatment protocol. Minimise contamination between conditions. **Receipt:** Ensure participant comprehension. Ensure participant ability to use cognitive skills. Ensure participant ability to perform behavioural skills. **Enactment:** Ensure participant use of cognitive skills. Ensure participant use of behavioural skills. |
| Hosseini, et al | 2018 | PA Tai Chi | RCT | We assessed the treatment fidelity of study by a standard guideline (Bellg et al, 2004). To ensure the exercise program was administered by the same frequency and duration of sessions for all participants. | NIH-BCC | NR |
| Hurd, et al | 2017 | PT | PTCL RCT | weekly teleconference meetings, in which the documentation from training is reviewed, and discrepancies addressed. Video recordings of a full training session are compared periodically. Finally, each therapist visits the other centre at least once a year to observe and discuss the training. | NR | NR |
| Hurley, et al | 2019 | PT | cRCT | PT behaviour was assessed during delivery of the SOLAS intervention by audio coded by one blinded expert rater and physios' self-reported checklists to evaluate fidelity to | Borrelli | NR |

| | | | | | | |
|---|---|---|---|---|---|---|
| | | | | the intervention content | | |
| Hurley, et al | 2016 | PT | PTCL cRCT | direct observation and audio recording by researcher, PT self-report to assess the content and quality of treatment fidelity during the trial. interviews with intervention PTs .Fidelity will be assessed and reported by separate evaluators from the outcome evaluators. | | |
| Jafar, et al | 2016 | LI | FS | Process evaluation | NR | fidelity defined as the proportion of a) the planned orientation sessions delivered to physicians and nurses, b) the prescription of FDC to eligible participants, c) the delivery of MC to eligible participants, and d) the telephone follow-ups. |
| Jago, et al. | 2015 | PA | PE RCT FS | Process evaluation interviews | NR | dose delivered, the reach (or number of people who receive the intervention), fidelity (extent to which the intervention was delivered as planned), implementation (how well the programme was implemented) and context which provides critical information on the environment in which the programme was delivered |
| Jaka, et al | 2016 | LI | SR | Modified NIH-BCC Fidelity checklist. | NIH-BCC | treatment design, provider training, and treatment delivery, receipt, and enactment. |
| Johnston, et al | 2009 | OT | PP | Fidelity to manual | NR | NR<br><br>No definition of Fidelity |
| Johnston, et al | 2019 | SLP | RCT | Sessions were all video-recorded. Each session had a Treatment Fidelity Checklist that contained the essential elements of each session for the therapist. Therapists rated each element of therapist fidelity. A random sample of 10% of each therapist's video-recorded sessions was examined quarterly by an independent observer to determine inter-rater reliability. | NR | therapist integrity in the delivery of treatment as intended and (b) parent adherence to the delivered treatment |
| Jørgensen, et al | 2012 | PT | RCT | Log-books on adherence, fidelity and context were held during the intervention. | NR | adherence, context and fidelity and the interplay between adherence and contextual events. implementation is reflected in the intervention dose received by the participants (i.e., adherence) and the fidelity (i.e. the quality of intervention delivery). |
| Karas | | MT PT | | | | |
| Katz et al | 2015 | Surg | PP | n/a | Cites Nelson and Cordray | Intervention fidelity refers to the extent that the intervention is delivered in an identical fashion to each subject.<br>To address these issues of intervention fidelity and learning curve, the surgical investigators should meet prior to the study launch in order to develop consensus on the precise surgical protocol, including the decisions |

| | | | | | | |
|---|---|---|---|---|---|---|
| | | | | | | to be made in the face of particular intraoperative findings. |
| Kearney, et al | 2006 | LI/HP | FidSt Pilot | Fidelity to protocol not assessed or recorded during trial. | Bellg<br><br>Leventhal and Friedman | Full execution of an intervention has been termed treatment fidelity, which has two components: integrity (the research team delivers the intervention as intended) and differentiation (the groups' exposure to the intervention differed as intended). Integrity includes treatment receipt (participants absorbed the knowledge and skills imparted in the intervention) and treatment enactment (participants used the skills in the manner intended. treatments be supported by a theoretical explanation for their mechanism of action so that they can be adapted without losing integrity, and failures can be explored in light of the theory. Differentiation of groups is the second major component of fidelity. |
| Keogh, et al | 2018 | PT | FS | process evaluations to<br>assess the fidelity to, and implementation of, intervention components | NR<br><br>MRC and Borrelli cited. | Fidelity guidelines suggest that following training, providers should be competent to deliver the intervention. Fidelity evaluations involve the assessment of what has taken place |
| Kerr et al | 2018 | PA | cRCT | Evaluation of participation (dose?) and enactment. | NR | NR |
| Kippling, et al | 2016 | PA | FS<br>cRCT` | Process evaluation | NR | NR<br><br>reach and dose, some enactment |
| Lambert, et al | 2018 | PA | Pilot RCT | Delivery fidelity was tracked using Web usage statistics. Fidelity of receipt 5-point Likert response scale. assessed participants' perceived ability to use the intended BCTs by asking participants to rate their confidence in using specific BCTs. to assess enactment, asked participants if they had used specific BCTs related to BA in the last 2 months using a binary scale (yes/no) | NR<br><br>Cites Borrelli (NIH-BCC) | Intervention fidelity was conceptualized and measured in the domains of design fidelity, training fidelity, quality/completeness of delivery, participant receipt, and enactment. fidelity of intervention delivery, receipt, and enactment (use of techniques). |
| Lambert, et al | 2017 | PA | SR<br>Fid | NIH-BCC criteria | NIH-BCC<br>MRC | Study Design, Provider Training, Intervention Delivery, Intervention Receipt and Enactment. Study Design is concerned with whether a study adequately tests its hypotheses in relation to its underlying theoretical and clinical processes. Provider Training involves standardizing training between providers and ensuring they are trained to clear criteria and monitored over time. Intervention Delivery involves assessing and monitoring differentiation (differences between the intervention and any comparison treatments), competency (skills set of provider), and adherence (delivery of intended components). Intervention Receipt refers to whether the intervention was understood and 'received' by participants and enactment refers to whether participants used intervention related skills in day to day settings |
| Lamdesman-Ramey, et al | 2019 | PT<br>OT<br>CIMT | PTCL<br>RCT | Videotaped sessions each week for every child and then scoring randomly selected 15 min segments | Standardised Fidelity of Implementation Measur | Not reported in the protocol. Emailed lead author 06/10/19. |

| | | | | | | |
|---|---|---|---|---|---|---|
| | | | | | e (FIRM) developed for the trial. | |
| Lawford, et al | 2019 | PT | Case Study | Training facilitator audited audio-recordings of all consultations, and therapists self-audited 50% of consultations using a tool. | NR | NR |
| Lawrie, et al | 2018 | PA | Pilot RCT | a 2-week formal training in the execution and governance of the protocols, procedures for data collection and recruitment was given to the ward staff and the research assessors. See last column… | NR | NR<br><br>Fidelity defined by: “adherence to protocol (intervention fidelity).<br>to monitor and provide support to increase fidelity with the research protocol, a further visit after 3 months was undertaken and then regular video-conferencing calls were made every 2 weeks, and data were sent via protected email every 2 weeks to Oxford for storage and analysis” |
| Leeuw, et al | 2009 | RH<br>LI | PP<br>FidSt | Method of Assessing Treatment Delivery (MATD) aimed to determine protocol adherence and treatment contamination | MATD<br><br>Cites Pereple tchikova | The assessment of treatment delivery consists of verifying the occurrence of essential components (protocol adherence) and the non-occurrence of prohibited protocol deviations (absence of treatment contamination) as well as verifying sufficient treatment differentiation<br>protocol adherence, referring to the degree to which specific treatment procedures are used by the therapists during actual delivery of treatment [8], competence, which is the skilfulness of the therapists delivering the treatment [8], and differentiation, signifying whether the therapies differ from other treatments on several critical dimensions |
| Lenker, et al | 2010 | RH<br>OT<br>PT | PP | n/a | Moncher | Treatment fidelity is comprised of two components. The first is treatment integrity, the extent to which interventions are implemented as intended for the duration of a study (i.e., each participant receives the intended treatment). The second is treatment differentiation, the extent to which the differences between intervention and comparison conditions are maintained over the duration of study |
| Levy, et al | 2018 | PR<br>PA | PTCL<br>RCT<br>FS | NR | NR | Fidelity assessment: compliance with training protocol, monitoring of intervention delivery competency. |
| Li, et al | 2016 | RH<br>LI | FS<br>RCT | Fidelity check against competency/Rx checklist. | NR | NR<br><br>No definition of fidelity |
| Liu, et al | 2017, 2019 | PT<br>RH | PE<br>RCT | coordinators collectively trained at study initiation and annual collaborator meetings. on-site training as required. Day-to-day support by a clinical coordination team, neurologist and PT. A log of trial interventions kept by the coordinator for each participant for hospital and home visit activities. Intervention patients | NR | NR<br><br>Fidelity: components implemented as per protocol |

| | | | | | | |
|---|---|---|---|---|---|---|
| | | | | (with their caregivers) were encouraged to keep a daily log of rehabilitation activities for 30 days after discharge. | | |
| Liu, et al TaiChi | 2018 | RH<br>LI | PTCL<br>RCT | will be monitored through attendance records for each intervention component (peer support groups, health education seminars, and Tai Chi Ruler exercise) and personal records of the home practice of Tai Chi Ruler exercise kept by each participant. | NR | NR |
| Logan, et al | 2018 | RH<br>PT | PTCL | Evaluated using a trial specific SPIRES checklist that outlined all components of the functional standing frame programme intervention. PT will record the content of their sessions and adverse events in the Case Report Forms; an independent assessor will observe one intervention/one control group session at each of the four Stroke Rehabilitation Units at random timepoints during recruitment and complete a fidelity checklist (during qualitative interviews with PT. | MRC | The Medical Research Council guidance [43] recommends<br>process evaluation and highlights the importance<br>of capturing fidelity (whether the intervention was delivered<br>as intended); dose (the quantity of intervention implemented)<br>and reach (whether the intended target<br>population comes into contact with the intervention, and how). |
| Lotzke, et al | 2019 | PT | FS | Protocol developed and tested for feasibility<br>Observation of treatment delivery by PIs. | MRC<br>NIH-BCC | Fidelity is a process applied in the study design, training the provider, delivering of<br>treatment, receipt of treatment and enactment of treatment<br>skills<br>Rx Receipt: assessing and optimising the degree to which the participant<br>understands and demonstrates the knowledge provided by the intervention). |
| Lousada, et al | 2013 | SLP | RCT | Author and blind observation, checklist. | NR | "observational rating scale recording key elements: duration of session; target sound(s); type of reinforcement used; type of intervention; and main activities used." |
| Macleod, et al | 2018 | LI | FS | Programme implementation (by LCs) was estimated from a structured pro forma completed after every patient contact which recorded actual values or scaled ratings | NR | protocol adherence and acceptability |

| | | | | | | |
|---|---|---|---|---|---|---|
| Malden , et al | 2018 | LI | PTCL | Intervention fidelity will be assessed using questionnaires and interviews with parents and practitioners, observation, and session delivery records. Intervention fidelity. measured by assessing the practitioner's logbook of the number of sessions conducted per week, and structure followed in relation to classroom guides. Observation of delivery will also be undertaken, as will qualitative interviews with practitioners and parents on completion of the intervention. | NR | NR |
| Mars, et al | 2013 | CI<br>Pain | PP<br>FidSt | courses were audio recorded with the consent of participants and these recordings were used to assess and evaluate intervention integrity relating to the key elements prescribed in the COPERS facilitator's manual. | NIH-BCC | intervention delivery or integrity, defined as the monitoring and assessment of behaviours at the point of intervention delivery. The effectiveness of complex interventions may be dependent on the 'skills' of those delivering them. 'Skills' can be characterised by separate but related constructs of adherence and competence. Adherence is defined as: the extent to which a person delivers the essential content, delivery strategies and theories prescribed by the intervention designers and avoids activities proscribed by them. Competence refers to the level of 'skill' demonstrated by those delivering an intervention and may include the ability to respond appropriately to a wide variety of contextual cues.<br>(DOSE NOT PART HERE). |
| Master son-Algar | 2014 | OT | PP | n/a | Realist Evaluat ion, consoli dated framew ork for implem entatio n researc h (CFIR) | The programme theory incorporates four potential mechanisms through which fidelity within the trial can be investigated. These four programme theory areas are (1) the balancing of research and professional requirements that therapists performed in a number<br>of areas while delivering the study interventions; (2) the OTs rapport building with care home staff; (3) the work focused on re-engineering the personal environments of care home patients; and (4) the learning about the intervention within the context of the trial and its impacts over time. |
| Master son-Algar et al | 2017 | SLP | PTCL,<br>PE RCT | Process evaluation: questionnaire, observations, interviews. | NR | process evaluation will be focused on investigating the quality of implementation of the PD COMMinterventions as well as adherence to the outcome evaluation protocol |
| Master son-Algar, et al | 2018 | RH | PP<br>CP | n/a | | researchers should investigate barriers and enablers to implementation by<br>reviewing strategies in place to improve or support the fidelity of the rehabilitation intervention. The process evaluation should review strategies in place to measure 'dose delivered' and 'dose received'. Finally, participant's experiences and acceptability of the intervention should be investigated. To date, it is rare for research studies to provide intervention providers with clear guidance on how to assess which is the 'right amount' of tailoring |
| Master son-Algar, et al | 2014 | OT<br>RH | PP | N/a | CFIR | Consolidated framework for implementation fidelity (or research, CFIF or CFIR): adherence(intervention content, coverage, frequency, and duration), and<br>'moderating factors' that can potentially affect the degree of fidelity. |

| | | | | | | |
|---|---|---|---|---|---|---|
| | | | | | | These include the complexity of the intervention(s) to be implemented, facilitation strategies, quality of delivery and participant<br>responsiveness: context, described as the culture of organizations, social behaviour/interactions among members and<br>social structures, as an additional moderating factor |
| Mathews, et al | 2014 | PA | SR | REAIM | RE-AIM | Implementation refers to whether an intervention was delivered as intended in relation to protocol fidelity, provision of training and support for individuals delivering interventions may improve protocol fidelity. |
| Maxwell, et al | 2017 | RH<br>PT | PTCL | Realist Evaluation to track how the implementation is working (including fidelity to the PFMT protocol). | NR | fidelity or variation to PFMT protocol (e.g., number and type of sessions) and the impact of any variations |
| Mayer-Davis | 2018 | LI | RCT | Coaches received training in motivational interviewing and problem-solving skills training and in the specifics of intervention delivery. Review of 10% of random selection of the audiotaped sessions for assessment of adherence to motivational interviewing principles using the motivation- al interviewing treatment integrity system. | NR | NR<br><br>fidelity of intervention delivery,<br>To ensure intervention fidelity, meaning that the intervention would be delivered as designed |
| McCarthy, et al | 2015 | PA | PE | Audiotaped sessions reviewed by independent assessor for adherence to MI principles. | NR<br><br>Cites Miller and Rollnick | The implementation of the intervention according to protocol, which includes fidelity, dose, and context. Evaluation of the fidelity to the intervention (how closely it was implemented as designed) focused primarily on the incorporation of MI principles into the counselling sessions. Essential elements covered, Adherence to principals<br><br>DOSE NOT INCLUDED IN FIDELITY: Evaluation of the dose of the intervention that was delivered consisted of examining the quantity or amount of intervention delivered to participants. |
| McDowell, et al | 2017 | CR<br>PT | RCT | Various, including weekly phone calls with the research team to discuss individual patient treatment plans and regular training updates | NIH-BCC Borrelli | Design: “treatment fidelity practices relating to design ensure that a study adequately tests its hypothesis in relation to its underlying theoretical and clinical processes”<br>Training providers: “treatment fidelity of provider training involves standardising training between providers, ensuring that providers are trained to criterion, and monitoring and maintaining provider skills over time.”<br>Delivery of treatment: “the assessment and monitoring of treatment fidelity during treatment delivery involves treatment differentiation (did the providers only deliver the target treatment and not other treatments), treatment competency (did providers maintain the skill set learned in training), and treatment adherence (delivery of the treatment components as intended).”<br>Receipt of treatment: “whether the treatment that was delivered to the participant was actually “received” by the participant.”<br>Enactment of treatment skills “assessment, monitoring, and improving the ability of participants to perform |

| | | | | | | |
|---|---|---|---|---|---|---|
| | | | | | | treatment related behavioural skills and cognitive strategies in relevant real life settings |
| McPherson et al | 2018 | RH | | After each coaching session, the coach completed the fidelity measure, which was reviewed by a member of the research team with coaching expertise-Solution-Focused Fidelity Instrument (SFFI) | NR<br><br>Uses SFFI- Dumas et al, 2001 | NR<br><br>SFFI based on: Fidelity refers to delivering the intervention in a "comparable manner to all participants and is true to the theory and goals underlying the research" |
| McPherson et al | 2019 | PA | PTCL RCT | Solution Focused Coaching Fidelity Instrument. | NR | NR<br>Development of the fidelity instrument in a different paper. |
| Miller, et al | 2017 | PT | PTCL cRCT | Fidelity will be measured through an audit of the fidelity checklist and electronic medical record (EMR) of each included patient to determine consistency of the intervention with the protocol | NR<br><br>Cites Hildebrand | consistency of the intervention with the protocol |
| Mitchell, et al | 2018 | SLP | FS RCT | Data about how the intervention was delivered (face to face or independently) and by whom were extracted from therapists' records. | NR | NR |
| Moore, et al, | 2018 | PA | SR, NS | n/a SR | Borrelli<br>Also TIDIER | 1)Treatment fidelity strategies for design of study.<br>Ensure same treatment dose within conditions<br>Ensure equivalent dose across conditions<br>Plan for implementation setbacks<br>2)Treatment fidelity strategies for monitoring and improving provider training<br>Standardise training<br>Ensure provider skill acquisition<br>Minimise "drift" in provider skills<br>Accommodate provider differences<br>3) Treatment fidelity strategies for monitoring and improving delivery of treatment<br>Control for provider differences<br>Reduce differences within treatment<br>Ensure adherence to treatment protocol<br>Minimise contamination between conditions<br>4) Treatment fidelity strategies for monitoring and improving receipt of treatment<br>Ensure participant comprehension<br>Ensure participant ability to use cognitive skills<br>Ensure participant ability to perform behavioural skills<br>5) Treatment fidelity strategies for monitoring and improving enactment of treatment skills<br>Ensure participant use of cognitive Skills<br>Ensure participant use of behavioural skills |
| Moran, et al | 2015 | PT<br>PA | FS | ? | Borrelli | ability of participants to comprehend both the visual and auditory components of the DVD. |
| Morrison, et al | 2017 | RH | FidSt | adherence to the training protocol, all intervention sessions were audiotaped. Samples from the | NIH-BCC | 1) study design, (2) facilitator training, (3) intervention delivery, (4) intervention receipt, and (5) intervention enactment. |

| | | | | | | |
|---|---|---|---|---|---|---|
| | | | | audiotaped intervention sessions were reviewed by one of the principal investigators, who had developed a fidelity checklist to monitor intervention adherence. Facilitator trainers did in vivo observation of at least one session per facilitator. | | |
| Morse, et al | 2017 | PA | FS RCT? | Fidelity Checklist | NR | NR |
| Moseng, Dagfinrud, Østerås | 2019 | PT PA | RCT | Analysed study notes and self-reported questionnaire data. | Carrol | "Bottom-line measure of fidelity within this framework is the **evaluation of adherence.** Adherence should be addressed through the subcategories content and dose. To evaluate adherence in the SAMBA study, six core components representing content and dose of the implementation strategy were classified as either "high adherence", "partly adherence" or "low adherence" The components were: 1) Proportion GPs and PTs attending the workshops; 2) PT knowledge and attitudes after the workshop towards eight statements on evidence-based OA treatment; 3) Number of times the PT adjusted their patients' exercise programs; 4) Proportion of patients who received physiotherapy; 5) Proportion of patients who completed the patient education and exercise period and 6) Proportion of patients who exercised according to dose recommendations from ACSM." |
| Murphy, et al | 2011 | OT | PTCL | standardized modules for the intervention and OT and treatment receipt | NIH-BCC | Fidelity of Treatment Delivery: intervention groups are equivalent in dose and administration of treatment. And fidelity of treatment receipt: participant involvement in treatment. |
| Murphy, et al | 2008 | OT | PILOT | NR | NR<br><br>Cites Bellg | NR<br><br>"Controlling for treatment time is considered necessary for treatment fidelity in intervention studies" |
| Murray ,et al<br><br>Murray et al | 2015<br><br>2019 | PT | RCT | Mentions importance of fidelity, and implementing the communication strategies more closely to protocol. | NR | NR |
| Mustian, et al | 2017 | PA Onco | SR/MA | Treatment fidelity protocol as a variable. | NR | NR |
| Namasivayam, et al | 2018 | SLP | PILOT | Providing therapy cues are crucial for monitoring treatment delivery fidelity | NR Cites Hinkley Borrelli | NR<br><br>Adherence to Rx protocol described. |
| Naylor, et al | 2006 | PA | FS/PP | assessed fidelity by comparing actual to prescribed amount of physical activity delivered, actual versus potential number of weeks logged and by examining the planned and implemented activities coded across the six zones | NR | NR<br><br><br>fidelity to the model |

| | | | | | | |
|---|---|---|---|---|---|---|
| Neel, et al | 2019 | RH<br>PT<br>OT<br>NR | PTCL<br>RCT | highly manualized protocol is monitored and 10% of videos reviewed by an independent reviewer. Standard care is also monitored using nursing logs in both groups. randomly sampled fidelity of treatment (FOT) measures will be collected. checklist of critical elements, self-scored. | NR<br><br>Cites Borrelli | NR<br><br>checklist of critical elements and adherence to protocol described. |
| Nielsen , et al | 2014 | PT | QS | Comprehensive training and mentoring by psychologists. Intervention Training workshop. | | |
| Noble, et al | 2018 | LI | FidSt | Checklist, process evaluation | NR Cites Dane | Fidelity includes exposure, adherence to content and quality of delivery |
| ***Norris, et al*** | ***2015*** | | | | (NIH) model | The US National Institute of Health (NIH) model of fidelity |
| O'Neal et al | 2018 | PR | FS | Fidelity of the PAI was assessed using the checklist published by Borrelli | Borrelli | |
| O'Shea, et al | 2016 | PT<br>BChge | ScR | n/a | Bellg(NIH-BCC) | (1) design of study, (2) training providers (3) delivery of treatment (4) receipt of treatment (5) enactment of treatment skills |
| Ownsworth et al | 2013 | CR<br>ML | PTCL<br>RCT | monitored using a checklist based on Borelli's framework. Sessions will be audiotaped to enable therapists' adherence to the treatment protocol to be examined for a random sample (20%) of sessions by experts who are independent of the study | Borrelli | NR |
| Ownsworth, et al | 2017 | OT<br>RH | RCT | Therapist adherence to the treatment protocol was examined for a random sample (15%) of audiotaped sessions 1 to 8 for each intervention using a checklist based on Borelli's framework. | Borrelli | Delivering core components of intervention according to the manualized treatment protocol |
| Ownsworth, et al | 2013 | OT<br>RH | PTCL<br>RCT | Therapist adherence to the treatment protocol examined for a random sample (15%) of audiotaped sessions 1 to 8 for each intervention using a | Borrelli | Delivering core components of intervention according to the manualized treatment protocol |

| | | | | | | |
|---|---|---|---|---|---|---|
| | | | | checklist based on Borelli's framework. | | |
| Palmer, et al | 2015 | SLP | PTCL RCT | checklist guiding SLP selection of exercises based on the participant language profile identified during assessment. | NR | NR |
| Palsola et al, 2020 | 2020 | PA | FidStd | Semi-structured interviews of providers and participants-analysed with thematic analysis | Bellg/ NIH-BCC | Fidelity refers to the methodological strategies used to monitor and enhance the reliability and validity of behavioural interventions (Bellg et al,2004). However, interventions can be delivered with perfect fidelity, but not work as<br>intended if participants do not accept and engage with them.<br><br>Receipt refers to the participant side of fidelity, that is, comprehension of intervention and performance of the cognitive and behavioural skills taught in the intervention (Bellg et al, 2004) |
| Pastva et al | 2018 | PT RH | FidSt RCT | | NIH-BCC | 1. Ensuring the intervention dose is consistent across participants: Ensure that dose is adequately described and is the same for each participant within each condition and contamination across conditions must be minimized.<br>2 standardizing interventionist training. Ensure that training is conducted similarly across interventionists and maximize acquisition and maintenance of skills and to limit deviation from the standardized procedures over time.<br>3) monitoring intervention delivery. Ensure that intervention is being delivered as intended. Monitoring of intervention delivery involves assessment of intervention competency (did interventionists maintain the skill set learned in training), intervention differentiation (did the interventionists only deliver the target treatment and not other treatments), and intervention adherence (were intervention components delivered as intended).<br>4) evaluating participants' understanding of information provided. Ensure participant comprehends information to attend to and perform study-related skills. This particular aspect focuses on the participant's receipt of the study information, which is demonstrated by his/her ability to attend to and perform the physical rehabilitation exercises.<br>5) ensuring that participants use the skills taught in the intervention. Ensure the participant actually uses the skills provided in the intervention in appropriate life settings. |
| Patterson, et al | 2018 | SLP | Prosp. single cohort | content and treatment plans, recorded in patients' notes were evaluated by a CBT expert practitioner as part of supervision | NR | fidelity were measured by assessing whether the intervention could be delivered as planned, by a SLT with CBT training |
| Pennington, et al | 2019 | SLP | PILOT RCT | Video recordings of 31 Skype dysarthria therapy sessions (19%) were checked for treatment fidelity. | NR | NR<br><br>adherence to the treatment protocol |
| Persch | 2013 | OT | PP | manual of procedures (MOP; operating procedures for the study and for training personnel in the administration of outcome measures | NR | investigators should concern themselves with facets of treatment fidelity related to study design and the training of personnel. Fidelity practices related to study design help investigators discern whether the study will adequately achieve the aims and test the hypotheses that have been set forth. Fidelity in training refers to the extent to which the outcome measures and treatment |

| | | | | | | |
|---|---|---|---|---|---|---|
| | | | | and interventions. Video Rx and regularly scheduled checks of both our outcome assessors and our intervention therapists | | are administered in accord with the MOP. Thus, a team that has multiple protocol violations would be said to have low training fidelity, which would increase variability with which the protocol is administered. |
| Peters, et al | 2018 | HP | CRCT | Process evaluation | NR | NR<br>Fidelity gets heading but outcome not really described with data |
| Pfeiffer, et al | 2011 | OT | PILOT | Accordance with Parham SI Fidelity Measure | NR | NR |
| Poltawski, et al | 2014 | RH | PP | n/a | NHBCC | Study design Describing key ARNI elements and principles (A, B, C)<br>Ensuring intervention meets current best practice guidelines (C, D)<br>Developing intervention manual (A, B, C, D, F)<br>Identify appropriate assessment methods and outcome measures (A, E)<br>Identity process measures that might influence fidelity (A, B)<br>Provider training Identifying key elements of provider training regarding ARNI (B, C)<br>Identifying key elements of provider briefing regarding conduct of trial (A, C)<br>Developing trainer materials, quality standards and minimum experience levels (A, B, C, D)<br>Treatment delivery Distinguish core and flexible components of intervention (A, B, C)<br>Identifying necessary resources to deliver intervention (A, B)<br>Developing fidelity assessment instruments (A, F)<br>Identifying threats to fidelity and possible strategies to mitigate (A,B,F)<br>Treatment receipt Developing study participant information materials (A, C, E)<br>Identifying strategies to promote participant engagement (A, E)<br>Developing fidelity assessment instruments (A, B, F)<br>Treatment enactment Developing participant information materials (A, E)<br>Identify factors influencing adherence and ongoing engagement in intervention (A, B, E)<br>Developing fidelity assessment instruments (A, B, F) |
| Poltawski, et al | 2013 | PA<br>RH | CS<br>FS | participant interviews, audit of participant and EP records, and observation of training. | NR | NR |
| Pozehl, et al | 2010 | CR<br>PA | CT | | Bellg | Intervention fidelity strategies were based on the recommendations of Bellg et al The theoretic basis for specific intervention strategies was an important first step in assuring fidelity. Every participant in the treatment group received the same number of group sessions that were equal in frequency and length. The principal investigator led the group sessions for the treatment and control groups using a protocol developed from the Heart Failure Society of America's educational modules.26 The exercise training protocol was guided by a physical therapist in a cardiac rehabilitation setting during the first 3 weeks of the study. The principal investigator taught resistance training during the first 3 weeks of the study using detailed pictures and guidelines. Receipt of the treatment was assessed during each group session through questioning and verification of participant understanding. The principal investigator or physical therapist observed enactment of skills on a regular basis during the first 3 weeks of the study and every 4 weeks during the remaining 9 weeks. |

| | | | | | | |
|---|---|---|---|---|---|---|
| Pyatak, et al | 2017 | OT<br>LI | PILOT RCT | therapists document their adherence to the intervention protocol in treatment notes for each intervention session. Second, approximately 10% of sessions are observed by another therapist trained in the intervention, who completes a fidelity checklist and provides feedback to the treating therapist. Finally, weekly meetings are held with the full intervention team to facilitate problem-solving and prevent intervention drift. | NR | NR<br><br>intervention adherence, protocol deviations |
| Quinn, et al | 2016 | PT<br>PA | RCT | combination of self-report checklists, independent analysis of audio-recordings, and a self-assessment completed by the intervention coaches. | NR (Social grounded theory?) | |
| Reddington, et al | 2017 | PT | PTCL MMS | Implementation fidelity testing will be carried out in order to assess the treating clinicians are delivering what is intended by the protocol. An independent assessor will review video footage of physiotherapist and participant session in order to assess implementation fidelity. | MRC<br>NIH-BCC | treating clinicians are delivering what is intended by the protocol |
| Reeves, et al | 2017 | PT<br>ExPhy | Pilot RCT | in-person training of the study interventionists. Ongoing oversite of study rehabilitation sessions. Bi-weekly intervention teleconferences among all site intervention leaders and interventionists provide continued monitoring and guidance. | NR | NR |
| Resnick, et al | 2011 | RH<br>EP | FidSt PP | quantification of the sessions attended and activities that occurred within those sessions. delivery of the intervention was qualitatively evaluated based on | NIH-BCC<br>Moncher | when designing a study so as to maximize treatment fidelity, three issues need to be considered: treatment delivery, receipt, and enactment. Delivery focuses on assuring that the intervention was delivered as proposed to all participants. Receipt addresses whether the participant understood the intervention, learned new information, and can perform a new behaviour and therefore expands beyond just exposure or delivery of the intervention to the individual. Enactment seeks to |

| | | | | | | |
|---|---|---|---|---|---|---|
| | | | | 20-random observations using a checklist that included both control and treatment group interventions | | assure that the intervention is performed in real world settings. Treatment fidelity for an intervention should also be evaluated with regard to study design and training of interventionists. Treatment fidelity related to design considers adherence to group assignment and explores the degree to which the theoretical framework on which the intervention was developed is maintained. Design: Adherence to the underlying theory related to exercise and prior work testing similar interventions. Assurance that treatment group was exposed to the treatment intervention and control group exposed to the control intervention and that there was no carryover between groups. Treatment fidelity related to the training of interventionists assures that those implementing the intervention were adequately prepared to do so and implemented all aspects of the treatment as intended. |
| Rich, et al | 2017 | PA | ImpSt | qualitative and quantitative measures tracked over time by peer health coaches with tracking tablet to assesses and maintain the quantity and fidelity of the program delivered. Interviews. | NR | fidelity (quantity & quality of intervention delivered) |
| Robbins, et al | 2016 | PA | FidStd | Process evaluation, survey to reflect extent to which intervention reflects conceptual framework | | Measurement of "fidelity", which can also be accomplished via survey, assists in determining the extent to which the intervention is consistent with the conceptual framework on which it is based. extent to which intervention reflects conceptual framework (DOSE NOT PART OF FIDELITY HERE). Fidelity: theoretical integrity |
| Robinson et al | 2021 | Surg | SR | Evaluates reporting of intervention adherence in surgical trials. | NR | Intervention adherence reporting evaluated in the systematic review, but not defined and no theoretical model/framework cited. |
| Roberts, et al | 2018 | RH | PrEv Randomised FS | Process evaluation | NR | NR<br><br>Measurement of intervention fidelity: completion of workbook tasks, completion of diaries and number and content of therapy sessions.<br>Delivery/ delivery to individuals. What rehabilitation intervention is delivered? Is it what was intended by the researchers? What intervention is delivered to each participant? Is the delivered intervention the one intended by the researchers?<br>Theory: What theory has been used to develop the intervention?<br>Context: What is the wider context in which the feasibility study is conducted?<br>Response of rehabilitation teams/patients? How is the enhanced intervention adopted by the rehabilitation teams? How do the patient participants respond? |
| Robins, et al | 2019 | PA LI/HP | PP CTrial | Manual, provider training, multimedia materials. Observation of Rx and post-Rx interviews. | NIH-BCC | 1) treatment design, (2) training providers, (3) delivery of treatment, (4) receipt of treatment, and (5) enactment of treatment skills. design should ensure that the study hypotheses are tested in relation to both underlying theory and clinical processes. Provider training is focused on assessing and optimizing training processes for intervention delivery. Ensuring fidelity through treatment delivery is focused on processes that ensure the treatment is delivered as designed and focus on standardizing and improving delivery as well as assessing adherence. Processes of treatment receipt involve monitoring and optimizing participant understanding and performance of intervention skills during treatment |

| | | | | | | |
|---|---|---|---|---|---|---|
| | | | | | | delivery. treatment enactment focus on ensuring that cognitive and behavioural intervention elements are applied in relevant daily life situations. |
| Rodgers, et al | 2017 | RH | PTCL RCT | TIDIER items 10-12 Data from robot software and training sessions are periodically reviewed to monitor intervention adherence and feedback is provided to therapy staff delivering the intervention | NR | How well (planned)? 'If intervention adherence or fidelity was assessed, describe how and by whom, and if any strategies were used to maintain or improve fidelity, describe them. How well (actual)? 'If intervention adherence or fidelity was assessed, describe the extent to which the intervention was delivered as planned' |
| Ryan et al | 2016 | PT | PTCL RCT | Fidelity of the resistance training programme to trial protocol will be quantified by observations of exercise sessions. Semi-structured interviews. Process eval. | NR | NR<br>Fidelity of the resistance training programme to trial protocol. |
| Salamh, et al | 2019 | PT<br>PA | SR/MA | Modified NIH-BCC checklist to create customized fidelity Checklist for the systematic review/Meta-Analysis. | Borrelli (NIH-BCC) | "Item 1: Was information about the treatment dose in the intervention condition provided?<br>Item 2: Was information about the treatment dose in the control or comparison condition provided? Item 3: If more than 1 intervention was described, were they all described equally well?<br>Item 4: Were methods used to ensure the dose was equivalent between conditions?<br>Item 5: Were methods used to ensure the dose was equivalent within a condition?<br>Item 6: Were characteristics to be sought and avoided by the treatment provider addressed *a priori*, and was some mention made of credentials? Item 7: Was there a mention of a theoretical model or clinical guidelines on which the intervention was based?<br>Item 8: Did the authors indicate how providers were trained? Did the authors indicate that provider training was standardized? Item 9: Was there a method to ensure that the content of the intervention was being delivered as specified?<br>Item 10: Was there a method to ensure that the dose of the intervention was being delivered as specified?<br>Item 13: Were nonspecific treatment effects evaluated?" |
| Salamh, et al | 2015 | PT | SR/MA | Modified NIH-BCC checklist to create customized fidelity Checklist for the systematic review/Meta-Analysis. | Borrelli (NIH-BCC) | "Item 1: Was information about the treatment dose in the intervention condition provided?<br>Item 2: Was information about the treatment dose in the control or comparison condition provided? Item 3: If more than 1 intervention was described, were they all described equally well?<br>Item 4: Were methods used to ensure the dose was equivalent between conditions?<br>Item 5: Were methods used to ensure the dose was equivalent within a condition?<br>Item 6: Were characteristics to be sought and avoided by the treatment provider addressed *a priori*, and was some mention made of credentials? Item 7: Was there a mention of a theoretical model or clinical guidelines on which the intervention was based?<br>Item 8: Did the authors indicate how providers were trained? Did the authors indicate that provider training was standardized? Item 9: Was there a method to ensure that the content of the intervention was being delivered as specified?<br>Item 10: Was there a method to ensure that the dose of the intervention was being delivered as specified?<br>Item 13: Were nonspecific treatment effects evaluated?" |

| | | | | | | |
|---|---|---|---|---|---|---|
| Salmoiragо Blotcher | 2017 | PA CR | UCT | sessions were video recorded, and the study auditor reviewed 10% of videos for protocol consistency using a checklist developed for the study. | NIH-BCC | Not reported. |
| Sandborgh,et al<br><br>Part I and Part II | 2010 | PT | FidSt | PT training in intervention and how to adapt/tailor. Rx components were operationalized in accordance with the treatment manual. Integrity checklist developed for study: documentation, treatment components categorized as present or not. Treatment documents reviewed/ evaluated by second author not involved in training of PTs, or in supervision intervention phase. | NR<br><br>Cites Pereplechikova | Treatment integrity includes three components: adherence, competence, and differentiation). Adherence is the degree to which the therapist conducted treatment in adherence with the treatment manual; overall and for treatment components. Competence refers to the therapists' level of skill when delivering the treatment. The therapists' degree of competence could account for a potentially large degree of variance not explained by the treatment itself and may vary from patient to patient and depend on many extraneous factors. Differentiation refers to whether treatments in an intervention differ from each other in the intended manner and is closely related to therapist treatment adherence, i.e. that treatments are distinctly different from one another and do not overlap. |
| Schaaf, et al | 2012 | OT | FidSt | Manualisation of the intervention and examination of the treatment manual's adherence to fidelity Random sampling of 20% of the videotapes of the treatment sessions were rated for therapist's fidelity with sensory fidelity checklist. | NR<br><br>Cites Bellg | NR<br><br>fidelity to the manualized intervention. |
| Schaaf, et al | 2014 | OT | RCT | Manualisation of the intervention and examination of the treatment manual's adherence to fidelity Random sampling of 20% of the videotapes of the treatment sessions were rated for therapist's fidelity with sensory fidelity checklist. | NR<br><br>Cites Bellg | NR<br><br>fidelity to the manualized intervention. |
| Schepens, et al | 2012 | OT PA | RCT | OT protocol training module with the PI/written session reports regular check-in phone meetings with PI. | NIH-BCC | NR |
| Scott, et al | 2018 | PA | FS | Three 10-min segments were analysed from separate audio-tapes by an independent coder using checklist/tool. Provider training | NR | Treatment fidelity was assessed, including compliance to intervention delivery. protocol deviations. Assessing fidelity of intervention delivery can optimize intervention effectiveness by identifying and correcting protocol deviations early and help sustain practitioner's skills |

| | | | | | | |
|---|---|---|---|---|---|---|
| | | | | before RX to ensure competence/delivery . | | |
| Sharma , et al | 2019 | CP | RCT | NR | NR | Not eligible- fidelity not assessed, though in protocol |
| Shivon en, et al | 2013 | Surg | RCT | All procedures were standardized and recorded on video | NR | NR<br><br>Adherence to study protocol. |
| Shrubs ole, et al | 2018 | SLP | Pilot cRCT | Intervention delivery checklist (self-reported) | MRC guidan ce on feasibili ty. | Checklist has “core information about each intervention session (such as date, duration and number of participants) and the extent to which the components of each intervention was delivered as planned (e.g., PowerPoint presentation, video of person with aphasia)” (see Appendix D for details) |
| Silveira , et al | 2019 | PA<br>RH | FidSt<br>PTCL | Fidelity monitoring according to NIH-BCC guidance. | NIH-BCC | Study design: focuses on practices that ensure study procedures and implementation are in line with current theory and clinical processes. Study design fidelity goals include ensuring that conditions are congruent with relevant theory and practice, ensuring equivalent treatment dose within and across conditions, and planning for implementation setbacks.<br>Provider training: address preparation for uniform delivery of treatment.<br>delivery of treatment: focuses on ensuring the intervention is delivered as intended. Many of the concerns within delivery of treatment overlap with strategies for training and study design, including controlling for provider differences and adhering to created protocols; however, this area further addresses differences within treatment conditions and minimizes contamination receipt of treatment: involves strategies and monitoring of a participant’s ability to understand and adopt treatment-related behavioural skills and cognitive strategies.<br>enactment of treatment: strategies aimed at monitoring and improving participant ability to perform treatment-related behavioural skills and cognitive strategies in relevant real-world settings. |
| Skidmo re, et al | 2014 | RH<br>Stroke | PS<br>PE | All research intervention sessions were videotaped and rated for fidelity to the respective manualized procedures with validated, tailored fidelity checklist. | NR<br><br>Cites Hildebr and | We examined two facets of fidelity: 1) treatment integrity, and 2) treatment differentiation. To address treatment integrity, independent raters trained in the respective protocols assessed adherence to specified principles in each protocol (yes, no), and competence in execution (inadequate, adequate, exceptional). To address treatment differentiation between the two protocols, raters assessed adherence of both research interventions to the strategy training protocol to determine the degree to which the strategy training sessions adhered to the planned protocol, and the degree to which the attention control sessions did not include elements of the strategy training protocol. |
| Skolask y, et al | 2013 | PT | CT | Audio recordings are made for all telephone calls with all participants to assess the fidelity of the intervention with MI tool/checklist. ongoing monitoring of intervention integrity. Monthly booster sessions. | NR | NR<br><br>Fidelity not defined or reported. |
| Smith, et al | 2019 | PT<br>PA | FS<br>MMS | analysis of PTs clinical notes against a three-point checklist outlining important details and components of intervention to be completed by the PT. | NR | NR<br><br>Fidelity was defined as adherent and competent delivery of the intervention, |

| | | | | | | |
|---|---|---|---|---|---|---|
| | | | | The three-point checklist included: specific pain education; delivery of a loaded exercise programme; and discussion on self-management strategies, | | |
| Söderlund, et al | 2009 | PT Pain | PTCL, RCT | detailed treatment manual and a treatment protocol/ checklist manual has also been developed to guarantee that the treatment will be unchanged during the course of the study. | Bellg | To ensure the same treatment within condition a detailed treatment manual and a treatment protocol/ checklist is used for each patient separately manual has also been developed to guarantee that the treatment will be unchanged during the course of the study. Both IT-based and Face-to-face-based interventions are going to have equal number of treatment sessions/phases to ensure equivalent dose across conditions. 4. One therapist will deliver both group treatments to ensure standardized trained therapist and to minimize contamination between conditions.<br><br>5. Patients can e-mail their questions in IT-group to the therapist. These questions, we believe, will mirror patient's understanding of the treatment. Also, often asked questions and answers will be on the IT-group's home page for all patients in this group to read.6. By asking so called consumer questions, we are recording patients' beliefs and expectations about the intervention and also, if the expectations are fulfilled.7. To ensure that the patients are able to use cognitive and behavioural skills we are applying home exercises in daily activities. These are reported in a diary and always discussed with the patient.8. To ensure that the behavioural components are not given to the standard care-group in the acute stage the therapist follows a strict manual only dealing with physical symptoms and advice given for all patients at the initial visit. 9. The number of intervention contacts (e-mail contacts and Face-to-face group meetings) is reported. |
| Sosnowski et al | 2018 | RH | RCT FS | retrospective recording of care data on a case report form. | NR | Successful adherence to the protocol was defined as the administration of the entire prescribed ABCDE bundle on at least 80% of ventilated days. |
| Sprowson, et al | 2014 | Surg | PTCL RCT | n/a: only effect of options for randomization on RX fidelity discussed. | NR | NR |
| Stephens, et al | 2018 | Surg | PE cRCT | 37-item, online questionnaire, administered at the end of the study period. Sample of interviews audio recorded, and field notes recorded in a diary at the time of observation, or immediately afterwards. | | design of the intervention and the operational elements required for effective delivery. design (or programme) level and the hospital (operational) level. At the design level, adaptability is often essential in ensuring that quality improvement interventions can fit within different contexts. fidelity to key parts of an intervention is also important to maximise likelihood of success |
| Stephens, et al | 2018 | PS | RCT | routine QI programme activity data (records of meeting attendance and use of the web-based resource) data from an exit questionnaire sent to all QI leads and ethnographic | Carroll<br><br>Referenced but NR | NR<br><br>Adaptation/fidelity results discussed. |

| | | | | | | |
|---|---|---|---|---|---|---|
| | | | | data. The 37-item, online questionnaire, administered at the end of the study period. | | |
| Stevens, et al | 2007 | RH<br>PT<br>OT<br>SLP | PP<br>ImpSt | To ensure consistency and accuracy in the delivery of the workshop, we developed information feedback, individual consultations, and detailed outlines of all intervention components and established a timeline to ensure timely delivery of all intervention activities. | NIH-BCC<br>TI | The goal was to achieve a match between the written protocol of the intervention and the research staff's actual delivery of the treatment. Treatment Implementation (TI) framework: ability to deliver the intervention according to a specific and predefined protocol and that participants perceive and understand the treatment as intended. Participants' ability to enact the skills or behaviours outside the intervention or training setting. |
| Strasser, et al<br><br>(follows Stevens, et al) | 2008 | RH<br>PT<br>OT<br>SLP | cRCT | To ensure consistency and accuracy in the delivery of the workshop, we developed information feedback, individual consultations, and detailed outlines of all intervention components and established a timeline to ensure timely delivery of all intervention activities. | NIH-BCC<br>TI | The goal was to achieve a match between the written protocol of the intervention and the research staff's actual delivery of the treatment. Treatment Implementation (TI) framework: ability to deliver the intervention according to a specific and predefined protocol and that participants perceive and understand the treatment as intended. Participants' ability to enact the skills or behaviours outside the intervention or training setting. |
| Stuart, et al | 2009 | PT | FS | PT observation of classes to ensure adherence. | | Fidelity: exercise protocols are being followed. |
| Sturkenboom, et al | 2013 | OT | FS<br>RCT | analysis of protocol adherence. Assessors used an assessment log to register duration of the visit, adherence to the assessment protocol and any irregularities encountered. | NR | NR<br><br>adherence to the protocol and actual treatment delivery (process, content, and time) |
| Sturkenboom, et al | 2016 | OT | PE | Process evaluation with analysis of protocol adherence. | Gearing | The treatment fidelity: the dose, the protocol process adherence and content of treatment delivered compared to the protocol. 'treatment fidelity', which is defined as the extent to which the intended intervention was provided by the therapists.<br><br>NOT PART of FIDELITY: Another concept is 'treatment enactment', the extent to which recipients (i.e. patients and caregivers) apply the interventions in daily life. |
| Swank, et al | 2003 | Surg | RCT | Surgeons were allowed to apply their own techniques within limitations of the protocol. Procedures and adhesion assessments were recorded on video, | NR | NR<br><br>Adherence to protocol. |

| | | | | | | |
|---|---|---|---|---|---|---|
| | | | | and outcomes were reviewed by two surgeons. | | |
| Tang, et al | 2018 | PA | SR | n/a | Carrol, Frank Tomila and Braun | **Carroll**: Conceptual Framework for Implementation Fidelity:<br>Adherence: Content, coverage, frequency, duration.<br>Moderators: Intervention complexity, Facilitation Strategies, Quality of delivery, Participant responsiveness.<br>Identify: essential elements.<br>**Frank: (NIH-BCC)** 1. Intervention design, 2. provider training 3. treatment delivery of 4. receipt of treatment 5. treatment enactment.<br>**Tomika:** Four Step Fidelity Assurance Protocol: 1.Deconstruct program and prepare implementation plan. 2.sponser staff training.3.Monitor using standard checklists. 4.Track participant outcomes. |
| Tarrant, et al | 2018 | SLP | PTCL Pilot RCT | Session check lists, completed by facilitators, will capture whether the main content of the Intervention Manual is being delivered, indicate where flexibility of delivery is permitted (in session structure/content) and allow evaluation of intervention fidelity. Intervention fidelity will be assessed by several methods: singing group attendance, session checklists, observations, and video recordings of selected singing group sessions in the programme. *Researchers assess this.* | NR | NR<br><br>For analysis of intervention fidelity and engagement we will use trainer interview data, session checklists completed by facilitators (that will be part of the Intervention Delivery Manual), participant attendance data, researcher observations and videos of singing sessions. |
| Taylor, et al | 2015 | RH | PTCL RCT | process evaluation will assess fidelity of intervention delivery. A fidelity checklist developed as part of the programme will be used to assess fidelity of delivery of the intended intervention processes. This will be achieved by analysing recordings of all contacts (telephone and face to face) between intervention facilitators and 20 purposively sampled patient participants. | MRC | how well (or otherwise) intervention components are delivered and received and will also allow researchers to describe variability in fidelity of delivery across patients and facilitators. |
| Taylor, et al | 2015 | PA CR | FidStd | heart rate data recorded for these participants (n = 17; 7 females) to illustrate our fidelity assessment method | Resnick | Intervention fidelity refers to the extent an experimental manipulation has been implemented as intended in a comparable manner to all participants. address session attendance and compliance (meeting the prescribed exercise intensity), as this interaction constitutes the dose of the intervention and influences the physiological response to exercise training. to quantify the overall dose |

| | | | | | | |
|---|---|---|---|---|---|---|
| | | | | | | of the intervention, intention to treat fidelity analysis should include all participants irrespective of their attendance and compliance. per protocol fidelity analysis should involve only those participants who attended all of the<br>prescribed sessions. Both approaches are informative in a full exploration of fidelity. |
| Tew, et al | 2016 | PA | SR | TIDIER checklist | NR | NR<br>intervention adherence or fidelity |
| Thakur, et al | 2012 | Surg | ER | Fidelity to intended target. | NR | NR |
| Thomas, et al | 2018 | SLP | UCT<br>FidST | PI assessed parent and clinician treatment fidelity and reliability of perceptual judgements on randomly selected 10 minutes of the practice phase of each clinician-delivered session, and 100% of each parent-delivered home-based session. | NR<br><br>Cites Kaderavek | We can divide the elements of fidelity into perceptual and procedural components. The perceptual component of fidelity is evaluated through measurement of reliability of perceptual judgements of the child's speech. In contrast, fidelity for procedural aspects of the treatment, such as giving feedback after a 3–7second delay. |
| Thompson,et al | 2018 | PA<br>LI | FidSt | Recordings of sessions scored with checklist by authors. | NIH-BCC | Study Design, Provider Training, Treatment Delivery, Treatment Receipt, and Treatment Enactment |
| Toomey, et al | 2016 | PT | FidSt | direct observations, audio recordings, and self-report checklists. The direct observations were conducted using a checklist developed by the research team to assess the fidelity of the delivery of sessions and the treatment dose. | NIH-BCC | Study design-addresses factors that should be considered when designing the trial and are intended to enable the study to adequately assess its hypotheses in relation to the underlying theory and mechanisms of action of the study.<br>Training of providers-aims to ensure and assess that providers are able to deliver the intervention satisfactorily and as intended<br>Treatment delivery -relates to processes that assess and enhance the actual delivery of the intervention so that it is delivered as intended<br>Treatment receipt- involves using strategies to enhance and assess participant knowledge and use of intervention skills and learning during the intervention. It also considers factors that aim to enhance the acceptability of the intervention to the participant<br>Treatment enactment- uses strategies to enhance and assess their actual practice of the intervention skills and knowledge in daily life |
| Toomey, et al | 2015 | PT | RP | NIH-BCC checklist | NIH-BCC | Fidelity practices related to Study Design are factors that should be considered when designing the trial, and are intended to enable the study to adequately assess its hypotheses in relation to the underlying theory and mechanisms of action of the study (e.g. establishing the behaviour change theory underpinning the intervention and outlining proposed methods to assess its implementation), whilst Training of Providers assesses and ensures that the providers *can* deliver the intervention satisfactorily (e.g. procedures put in place to train the providers, and also procedures to assess the effectiveness of this training). The domain of Treatment Delivery relates to the monitoring of actual intervention delivery (e.g. direct observation of intervention sessions to evaluate delivery of the behaviour change theory techniques) whereas Treatment Receipt and Enactment both focus on the recipient of the intervention, or the participant; using strategies to enhance and monitor participant knowledge and use of intervention skills during the intervention (Receipt) and using strategies to enhance and monitor their actual practice of the intervention skills and knowledge in daily life (Enactment). |

| | | | | | | |
|---|---|---|---|---|---|---|
| Tuntland, et al | 2015 | OT<br>RH | RCT | Not adequately monitored. | NR<br>Cites Bellg | NR<br><br>Treatment fidelity, i.e. if the treatment was delivered as intended |
| Tully, et al | 209 | PA | PILOT RCT<br><br>PE | Training/support manual. structured observation of intervention delivery by a member of the research team responsible for mentor training, semi-structured interviews and focus groups with peer mentors and participants as part of the post-intervention follow-up. audio-recorded sessions to assess the content fidelity of<br>delivery. Fidelity checklists. | NR | NR:<br><br>Fidelity: Fidelity of delivery and receipt of intervention |
| Tyson, et al | 2015 | RH<br>PT | RCT | Not assessed | Hennessey ** | fidelity to the treatment protocol. |
| van Bysterveldt | 2010 | SLP | | | | |
| Vaughan-Graham, et al | 2014 | RH<br>PT<br>OT | SR | | Hildenrand | This refers to the extent of standardization of the actual intervention, as well as to the details on who provides the intervention including how they are trained and supervised throughout the study. Intervention fidelity also requires that the appropriate background and experience level of the study therapists is identified and ensured<br>The ability to operationalize and standardize the intervention, as well as quantify the level of skill of the therapist, supervise and evaluate adherence of the intervention<br>The degree to which a therapist implements an intervention under research conditions (treatment fidelity) is dependent upon the extent and operationalization of the intervention and skill level of the therapist.<br>level of training, skill, or evaluation of adherence of the therapists |
| Volkmer, et al | 2018 | SLP | PTCL RCT | Video/audio recording assessment, recording of sessions and analysis of random selection with treatment adherence checklist. Ind.Raters | NR | NR<br><br>measures of fidelity will demonstrate the consistency with which the intervention in delivered.<br><br>an assessment of treatment fidelity to determine necessary levels of SLT training,<br><br>fidelity measures (video recordings, local collaborator adherence questionnaire, and participant feedback questionnaires) |
| von Thiele Schwarz, et al | 2015 | LI | FidSt PP | Fidelity checklist from adapted CFIF | CFIF | Conceptual Framework for Implementation Fidelity. three aspects of fidelity<br>in the framework (content, coverage, and dose) were complemented with a fourth aspect, namely timeliness: i.e. if the intervention is carried out at the right time. fidelity involves assessing adherence, including its subcategories - content, frequency, duration (dose), and coverage. Thus, adherence relates<br>to whether participants have received the active components of the intervention as often and for as long as initially planned. It also relates to whether all the |

| | | | | | | |
|---|---|---|---|---|---|---|
| | | | | | | individuals who should be participating or receiving the benefits of an intervention are reached. |
| Vranceanu, et al | 2019 | Pain | PS<br>FS | Adherence checklist | NR | NR |
| Walker, et al | 2016 | RH<br>LI/HP | FS<br>RCT | 'train the trainer' manual, regular supervision, and checklist monitoring. | NR | strict protocol compliance |
| Wang, et al | 2014 | PT | PILOT<br>RCT | PT monitoring of exercise performance and training intensity to ensure the treatment fidelity | NR | NR<br><br>Fidelity not defined or reported. |
| Watling, et al | 2007 | OT | SST | rated videotapes of random selection of sessions (21%) with SI tool. | NR | NR |
| Watson, et al | 2017 | PA | SR | | | |
| Wells, et al | 2016 | RH<br>SLP | PTCL | Manual | NR | NR |
| Wenborn, et al | 2016 | OT | PTCL<br>RCT | audio record COTiD-UK sessions and transcribe a sample to monitor the occupational therapists' adherence to the intervention, using a checklist derived from the original study. | NIH-BCC | programme adheres to the intervention manual cover five domains: study design, provider training, intervention delivery, intervention receipt, and intervention enactment |
| Wesson, et al | 2013 | PT<br>OT | FS<br>Pilot<br>RCT | Adherence to the intervention protocol was recorded using field notes during each visit and included comments regarding acceptability of study components – whether participants were engaged in the exercise and/or home safety interventions. | | NR<br>Adherence to the intervention protocol.<br>acceptability of study components – whether participants were engaged in the exercise and/or home safety interventions. |
| Westland et al | 2017 | PA<br>LI | PTCL<br>cRCT | nurses allocated to the study arm will randomly audiotape one consultation from among the four consultations. The audiotapes will be coded using a coding list developed specifically for this study, consisting of the content of each of the four consultations and the Behaviour Change Counselling Index | NR<br><br>Cites Bellg | NR |
| Weston, et al | 2017 | PA<br>CR<br>Surg | UCT | detailed evaluation of the exercise sessions | Resnick | Intervention fidelity refers to the extent an experimental manipulation has been implemented as intended in a comparable manner to all participants. address session attendance and compliance (meeting the prescribed exercise intensity), as this interaction constitutes the |

| | | | | | | |
|---|---|---|---|---|---|---|
| | | | | | | dose of the intervention and influences the physiological response to exercise training. An assessment of fidelity permits an<br>Understanding of whether the exercise was performed at the Prescribed intensities, at all study sites and throughout all phases of the study. |
| White, et al | 2019 | PR | FS CRCT | Face to face, phone, smart phone recordings transcribed verbatim to facilitate assessment of intervention fidelity. | NIH-BCC | NR |
| Whitney, et al | 2013 | OT | NR | n/a | Gearing | Intervention fidelity examines the extent to which the intervention is delivered as it was intended. Describing the specific intervention protocol, which might be in the form of a manual, is the first step toward being able to achieve fidelity in adherence to the protocol |
| Wilbur, et al | 2016 | PA | FidSt | Breitenstein's Fidelity Checklist and fidelity manual. digital audio recording assessment, Enactment was measured by assessing participants' self-monitoring of their lifestyle PA prescription | NIH-BCC | fidelity delivery, receipt, and enactment. study design assures that treatment effects are not confounded with extraneous differences between the treatment and control condition. Treatment fidelity related to training of interventionists assures that interventionists are satisfactorily trained to deliver the intervention to the participants. Treatment fidelity related to delivery of treatment considers that the interventionist delivers the intervention as intended. Treatment fidelity of receipt of treatment focuses on exposure of the participant to the intervention and their ability to understand the skills and perform the treatment-related behaviour skills during treatment delivery. enactment is how well the participant can apply the treatment-related behaviour skills. |
| Williamson, et al | 2018 | PT | RCT | structured record of the interventions (treatment log is completed by the physiotherapists and used to monitor fidelity. | NR<br><br>Cites Bellg | Adherence with the intervention (attendance and the participants' engagement with the programme rated by the physiotherapist). |
| Williams, et al | 2015 | PA | cRCT | SEE FRENCH 2011 | Bellg | SEE FRENCH 2011 |
| Wilson, et al | 2009 | PA | PE | Evaluation against essential elements framework. data collected by a trained, *independent process evaluator* using systematic observation of after-school program activities, checklist. To assess dose and fidelity, the process evaluator observed sessions. | essential elements framework,<br><br>SDT* | Fidelity: fidelity and dose (completeness) of implementation. essential elements informed the development of dose (completeness) and fidelity. essential elements framework that defined dose and fidelity or "complete and acceptable delivery" of the ACT intervention. 1) Fidelity (for PA and behavioural skills components)- To what extent was the social environment autonomy supportive? 2) Dose delivered (completeness for all components)-To what extent were all planned components of the program provided to program participants? and 3) Reach-What percentage of the possible target group attends each week of the program? |
| Wilson, et al | 2010 | PA | IMPST | participants given a manual. ite co-ordinator and project director reports | Durlak | Fidelity: fidelity (degree to which the protocol was implemented as planned), the extent to which the intervention has been received by the audience.<br>defining the active ingredients: using theory or past research to delineate the intervention's active ingredients in clear operational terms, which should be guided by beliefs explaining why they should be successful<br>(ii) using good methods to measure implementation: developing an accurate and valid system for assessing implementation. This should include assessing both the fidelity and dose. DOSE NOT PART OF FIDELITY HERE.<br>(iii) monitoring implementation: assessing the program's active ingredients throughout implementation |

| | | | | | | |
|---|---|---|---|---|---|---|
| | | | | | | (iv) relating implementation to outcomes: using implementation data to better understand program effects |
| Winstein, et al | 2013 | OT<br>RH | PTCL<br>RCT | Expert reviewer assesses the digital video footage accompanying documentation for therapist mastery of each ASAP principle implemented during a 1 hr session. Ongoing training/supervision. | NR<br><br>Cites Bellg | NR<br><br>execution of intervention adherence |
| Wong, et al | 2019 | OT<br>RH | FS | Delphi consensus | Cross<br>**<br>Hildebrand | Fidelity' generally refers to the degree to which a programme is delivered as intended by its developers. However, simple adherence to the manual<br>is only part of the picture; there is an increasing<br>call for a distinction between adherence and<br>competence12,13<br>in measuring treatment fidelity. |
| Woolf, et al | 2016 | SLP | FS<br>qRCT | fidelity checklist was developed, which covered each stage of the treatment protocol as described in the manual and recorded any deviations from it. Video of Rx. *Independent assessment.* | NR | The current study coded individual therapist behaviours as compliant/not compliant with the treatment manual. This very stringent procedure showed that there were deviations from the protocol, e.g. because cues prescribed in the manual were omitted or augmented. |
| Wright, et al | 2019 | SURG | | The proportion of management protocol components completed as intended will be assessed using a checklist at the time of preformed silo application and defect closure. The checklist will be completed by the person undertaking the intervention for every neonate included in the study. A second observer, who has been trained in the gastroschisis management protocol, will independently complete the checklist for 50% of the cases. | Cites Schoenwald and Cohen | Fidelity: The proportion of management protocol components completed as intended. protocol uptake and fidelity. compliance with the protocol (fidelity). important to distinguish between non-compliance and purposive adaptations<br><br>Cohen DJ, Crabtree BF, Etz RS, et al: Fidelity versus flexibility: translating evidence-based research into practice. Am J Prev Med. 2008; 35(5 Suppl): S381–9. Schoenwald SK: It's a Bird, It's A Plane, It's … Fidelity Measurement In the Real World. Clin Psychol (New York). 2011; 18(2): 142–7. |
| Yates, et al | 2013 | CR<br>PA | FidSt<br>PP | Checklists to assess congruence with delivery of the components of CR, random observations. study manual was created | NIH-BCC | study design, training providers, delivery, receipt, and enactment of intervention<br>skills. Design: delivery consistent within and across CR clinical sites so that a study can adequately test its hypotheses in relation to the underlying theory and clinical processes. Training: assessment and ongoing evaluation of the training of interventionists. Delivery: intervention is delivered as intended. It refers mainly to actions of the interventionist. Receipt: treatment has been received and understood. Enactment: skills used in real-life settings as intended. fidelity components of design, training, and delivery of the intervention were the |

| | | | | | | |
|---|---|---|---|---|---|---|
| | | | | | | most different from fidelity in typical intervention studies. |
| Yu-Yahiro, et al | 2009 | PA | RCT | Treatment fidelity visits were performed by 2 investigators on 5 different exercise trainers. Trainers were observed a total of 70 times | NR | study design, training providers, delivery of treatment, receipt of treatment, and enactment of treatment skills |
| Zingmark, et al | 2014 | OT | RCT | Participant attendance rate measured, supervision and interviews with therapists. | Borelli/ NIH-BCC | Fidelity measured but never defined or described. E.g.," From our study, the possibility to draw conclusions about feasibility is limited to attendance rate and programme fidelity." |

Study design:

| | | |
|---|---|---|
| CP: Consensus paper | CT: Controlled study (non-randomised) | DS: Development study |
| ER: Experimental Results | FidSt: Fidelity study | FS: Feasibility Study |
| ImpSt: Implementation Study | PS: Pilot Study | PTCL: protocol |
| PE: Process Evaluation PP: | PS: post or pre surgical care. | Perspective or methodological paper |
| RCT: Randm. Controlled Trial | RP: Review paper | SR: Systematic Review |
| UCT: Uncontrolled clinical trial | QS: Qualitative Study | |

Field/discipline:

| | | |
|---|---|---|
| CR: Cardiac rehab | Complex Interventions | CRML: Cognitive Rehab-Motor Learning |
| CBT: Cognitive-Behav.Therapy | EP: Exercise Physiology/ists | LI/HP: Lifestyle/General health, health promotion (with physical component) |
| OT: occupational Therapy | PA: Physical Activity/Exercise | PT: Physical Therapy |
| PR: Pulmonary Rehab | RH: Rehabilitation-mixed | SLP: Speech Therapy |

Other: "*" Paper may contain more detail. n/a: not applicable NR: not reported
